# Exploring Neglected Tropical Diseases in Somalia: A Scoping Review of Research Efforts and Gaps

**DOI:** 10.1101/2025.10.23.25338432

**Authors:** Fardawsa Ahmed, Owen Nyamwanza, Abdulhakim Guled, Caroline Pensotti

**Author notes:** **Corresponding author:** Fardawsa Ahmed.

## Abstract

Neglected tropical diseases (NTDs) comprise 20 chronic and debilitating conditions that affect over 1.7 billion people worldwide, predominantly in marginalized and impoverished communities. In Somalia, the prevalence of NTDs is fueled by limited access to clean water, inadequate sanitation, and insufficient healthcare infrastructure, which disproportionately affect vulnerable groups such as women, children, and rural populations. Prolonged political instability has further impeded efforts to conduct research and establish effective surveillance systems for NTDs. Unlike its neighboring countries, Somalia lacks a comprehensive master plan for NTD prioritization.

This scoping review mapped the existing research on NTDs in Somalia, identified knowledge gaps, and proposed future research and policy priorities. We included 36 studies published between January 1968 and July 2025, which reported eight neglected tropical diseases, with visceral leishmaniasis (38.9%) and schistosomiasis (33.4%) being the most studied. Other diseases, such as soil-transmitted helminthiases, chikungunya, rabies, and mycetoma, remain severely under-researched. Despite being a WHO priority country for leprosy, only two studies on this disease were identified. The studies predominantly used descriptive designs, with 52.8% led by authors outside Somalia, highlighting gaps in local research capacity.

The review underscores the urgent need for systematic epidemiological studies, enhanced surveillance systems, and integration of NTD research into Somalia’s health policies. Addressing these gaps requires building local research infrastructure, promoting community-based interventions, and fostering collaborations between the government, Somali researchers, and international organizations. This evidence-based approach is vital to mitigating the burden of NTDs and improving health outcomes for Somalia’s underserved populations.

## 1. INTRODUCTION

The neglected tropical diseases (NTDs) are a group of 20 major chronic, disabling, and disfiguring conditions that disproportionately affect over 1 billion people, primarily those living in extreme poverty (1). An estimated 1.7 billion individuals require NTD interventions, including preventive and curative measures (2). The epidemiology of NTDs is intricate, often influenced by environmental factors. Many are vector-borne, involve animal reservoirs, and follow complex life cycles, posing significant challenges for public health interventions (3). Approximately 500 million people in sub-Saharan Africa are affected by NTDs (4, 5).

In Somalia, NTDs are both a consequence and a driver of poverty, thriving in environments where access to clean water, sanitation, and healthcare is minimal (6, 7). According to the Somalia poverty analysis generated from the SIHBS-2022, 54.4 percent of the Somali population lives below the poverty threshold, consuming less than $2.06 daily (8, 9). The burden falls heaviest on marginalized groups such as women, children, the elderly, and those in rural or peri-urban settings (10). Notably, in 2019, Somalia reported over 1,000 new cases of leprosy and continues to face endemic challenges from diseases like visceral leishmaniasis (VL) in nine of its regions (6) where VL incidence is 4.98 per 10,000 of the population in endemic areas (11).

Decades of political instability and civil unrest have hampered disease surveillance and research in Somalia (12, 13). While the formation of the Federal Government of Somalia in 2012 has sparked gradual growth in health-related research, studies on NTDs remain sparse (14). Somalia’s National Development Plan prioritizes infectious disease control but is impeded by limited data on the prevalence and impact of NTDs (15). Unlike neighboring East African nations such as Kenya (16), Ethiopia (17), and Uganda (18), Somalia has not created a master plan for the NTD prioritization exercise to guide surveillance, research, and intervention planning.

We conducted a scoping review of Somalia’s neglected tropical diseases research with the aim of **a)** To map the existing research on NTDs in Somalia. **b)** To identify gaps in the current knowledge and research on NTDs in Somalia. **c)** To highlight priority areas for future research and policy action.

Through this review, we seek to build a foundation for evidence-based strategies to address NTDs in Somalia, ensuring research and policy efforts are aligned with the country’s specific needs and challenges.

## 2. METHODOLOGY

This scoping review followed the framework that was created by Arksey and O’Malley (2005), and Levac et al. (2010) and Colquhoun et al. (2014) developed on it, while Peters, Godfrey et al. (2020) added to it in the Joanna Briggs Institute Manual (2020 edition). The steps in the review are: 1) defining the research question, 2) identifying relevant studies, 3) selecting studies, 4) charting the data, 5) collating, summarizing, and reporting the results and 6) additional considerations and implications of findings for research and practice (19).

### 2.1. Search approach

The review used the Boolean operator method to comprehensively search for relevant existing studies. We developed the keywords and MeSH terms in table 1 from the relevant words in the PCC framework and conducted searches on various databases such as Medline, Global Health, CINAHL, Web of Science, PubMed, and Google Scholar. We did not apply geographical, language, age, and time limitations as indicated in the inclusion and exclusion criteria to the search (See Table 2). We also explored the reference lists of the selected articles to uncover additional relevant studies. We also included a few pieces of grey literature from the World Health Organization.

**Table 1:** Main relevant terms and key words used during the study-search process.

| Relevant terms | Key Words |
| --- | --- |
| **Neglected Tropical Diseases** | "Neglected Tropical Diseases" OR<br>"NTDs" OR "tropical diseases" OR<br>"neglected" OR "Buruli ulcer" OR<br>"Chagas disease" OR "Dengue" OR<br>"chikungunya" OR "Dracunculiasis" OR |
|  | “Echinococcosis” OR “Foodborne<br>trematodiasis” OR “Human African<br>trypanosomiasis” OR “Leishmaniasis”<br>OR “Leprosy” OR “Lymphatic filariasis”<br>OR “Mycetoma” OR<br>“Chromoblastomycosis” OR “deep<br>mycoses” OR “Noma” OR<br>“Onchocerciasis” OR “Rabies” OR<br>“Scabies” OR “Ectoparasitoses” OR<br>“Schistosomiasis” OR “Soil-transmitted<br>helminthiasis” OR “Snakebite<br>envenoming” OR<br>“Taeniasis/cysticercosis” OR “Trachoma”<br>OR “Yaws” |
| **Somalia** | “Somalia” OR “Somali” OR “Somaliland”<br>OR “Puntland” OR “Galmudug” OR<br>“Hirshaabelle” OR “South West” OR<br>“Jubaland” |

**Table 2.** Inclusion and exclusion criteria for scoping review of NTDs in Somalia using PCC Framework.

| Inclusion Criteria | Exclusion Criteria |
| --- | --- |
| ▪ **Population:** Studies involving populations from Somalia OR migrants/refugees from Somalia affected by NTDs.<br>▪ **Concept:** Research focusing on the epidemiology, diagnostic | ▪ Studies not focused on NTDs.<br>▪ Studies with no relevant primary data/summary statistics presented<br>▪ Studies for which the full text was unavailable after conducting a search and requesting it from the |
| techniques, vector control, treatment, prevention quality of care, and policy related to NTDs.<br>▪ **Context:** Studies conducted in Somalia or involving Somali populations in any country. | library. |

### 2.2. Screening and data extraction

This study utilized the Preferred Reporting Items for Systematic Reviews and Meta-Analyses (PRISMA) tool for screening. We imported deduplicated records into the systematic review software (Covidence; Veritas Health Innovation, Melbourne, Australia). We further deduplicated the records using Covidence’s built-in feature to find any duplicates that Endnote had overlooked. Two reviewers (FA and AG) independently screened the title and abstract of all studies, with clearly irrelevant studies excluded at this stage. Subsequently, the same reviewers read the full paper to confirm whether they met the eligibility criteria (Table 2). When necessary, we resolved conflicts through discussion or by inviting a third reviewer (ON). A reviewer (FA) used Covidence Extraction 2 to extract data from the selected publications. The retrieved information included the following: publication year, research design/methodology, author affiliation, major results, and NTD focus.

### 2.3. Data analysis

We downloaded the extracted data from Covidence as a CSV file and imported it into SPSS statistical software (IBM Inc.; Chicago, IL) for analysis. We summarised the data using frequency tables. Finally, we summarized the qualitative and quantitative findings from individual papers for all the NTDs studied in Somalia.

### 2.4. Ethics and dissemination

This scoping review used only previously published, publicly available sources; ethics approval and consent were not required.

## 3. RESULTS

### 3.1. General findings

Fig.1 shows the PRISMA flow diagram that depicts the number of records retrieved, screened, and included in the analysis. The literature search identified a total of 378 studies. Following removal of 89 duplicates, the title and abstract of 289 papers was screened and 246 were excluded as being irrelevant at this stage. Of the 43 papers that proceeded to full text review, a further 7 were excluded for different reasons (Fig.1) The final analysis included 36 papers, with the publication years ranging from **1968 and 2024; no eligible publications** were identified for **January–July 2025**. Additionally, Detailed study characteristics are provided in **S2 Table**.

**Figure 1:**
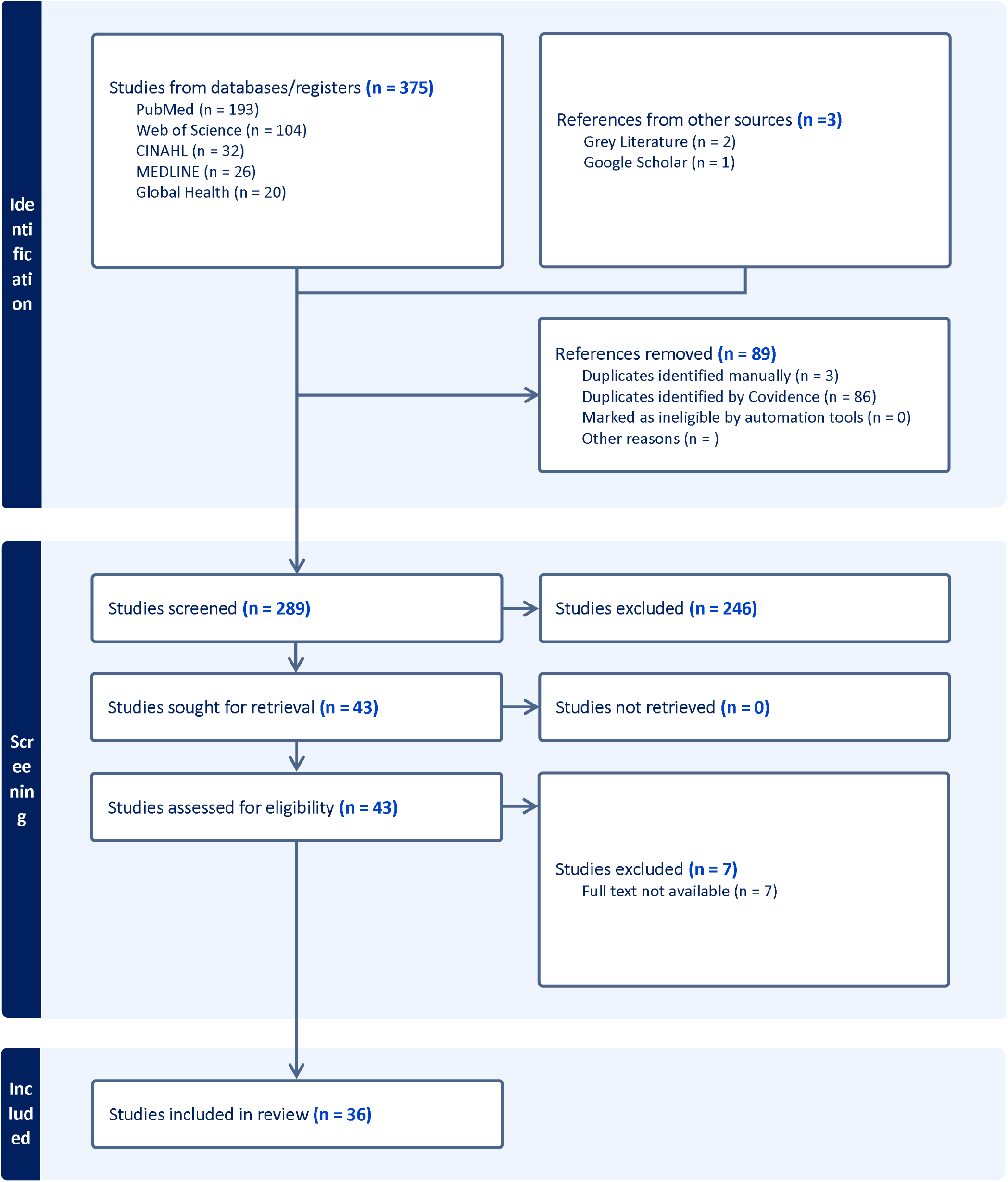
Prisma flow diagram illustrating the relevant studies selection process.

**Figure 2:**
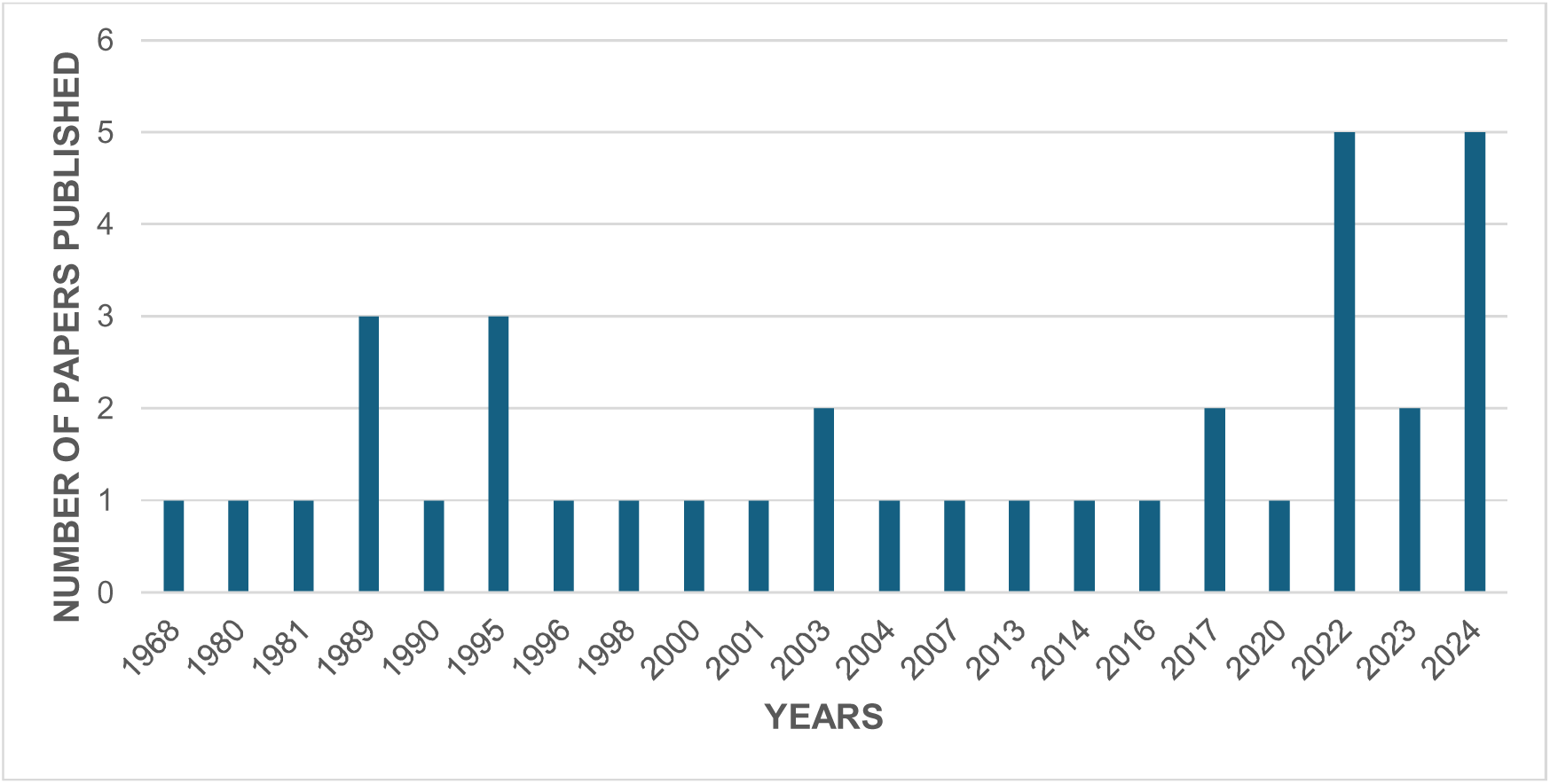
Number of publications on NTDs in Somalia, by year of publication.

#### 3.1.1. Disease focus

Overall, 8 neglected tropical diseases were reported across 36 papers. Table 3 shows the number of publications by disease. Most studies focused on Visceral leishmaniasis (n=14), Schistosomiasis (n=12), Dengue fever (n=5). The remaining diseases were reported in less than 5% of studies. The key findings for each of these 8 neglected tropical diseases are presented later.

**Table 3.**
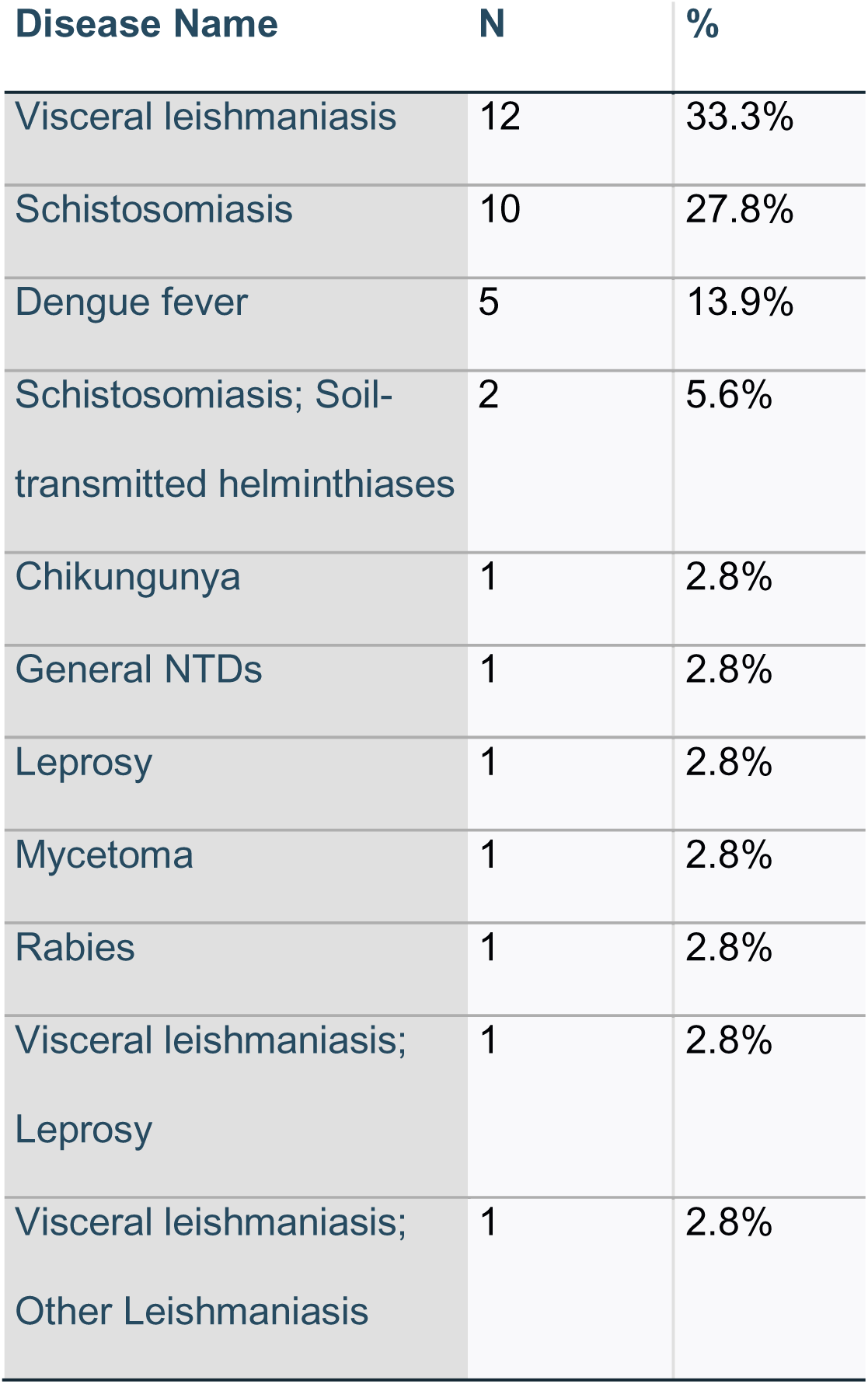
Number of publications on NTDs in Somalia, by disease (*n* = 36)

| Disease Name | N | % |
| --- | --- | --- |
| Visceral leishmaniasis | 12 | 33.3% |
| Schistosomiasis | 10 | 27.8% |
| Dengue fever | 5 | 13.9% |
| Schistosomiasis; Soil-transmitted helminthiases | 2 | 5.6% |
| Chikungunya | 1 | 2.8% |
| General NTDs | 1 | 2.8% |
| Leprosy | 1 | 2.8% |
| Mycetoma | 1 | 2.8% |
| Rabies | 1 | 2.8% |
| Visceral leishmaniasis; Leprosy | 1 | 2.8% |
| Visceral leishmaniasis; Other Leishmaniasis | 1 | 2.8% |

#### 3.1.2. Geographic focus

Table 4 provides an overview of the geographical distribution of data collection and laboratory analysis for publications on neglected tropical diseases (NTDs) in Somalia. The majority of the studies were conducted within Somalia, with 26 publications involving data collection and 19 utilizing laboratory analysis within the country, underscoring the focus on in-country research efforts. A smaller number of studies involved laboratory analysis or data collection in other countries, including the United States (2 studies each), Kenya (2 and 1, respectively), Ethiopia (1 study each for both activities), and Italy (1 study for each activity). Notably, there were instances where laboratory analysis was conducted in multiple countries, such as Kenya and Belgium.

**Table 4.** Number of publications on NTDs in Somalia, by country of data collection and laboratory analysis (n = 36)

| Country | Laboratory Analysis | Data Collection |
| --- | --- | --- |
| Somalia | 19 | 26 |
| United States | 2 | 2 |
| Kenya | 2 | 1 |
| Ethiopia | 1 | 1 |
| Italy | 1 | 1 |
| Not applicable | 4 | 5 |
| Sweden | 1 |  |
| Belgium | 1 |  |
| Kenya; Belgium | 1 |  |
| Not specified | 4 |  |

#### 3.1.3. Study design and methods

Among 36 studies on NTDs in Somalia (Table 5), descriptive epidemiology dominated, comprising 58.3% (n = 21) of the studies. Observational and experimental epidemiology were less common, at 8.3% (n = 3) and 5.6% (n = 2), respectively, while 13.9% (n = 5) lacked clear methodological reporting. There were no studies that employed social science methods, economics, or program evaluation.

**Table 5.** Number of publications on NTDs in Somalia, by study design (n = 36) Category.

| Category | N | % |
| --- | --- | --- |
| Descriptive epidemiology | 21 | 58.3% |
| Not specified | 4 | 11.1% |
| Observational epidemiology | 3 | 8.3% |
| Experimental epidemiology | 2 | 5.6% |
| Grey Literature (Policy analysis) | 3 | 8.3% |
| Descriptive epidemiology;<br>Experimental epidemiology | 1 | 2.8% |
| Narrative review | 1 | 2.8% |
| Not applicable | 1 | 2.8% |

Table 6 shows that serological testing was the most common laboratory method, used in 33.3% (n = 12) of studies, followed by pathobiological methods at 25% (n = 9). Notably, 19.4% (n = 7) did not specify laboratory methods, highlighting gaps in methodological transparency and diversity.

**Table 6.** Number of publications on NTDs in Somalia, by laboratory methods used. (n = 36)

| Type | N | % |
| --- | --- | --- |
| Serology | 12 | 33.3% |
| Pathobiology | 9 | 25.0% |
| Not specified | 7 | 19.4% |
| Not applicable | 5 | 13.9% |
| Taxonomic Classification | 2 | 5.6% |
| Serology; Pathobiology | 1 | 2.8% |

#### 3.1.4. Authorship

Table 7 highlights the representation of Somali researchers as first authors in studies related to the discussed topics. Of the total publications, 38.9% (n=14) had first authors originating from Somalia, while 52.8% (n=19) originated from outside Somalia.

**Table 7.** Proportion of Publications with First Authors from Somalia (n=36)

| Category | N | % |
| --- | --- | --- |
| No | 19 | 52.8% |
| Yes | 14 | 38.9% |
| N/A | 3 | 8.3% |

### 3.2. Disease specific findings

#### 3.2.1. Overview of research focus areas

Table 8 highlights the distribution of studies based on their focus areas. The majority of the reviewed studies (52.8%) (n=19) were primarily focused on epidemiological aspects, emphasizing the prevalence and patterns of NTDs in Somalia. Approximately 30.6% (n=11) of the studies combined epidemiology with clinical management, reflecting efforts to link disease patterns with patient care approaches. Policy-oriented studies were comparatively limited, accounting for only 11.1% (n=4) of the total, while those integrating both epidemiology and policy aspects constituted the smallest category (5.6%) (n=2).

**Table 8:** Distribution of Reviewed Studies by Primary Focus Areas (n=36) Area.

| Area | N | % |
| --- | --- | --- |
| Epidemiology; Policy | 2 | 5.6% |
| Policy | 4 | 11.1% |
| Epidemiology; Clinical Management | 11 | 30.6% |
| Epidemiology | 19 | 52.8% |

#### 3.2.2. Visceral leishmaniasis

Visceral leishmaniasis (VL), also known as *kala-azar*, was the most studied disease, accounting for 38.9% (n=14) of all NTD-related studies in Somalia. Reports of VL span from outbreaks between 1965 and 2006 in regions such as Middle Shebelle and Lower Jubba (20, 21) to more recent case series (2013–2019) from northern Somalia, specifically in Bosaso, Puntland (22).

The first documented clinical cases of VL were reported in 1965 by Baruffa, who identified nomadic populations in the Middle Shabelle region as being affected. This was further substantiated by Cahill et al. in 1967, who confirmed endemic VL foci in the same region (23, 24). Despite the high burden of VL in Somalia, treatment facilities remain concentrated in only three centers in Bakool and Bay, which are supported by WHO. Most patients from rural areas face significant barriers to accessing treatment (6, 25). Additionally, regardless of the high prevalence of VL seropositivity reported to be as high as 23% in one village (23), epidemiological data on leishmaniasis in Somalia remains scarce (26). Between 2002 and 2009, Médecins Sans Frontières (MSF) played a pivotal role in controlling visceral leishmaniasis (VL) in Somalia, particularly through their project in Xuddur, Bakool region, during a significant outbreak. Program data from this period indicates an overall case fatality rate (CFR) of 4.5%, with 88% of patients successfully cured between 2002 and 2008 (20, 26, 27). Unfortunately, due to escalating insecurity in the region, MSF was forced to close the Xuddur project in 2009 (26), leaving a critical gap in VL management efforts.

More recently, a 2022 case report highlighted a 19-year-old male patient from El-Barde District (Bakool Region) presenting with acute renal failure as a complication of untreated VL (28). Another case report emphasized the diagnostic challenges of visceral leishmaniasis (VL) in the context of co-infection with malaria. The overlap in clinical features, such as prolonged fever and splenomegaly, often leads to diagnostic confusion, with malaria frequently being misdiagnosed instead of VL (29).

#### 3.2.3. Schistosomiasis

Schistosomiasis was the second most studied NTD in Somalia, representing 33.4% (n=12) of the studies reviewed. Most research on this disease was conducted between 1968 and 2004 (30–32). However, there has been a marked decline in recent studies, despite a 2016–2017 mapping exercise by WHO and Ministry of Health and Human Services of Somalia, which found unexpected cases of intestinal schistosomiasis caused by *Schistosoma mansoni* in Southwest State and Banadir (33).

In 1976, a WHO-assisted Schistosomiasis Control Project was launched in Somalia, focusing on Koryole as the pilot area and Merca as an additional site. The overall prevalence was 28.8%, with Middle Juba having the lowest rate (25.4%) and Middle Shebelli the highest (42.1%), a statistically significant difference (P<0.014) (34). Moreover, a 1968 study conducted in three locations along the Webbi Shebelle Giohar, Mahaddei Uen, and El Ad-El Gambole, detected schistosome antibodies in 36.6% of participants using the cholesterol-lecithin (CL) test, 56.5% using the bentonite flocculation (BF) test, and 56.1% using the indirect fluorescent antibody (IFA) test. Initial observations also identified potential snail vectors, including *Bulinus* and *Physopsis* species, in the canals of Giohar. However, comprehensive malacological studies were needed to confirm the role of these snails in disease transmission (30). Another study, conducted in 1988 in Sigaale village, located in a fertile region along the Shabelle River, reported an alarmingly high prevalence of *Schistosoma haematobium* infection at 89%. The prevalence and infection intensity were particularly pronounced among individuals aged 10 to 14 years (32). Despite these findings, there remains a critical gap in updated epidemiological studies on schistosomiasis in Somalia.

A rare case of spinal cord schistosomiasis (neuroschistosomiasis) was reported in 2022, involving a life-threatening complication in a 4-year-old child due to untreated schistosomiasis (35).

#### 3.2.4. Dengue Fever

Dengue fever accounted for 13.9% (n=5) of the studies reviewed. The first documented dengue fever outbreak in Somalia occurred in 1985 in Hargeisa (36). In June 2011, an outbreak of acute febrile illness (AFI) among AMISOM peacekeepers in Mogadishu resulted in three deaths, with 82% of tested samples confirming dengue virus (DENV) infections. Enhanced surveillance at military hospitals from June to August 2011 identified 62% of 134 samples as DENV-positive via RT-PCR, with DENV-1 as the dominant serotype. Co-infections with multiple serotypes were observed. Most patients were young males, with 60% hospitalized, 87% experiencing leucopenia, 83% thrombocytopenia, and 13% showing hemorrhagic symptoms. The outbreak underscored the co-circulation of multiple DENV serotypes, the severity of dengue infections, and concerns about under-reported cases and potential spread to other African nations (37).

More recent research highlights the recurrence and growing burden of dengue fever outbreaks across Somalia, such as a recent retrospective study on the 2024 dengue fever outbreak in Puntland State, Somalia, analyzed 956 suspected cases and confirmed 118 dengue-positive patients. Geographically, 43.1% of cases were from the Nugal region, with 38.1% residing in the Garowe district and 5% in the Burtinle district. Other affected regions included Mudug (16.1%, Galkayo district), Sool (15.3%, Las Anod district), and Karkar (13.6%, Gardo district). Additionally, 9.3% of cases were reported from Bari (Bossaso district), and 2.5% from Sanag (Dhahar district) (38). Additionally, a study of 735 febrile patients in Banadir, Somalia, found that 10.8% tested positive for dengue virus (DENV) IgM antibodies and 11.8% for DENV NS1 antigen, with fever and myalgia being the most common symptoms. Most patients were children aged 14 and younger. The study confirmed a dengue fever outbreak in Banadir, highlighting Deynile, Hodan, and Wadajir as the most affected districts (39).

#### 3.2.5. Leprosy

Leprosy, also known as Hansen’s disease, is recognized as a high-prevalence NTD in Somalia. However, it was discussed in only 5.6% (n=2) of the included studies. One study analyzed leprosy detection rates and disability outcomes from 2015 to 2021 (40), highlighting a significant increase in newly detected cases from 107 in 2015 to 2,638 in 2021.

The other paper, a policy analysis (41), discusses the joint efforts of the World Health Organization (WHO) and the Somalia Ministry of Health and Human Services to combat leprosy and VL in Somalia. In 2015, Somalia’s Ministry of Health reportedly set up an NTD section that helped improve detection of new leprosy cases in the country. As a result, there were 2638 newly detected cases in 2021, compared with 107 in 2015. However, the WHO Global Leprosy Programme designated Somalia as a priority country in 2019 after it reported more than 1000 new cases of the disease (6).

#### 3.2.6. Soil-transmitted helminthiases

Despite soil-transmitted helminthiases being one of the four most prevalent neglected tropical diseases in Somalia, alongside leprosy, schistosomiasis, and visceral leishmaniasis (6), it is severely under-researched. One grey literature source from WHO discusses the disease in general, indicating that most cases are concentrated in Southwest State. Between 2017 and 2020, WHO supported mass drug administration (MDA) campaigns, providing treatment to nearly 2.9 million individuals (6).

The only research confirming the disease’s presence was conducted in the United States in 2000. It documented intestinal parasites in Somali Barawan refugees, with 38% of 1,500 individuals tested found to have intestinal parasitic infections.

#### 3.2.7. Rabies

A 2024 case report described a tragic instance of rabies involving a 14-year-old boy from rural Somalia, bitten by a honey badger—a rare rabies vector. The boy presented to a hospital four weeks after the bite with advanced symptoms, including hydrophobia and agitation. Tragically, his younger sister, bitten by the same animal, had died two weeks earlier without receiving post-exposure prophylaxis (PEP). Both children lacked access to timely medical care, highlighting significant healthcare gaps in the region (42). This case underscores the critical importance of early administration of PEP following any animal bite, especially from wildlife, and raises awareness about the potential rabies risk posed by less common vectors like honey badgers.

#### 3.2.8. Mycetoma

A 2024 case series analyzed 10 cases of mycetoma, a chronic and disabling fungal infection. Most patients were middle-aged men from rural areas engaged in farming or livestock care. Lesions primarily affected the lower extremities, with one case involving thoracolumbar (TL) spine lesions. Diagnosis was made through biopsy, and surgical treatment was required in some cases. Mycetoma appears to be particularly prevalent in central Somalia among agricultural workers or livestock breeders (43).

#### 3.2.9. Chikungunya

Chikungunya virus (CHIKV) infections have been indirectly linked to Somalia through case report from Italy in 2016. Two Somali Italian travelers returning from Mogadishu were confirmed to have chikungunya fever, with reports of similar symptoms among their relatives in Somalia (44). Despite these anecdotal findings, no studies directly assess the prevalence or epidemiology of chikungunya in Somalia.

## 4. DISSCUSSION

This scoping review reveals significant knowledge gaps and critical challenges in addressing neglected tropical diseases (NTDs) in Somalia, a country severely affected by poverty, fragile healthcare systems, and prolonged political instability (13, 45). The 36 studies included in this review highlight the disproportionate focus on a subset of NTDs, namely visceral leishmaniasis and schistosomiasis, while other diseases such as soil-transmitted helminthiases, chikungunya, rabies, and mycetoma remain underexplored. This disparity underscores the fragmented nature of research efforts and the urgent need for a more comprehensive approach to NTD research in Somalia. Our findings are consistent with prior review conducted in sub-Saharan Africa, suggesting that the overall burden of neglected tropical diseases (NTDs) in Africa may be significantly underestimated (4).

The predominance of studies on visceral leishmaniasis and schistosomiasis reflects the historically recognized burden of these diseases in the country. For example, visceral leishmaniasis studies dating back to the 1960s have documented endemic regions and highlighted critical gaps in treatment access, particularly in rural areas. Recent studies, however, remain scarce and often focus on case reports, leaving significant gaps in epidemiological data and diagnostic capacity. Similarly, despite the high prevalence of schistosomiasis reported in earlier studies, updated prevalence data is limited, and no recent large-scale control initiatives have been documented. The decline in recent research suggests a stagnation in efforts to understand and mitigate these diseases. A striking finding is the paucity of research on other WHO-recognized NTDs, despite their known presence in Somalia and neighboring countries. For instance, leprosy is a WHO priority in Somalia, yet only two studies addressed this disease, and no studies employed social science or program evaluation methods to understand barriers to diagnosis, treatment, or stigma. Likewise, diseases like rabies, soil-transmitted helminthiases, and chikungunya, which are prevalent in resource-limited settings (3, 46), remain underrepresented in Somali research. This lack of diversity in research focus hampers efforts to design effective interventions for the full spectrum of NTDs affecting the country.

The review also underscores systemic challenges in local research capacity and infrastructure. With 52.8% of studies led by non-Somali researchers, there is a clear need to strengthen the role of Somali researchers in NTD research. Limited local leadership not only impacts the relevance and sustainability of research but also hinders the translation of findings into actionable policy (47). Enhancing local capacity through training, infrastructure development, and partnerships with international organizations is imperative for sustainable progress. Moreover The minimal involvement and/or recognition of local researchers as authors likely mirrors wider patterns in academic publishing, where researchers from Africa are often underrepresented (48).

Methodological limitations further constrain the utility of existing studies. Over half of the included studies were descriptive, with few employing robust epidemiological or experimental designs. Notably, there was no use of social science or economic approaches, which are essential for understanding the broader social determinants of NTDs and the economic impact of interventions. The lack of methodological diversity and transparency, such as unspecified laboratory techniques in some studies, weakens the evidence base needed to inform policy and program development. Additionally, It is well-documented that data challenges related to NTDs remain a global issue (49, 50). Policy-oriented research was another critical gap identified. While neighboring countries like Kenya (16) and Ethiopia (17) have implemented master plans for NTD prioritization, Somalia lacks a comprehensive strategy for NTD control and surveillance. Only 11.1% of studies in this review focused on policy, and there were no comprehensive evaluations of existing health system capacities or community-based interventions. Given Somalia’s unique sociopolitical and environmental challenges, developing a context-specific NTD master plan should be a priority.

The findings of this review have several implications for future research and policy. **Firs**t, there is a pressing need to prioritize systematic epidemiological studies to map the current prevalence, risk factors, and geographic distribution of NTDs in Somalia. Integrating these studies with capacity-building initiatives, such as training Somali researchers and establishing local laboratories, will help address critical infrastructure gaps. **Second,** expanding research to underexplored NTDs and employing diverse methodologies, including social science and economic evaluations, will provide a more comprehensive understanding of the disease burden and the effectiveness of interventions. **Third,** fostering collaborations between Somali researchers, government bodies, and international partners is essential to establish a coordinated approach to NTD research and policy development.

Finally, this review highlights the importance of embedding NTD research within Somalia’s broader health and development agenda. Addressing NTDs requires not only biomedical solutions but also interventions targeting the social determinants of health, such as poverty, sanitation, and education. Community-based approaches and participatory research methods can empower local communities and ensure that interventions are culturally appropriate and sustainable.

## 5. Strengths and Limitations

This review is the first to map neglected tropical disease (NTD) research in Somalia, drawing on both peer-reviewed and grey literature and following PRISMA-ScR guidelines. It provides a transparent overview of available evidence and identifies key gaps.

Nonetheless, limitations exist. Most studies focused on visceral leishmaniasis and schistosomiasis, while other NTDs were rarely addressed. Research designs were largely descriptive, with limited methodological diversity and frequent gaps in reporting laboratory methods. Less than half of the publications had Somali first authors, reflecting restricted local leadership. The review was not prospectively registered, and newer studies may not have been captured.

Overall, the findings provide a useful foundation but also underline the need for more diverse, locally led, and methodologically robust research on NTDs in Somalia.

## 6. CONCLUSION

This scoping review highlights the significant burden of NTDs in Somalia and exposes critical gaps in research, surveillance, and policy. Among the 36 included papers, eight NTDs were identified, with visceral leishmaniasis and schistosomiasis receiving relatively more attention. However, others, including soil-transmitted helminthiases, rabies, chikungunya, and mycetoma, remain severely under-researched. The predominance of external authorship and the limited application of advanced study designs emphasize the pressing need for robust investments in local research capacity and infrastructure.

To address the multifaceted challenges posed by NTDs, Somalia must implement a comprehensive approach that prioritizes research, strengthens surveillance systems, and integrates NTD interventions within broader public health frameworks. Collaborative efforts involving the government, local researchers, and international partners are crucial for developing sustainable strategies to reduce the burden of NTDs and improve health outcomes for the nation’s most vulnerable populations.

## Supporting information

S1_Searching strategy and results

S2_Included Papers Characteristics

S3_PRISMA ScR Checklist

S4_PRISMA -P-Checklist

## Data Availability

All supporting materials are uploaded as supplementary files with this submission.

## 7. Author Contributions

Fardawsa Ahmed: Conceptualization, Methodology, Data collection, Formal analysis, Writing-original draft, Project administration.

Owen Nyamwanza: Supervision, Methodology, Validation, Writing - review & editing. Abdulhakim Guled: Data curation, Visualization, Writing - review & editing.

Caroline Pensott: Methodology, Writing - review & editing.

All authors read and approved the final manuscript.

## 10. Ethics Statement

This study is a scoping review that synthesizes evidence from previously published and publicly available sources. No new human participants, patient data, or animal subjects were involved; therefore institutional ethics approval and informed consent were not required. The review followed recognized scoping_-_review guidance (Arksey & O’Malley; Levac et al.; JBI) and PRISMA-ScR reporting. Where applicable, we relied on the ethics approvals and consent procedures reported within the original studies.

## 11. Data Availability Statement

All data underlying the results are available within the manuscript and Supporting Information files (S1–S3). The full search strategies are provided in S1, and the data-charting sheet is provided in S2. No restrictions apply.

## 12. Supporting Information

- **S1 File. Full search strategies.** Complete database strings (MEDLINE, Global Health, CINAHL, Web of Science, PubMed, Google Scholar) with search dates and limits.
- **S2 Table. Study characteristics of included studies.** Data-charting sheet (variables extracted, definitions, and study-level details).
- **S3 File. PRISMA-ScR checklist.** Completed reporting checklist for scoping reviews.

