## Supplementary material for "Exploring Neglected Tropical Diseases in Somalia: A Scoping Review of Research Efforts and Gaps": S1_Searching strategy and results

### S1 Appendix. Full Search Strategy and Results

This appendix provides the detailed database-specific search strategies and retrieved records. Each section is organized by database and presented in a structured table.

#### Database: Global health NTDs

Total records retrieved: 20

| Year | Title | Journal | Keywords | DOI | URL |
| --- | --- | --- | --- | --- | --- |
| 1980 | The tropical diseases | Somali Range Bulletin | bilharzia |  | https://lstmed.idm.oclc.org/login?url=https://search.ebscohost.com/login.aspx?direct=true&AuthType=sso&db=lhh&AN=19802604192&site=ehost-live&scope=site |
| 2003 | Special issue on Tropical Diseases Research | Eastern Mediterranean Health Journal | animal-parasitic nematodes |  | https://lstmed.idm.oclc.org/login?url=https://search.ebscohost.com/login.aspx?direct=true&AuthType=sso&db=lhh&AN=20053064500&site=ehost-live&scope=site |
| 2015 | Visceral leishmaniasis: control strategies and epidemiological situation update in East Africa: report of a WHO bi-regional consultation Addis Ababa, Ethiopia, 9-11 March 2015 | Visceral leishmaniasis: control strategies and epidemiological situation update in East Africa: report of a WHO bi-regional consultation Addis Ababa, Ethiopia, 9-11 March 2015 | health programs | https://apps.who.int/iris/handle/10665/190168 | https://lstmed.idm.oclc.org/login?url=https://search.ebscohost.com/login.aspx?direct=true&AuthType=sso&db=lhh&AN=20163391986&site=ehost-live&scope=site |
| 2016 | Global, regional, and national incidence, prevalence, and years lived with disability for 310 diseases and injuries, 1990-2015: a systematic analysis for the Global Burden of Disease Study 2015 | Lancet (British edition) | animal-parasitic nematodes | https://www.sciencedirect.com/science/article/pii/S0140673616316786 | https://lstmed.idm.oclc.org/login?url=https://search.ebscohost.com/login.aspx?direct=true&AuthType=sso&db=lhh&AN=20163397734&site=ehost-live&scope=site |
| 2023 | Improved leprosy elimination efforts in Somalia, 2015-2021: achievements in a challenging environment and the way forward | Leprosy Review | subsaharan Africa | 10.47276/lr.94.2.98 | https://lstmed.idm.oclc.org/login?url=https://search.ebscohost.com/login.aspx?direct=true&AuthType=sso&db=lhh&AN=20230265616&site=ehost-live&scope=site |
| 2024 | Epidemiology of Echinococcus granulosus sensu lato in the greater Horn of Africa: a systematic review | PLoS Neglected Tropical Diseases | Echinococcus canadensis | 10.1371/journal.pntd.0011894 | https://lstmed.idm.oclc.org/login?url=https://search.ebscohost.com/login.aspx?direct=true&AuthType=sso&db=lhh&AN=20240049332&site=ehost-live&scope=site |
| 2016 | Leishmaniasis | Neglected tropical diseases - Sub-Saharan Africa | parasitosis | https://link.springer.com/chapter/10.1007/978-3-319-25471-5_5 | https://lstmed.idm.oclc.org/login?url=https://search.ebscohost.com/login.aspx?direct=true&AuthType=sso&db=lhh&AN=20183261358&site=ehost-live&scope=site |
| 2008 | Complexities of assessing the disease burden attributable to leishmaniasis | PLoS Neglected Tropical Diseases | animal-parasitic nematodes | 10.1371/journal.pntd.0000313 | https://lstmed.idm.oclc.org/login?url=https://search.ebscohost.com/login.aspx?direct=true&AuthType=sso&db=lhh&AN=20093042828&site=ehost-live&scope=site |
| 2020 | Clinical characteristics and treatment of actinomycetoma in northeast Mexico: a case series | PLoS Neglected Tropical Diseases | etiology | 10.1371/journal.pntd.0008123 | https://lstmed.idm.oclc.org/login?url=https://search.ebscohost.com/login.aspx?direct=true&AuthType=sso&db=lhh&AN=20203149536&site=ehost-live&scope=site |
| 2018 | Visceral leishmaniasis in selected communities of Hamar and Banna-Tsamai districts in Lower Omo valley, South West Ethiopia: sero-epidemological and Leishmanin Skin Test surveys | PLoS ONE | direct agglutination | 10.1371/journal.pone.0197430 | https://lstmed.idm.oclc.org/login?url=https://search.ebscohost.com/login.aspx?direct=true&AuthType=sso&db=lhh&AN=20183233081&site=ehost-live&scope=site |
| 2003 | Imported dengue in French University Hospitals: a 6-year survey | Journal of Travel Medicine | arthropod-borne viruses |  | https://lstmed.idm.oclc.org/login?url=https://search.ebscohost.com/login.aspx?direct=true&AuthType=sso&db=lhh&AN=20033178190&site=ehost-live&scope=site |
| 2021 | Another dengue fever outbreak in eastern Ethiopia - an emerging public health threat | PLoS Neglected Tropical Diseases | hemorrhage | 10.1371/journal.pntd.0008992 | https://lstmed.idm.oclc.org/login?url=https://search.ebscohost.com/login.aspx?direct=true&AuthType=sso&db=lhh&AN=20210044438&site=ehost-live&scope=site |
| 2017 | Urban Chikungunya in the Middle East and North Africa: a systematic review | PLoS Neglected Tropical Diseases | Misr | https://journals.plos.org/plosntds/article?id=10.1371/journal.pntd.0005707 | https://lstmed.idm.oclc.org/login?url=https://search.ebscohost.com/login.aspx?direct=true&AuthType=sso&db=lhh&AN=20173329054&site=ehost-live&scope=site |
| 2018 | Neglected tropical diseases, neglected communities, and conflict: how do we leave no one behind? | Trends in Parasitology | animal-parasitic nematodes | 10.1016/j.pt.2017.10.013 | https://lstmed.idm.oclc.org/login?url=https://search.ebscohost.com/login.aspx?direct=true&AuthType=sso&db=lhh&AN=20183102951&site=ehost-live&scope=site |
| 2021 | Leishmaniasis beyond East Africa | Frontiers in Veterinary Science | parasitosis | 10.3389/fvets.2021.618766 | https://lstmed.idm.oclc.org/login?url=https://search.ebscohost.com/login.aspx?direct=true&AuthType=sso&db=lhh&AN=20210493634&site=ehost-live&scope=site |
| 2015 | Mapping and modelling the geographical distribution and environmental limits of podoconiosis in Ethiopia | PLoS Neglected Tropical Diseases | animal-parasitic nematodes | https://journals.plos.org/plosntds/article?id=10.1371/journal.pntd.0003946 | https://lstmed.idm.oclc.org/login?url=https://search.ebscohost.com/login.aspx?direct=true&AuthType=sso&db=lhh&AN=20153287031&site=ehost-live&scope=site |
| 2008 | Risk factors of visceral leishmaniasis in East Africa: a case-control study in Pokot territory of Kenya and Uganda | International Journal of Epidemiology | parasitosis | 10.1093/ije/dym275 | https://lstmed.idm.oclc.org/login?url=https://search.ebscohost.com/login.aspx?direct=true&AuthType=sso&db=lhh&AN=20083137624&site=ehost-live&scope=site |
| 2007 | Epidemiology and clinical features of patients with visceral leishmaniasis treated by an MSF clinic in Bakool region, Somalia, 2004-2006 | PLoS Neglected Tropical Diseases | parasitosis | 10.1371/journal.pntd.0000085 | https://lstmed.idm.oclc.org/login?url=https://search.ebscohost.com/login.aspx?direct=true&AuthType=sso&db=lhh&AN=20083057017&site=ehost-live&scope=site |
| 2013 | Spatially explicit Schistosoma infection risk in eastern Africa using Bayesian geostatistical modelling | Acta Tropica | bilharzia | 10.1016/j.actatropica.2011.10.006 | https://lstmed.idm.oclc.org/login?url=https://search.ebscohost.com/login.aspx?direct=true&AuthType=sso&db=lhh&AN=20133399131&site=ehost-live&scope=site |
| 2017 | Visceral leishmaniasis in Somalia: a review of epidemiology and access to care | PLoS Neglected Tropical Diseases | control programs | 10.1371/journal.pntd.0005231 | https://lstmed.idm.oclc.org/login?url=https://search.ebscohost.com/login.aspx?direct=true&AuthType=sso&db=lhh&AN=20173245738&site=ehost-live&scope=site |

#### Database: GREY LIT NTDS

Total records retrieved: 2

| Year | Title | Authors | URL |
| --- | --- | --- | --- |
| 2022 | Ending the neglect: eliminating worm infections as a public health problem in Somalia | WHO | https://reliefweb.int/report/somalia/ending-neglect-eliminating-worm-infections-public-health-problem-somalia |
| 2022 | Forgotten diseases: Achieving health equity to end the neglect of poverty-related diseases in Somalia | WHO | https://www.emro.who.int/somalia/news/forgotten-diseases-achieving-health-equity-to-end-the-neglect-of-poverty-related-diseases-in-somalia.html |

#### Database: medline complete NTDS

Total records retrieved: 26

| Year | Title | Journal | Authors | Keywords | DOI | URL |
| --- | --- | --- | --- | --- | --- | --- |
| 2024 | A Case Study on Unreported First Probable Human Rabies Following Honey Badger in Somalia | Open access emergency medicine : OAEM | Ali Osman, Ubah Mumin; Turfan, Selim; Farah Yusuf Mohamud, Mohamed | Somalia | 10.2147/OAEM.S439996 | https://lstmed.idm.oclc.org/login?url=https://search.ebscohost.com/login.aspx?direct=true&AuthType=sso&db=mdc&AN=38314068&site=ehost-live&scope=site |
| 2024 | Epidemiology of Echinococcus granulosus sensu lato in the Greater Horn of Africa: A systematic review | PLoS neglected tropical diseases | Aregawi, W. G.; Levecke, B.; Ashenafi, H.; Byaruhanga, C.; Kebede, N.; Mulinge, E.; Wassermann, M.; Romig, T.; Dorny, P.; Dermauw, V. | Echinococcus granulosus* | 10.1371/journal.pntd.0011894 | https://lstmed.idm.oclc.org/login?url=https://search.ebscohost.com/login.aspx?direct=true&AuthType=sso&db=mdc&AN=38271288&site=ehost-live&scope=site |
| 2018 | Visceral leishmaniasis in selected communities of Hamar and Banna-Tsamai districts in Lower Omo Valley, South West Ethiopia: Sero-epidemological and Leishmanin Skin Test Surveys | PloS one | Bekele, F.; Belay, T.; Zeynudin, A.; Hailu, A. | Residence Characteristics* | 10.1371/journal.pone.0197430 | https://lstmed.idm.oclc.org/login?url=https://search.ebscohost.com/login.aspx?direct=true&AuthType=sso&db=mdc&AN=29795589&site=ehost-live&scope=site |
| 2020 | Clinical characteristics and treatment of actinomycetoma in northeast Mexico: A case series | PLoS neglected tropical diseases | Cárdenas-de la Garza, J. A.; Welsh, O.; Cuéllar-Barboza, A.; Suarez-Sánchez, K. P.; De la Cruz-Valadez, E.; Cruz-Gómez, L. G.; Gallardo-Rocha, A.; Ocampo-Candiani, J.; Vera-Cabrera, L. | Anti-Bacterial Agents/*therapeutic use | 10.1371/journal.pntd.0008123 | https://lstmed.idm.oclc.org/login?url=https://search.ebscohost.com/login.aspx?direct=true&AuthType=sso&db=mdc&AN=32097417&site=ehost-live&scope=site |
| 2015 | Mapping and Modelling the Geographical Distribution and Environmental Limits of Podoconiosis in Ethiopia | PLoS neglected tropical diseases | Deribe, K.; Cano, J.; Newport, M. J.; Golding, N.; Pullan, R. L.; Sime, H.; Gebretsadik, A.; Assefa, A.; Kebede, A.; Hailu, A.; Rebollo, M. P.; Shafi, O.; Bockarie, M. J.; Aseffa, A.; Hay, S. I.; Reithinger, R.; Enquselassie, F.; Davey, G.; Brooker, S. J. | Elephantiasis/*epidemiology | 10.1371/journal.pntd.0003946 | https://lstmed.idm.oclc.org/login?url=https://search.ebscohost.com/login.aspx?direct=true&AuthType=sso&db=mdc&AN=26222887&site=ehost-live&scope=site |
| 2018 | Madura foot: an imported case of a non-common diagnosis | Le infezioni in medicina | Fasciana, T.; Colomba, C.; Cervo, A.; Di Carlo, P.; Scarlata, F.; Mascarella, C.; Giammanco, A.; Cascio, A. | Communicable Diseases, Imported*/diagnosis |  | https://lstmed.idm.oclc.org/login?url=https://search.ebscohost.com/login.aspx?direct=true&AuthType=sso&db=mdc&AN=29932092&site=ehost-live&scope=site |
| 2023 | Prevalence of trachoma in Somali region, Ethiopia: results from trachoma impact surveys in 50 woredas | International health | Gebreselassie, Getachew; Negash, Kasahun; Tsegaye, Sentayehu; Makonnen, Misrak; Deneke, Baye; Desalegn, Muluken; Harding-Esch, Emma M.; Harte, Anna; Solomon, Anthony W.; Boyd, Sarah; Bakhtiari, Ana; Hassen, Mussie Abdosh; Hambali, Abdulahi; Dejene, Michael; Beckwith, Colin; Tadesse, Fentahun; Sei... | Trachoma*/epidemiology | 10.1093/inthealth/ihad063 | https://lstmed.idm.oclc.org/login?url=https://search.ebscohost.com/login.aspx?direct=true&AuthType=sso&db=mdc&AN=38048381&site=ehost-live&scope=site |
| 2021 | Another dengue fever outbreak in Eastern Ethiopia-An emerging public health threat | PLoS neglected tropical diseases | Gutu, M. A.; Bekele, A.; Seid, Y.; Mohammed, Y.; Gemechu, F.; Woyessa, A. B.; Tayachew, A.; Dugasa, Y.; Gizachew, L.; Idosa, M.; Tokarz, R. E.; Sugerman, D. | Aedes/*classification | 10.1371/journal.pntd.0008992 | https://lstmed.idm.oclc.org/login?url=https://search.ebscohost.com/login.aspx?direct=true&AuthType=sso&db=mdc&AN=33465086&site=ehost-live&scope=site |
| 2023 | Tropical Data: Approach and Methodology as Applied to Trachoma Prevalence Surveys | Ophthalmic epidemiology | Harding-Esch, Emma M.; Burgert-Brucker, Clara R.; Jimenez, Cristina; Bakhtiari, Ana; Willis, Rebecca; Bejiga, Michael Dejene; Mpyet, Caleb; Ngondi, Jeremiah; Boyd, Sarah; Abdala, Mariamo; Abdou, Amza; Adamu, Yilikal; Alemayehu, Addisu; Alemayehu, Wondu; Al-Khatib, Tawfik; Apadinuwe, Sue-Chen; Awa... | Trachoma*/epidemiology | 10.1080/09286586.2023.2249546 | https://lstmed.idm.oclc.org/login?url=https://search.ebscohost.com/login.aspx?direct=true&AuthType=sso&db=mdc&AN=38085791&site=ehost-live&scope=site |
| 2023 | A Late Diagnosis of Visceral Leishmaniasis Using Tru-Cut Biopsy of the Spleen and Malaria Co-Infection - A Diagnostic Challenge: A Case Report in Somalia | Infection and drug resistance | Hassan, Mohamed Abdulahi; Omar, Abdullahi Abdirahman; Mohamed, Ibrahim Abdullahi; Garba, Bashiru; Fuje, Mohamed Mohamud Ali; Salad, Sagal Omar | co-infection | 10.2147/IDR.S420832 | https://lstmed.idm.oclc.org/login?url=https://search.ebscohost.com/login.aspx?direct=true&AuthType=sso&db=mdc&AN=37809037&site=ehost-live&scope=site |
| 2017 | Urban Chikungunya in the Middle East and North Africa: A systematic review | PLoS neglected tropical diseases | Humphrey, J. M.; Cleton, N. B.; Reusken, Cbem; Glesby, M. J.; Koopmans, M. P. G.; Abu-Raddad, L. J. | Urban Population* | 10.1371/journal.pntd.0005707 | https://lstmed.idm.oclc.org/login?url=https://search.ebscohost.com/login.aspx?direct=true&AuthType=sso&db=mdc&AN=28651007&site=ehost-live&scope=site |
| 2023 | Sero-prevalence of visceral leishmaniasis and its associated factors among asymptomatic individuals visiting Denan health center, southeastern Ethiopia | Tropical diseases, travel medicine and vaccines | Ismail, Ahmed; Yared, Solomon; Dugassa, Sisay; Abera, Adugna; Animut, Abebe; Erko, Berhanu; Gebresilassie, Araya | Denan | 10.1186/s40794-023-00196-8 | https://lstmed.idm.oclc.org/login?url=https://search.ebscohost.com/login.aspx?direct=true&AuthType=sso&db=mdc&AN=37430336&site=ehost-live&scope=site |
| 2008 | Risk factors of visceral leishmaniasis in East Africa: a case-control study in Pokot territory of Kenya and Uganda | International journal of epidemiology | Kolaczinski, J. H.; Reithinger, R.; Worku, D. T.; Ocheng, A.; Kasimiro, J.; Kabatereine, N.; Brooker, S. | Leishmania donovani* | 10.1093/ije/dym275 | https://lstmed.idm.oclc.org/login?url=https://search.ebscohost.com/login.aspx?direct=true&AuthType=sso&db=mdc&AN=18184669&site=ehost-live&scope=site |
| 2019 | Optimising age adjustment of trichiasis prevalence estimates using data from 162 standardised surveys from seven regions of Ethiopia | Ophthalmic epidemiology | Macleod, C. K.; Porco, T. C.; Dejene, M.; Shafi, O.; Kebede, B.; Negussu, N.; Bero, B.; Taju, S.; Adamu, Y.; Negash, K.; Haileselassie, T.; Riang, J.; Badei, A.; Bakhtiari, A.; Willis, R.; Bailey, R. L.; Solomon, A. W. | Population Surveillance/*methods | 10.1080/09286586.2018.1555262 | https://lstmed.idm.oclc.org/login?url=https://search.ebscohost.com/login.aspx?direct=true&AuthType=sso&db=mdc&AN=30592237&site=ehost-live&scope=site |
| 2003 | A neglected disease of humans: a new focus of visceral leishmaniasis in Bakool, Somalia | Transactions of the Royal Society of Tropical Medicine and Hygiene | Marlet, M. V. L.; Wuillaume, F.; Jacquet, D.; Quispe, K. W.; Dujardin, J. C.; Boelaert, M. | Leishmania donovani*/enzymology | 10.1016/s0035-9203(03)80099-8 | https://lstmed.idm.oclc.org/login?url=https://search.ebscohost.com/login.aspx?direct=true&AuthType=sso&db=mdc&AN=16117959&site=ehost-live&scope=site |
| 2013 | Prevalence and distribution of schistosomiasis in afder and gode zone of somali region, ethiopia | Journal of global infectious diseases | Negussu, Nebiyu; Wali, Mohamed; Ejigu, Milion; Debebe, Fikiru; Aden, Sirage; Abdi, Rashid; Mohamed, Yusuf; Deribew, Amare; Deribe, Kebede | S. haematobium | 10.4103/0974-777X.122007 | https://lstmed.idm.oclc.org/login?url=https://search.ebscohost.com/login.aspx?direct=true&AuthType=sso&db=mdc&AN=24672176&site=ehost-live&scope=site |
| 2020 | Prevalence and pattern of waterborne parasitic infections in eastern Africa: A systematic scoping review | Food and waterborne parasitology | Ngowi, Helena A. | Burden | 10.1016/j.fawpar.2020.e00089 | https://lstmed.idm.oclc.org/login?url=https://search.ebscohost.com/login.aspx?direct=true&AuthType=sso&db=mdc&AN=32995583&site=ehost-live&scope=site |
| 2007 | Epidemiology and clinical features of patients with visceral leishmaniasis treated by an MSF clinic in Bakool region, Somalia, 2004-2006 | PLoS neglected tropical diseases | Raguenaud, Marie-Eve; Jansson, Anna; Vanlerberghe, Veerle; Deborggraeve, Stijn; Dujardin, Jean-Claude; Orfanos, Giannos; Reid, Tony; Boelaert, Marleen | Leishmaniasis, Visceral/*therapy | 10.1371/journal.pntd.0000085 | https://lstmed.idm.oclc.org/login?url=https://search.ebscohost.com/login.aspx?direct=true&AuthType=sso&db=mdc&AN=17989791&site=ehost-live&scope=site |
| 2021 | Correction: Epidemiology and Clinical Features of Patients with Visceral Leishmaniasis Treated by an MSF Clinic in Bakool Region, Somalia, 2004-2006 | PLoS neglected tropical diseases | Raguenaud, M. E.; Jansson, A.; Vanlerberghe, V.; Van der Auwera, G.; Deborggraeve, S.; Dujardin, J. C.; Orfanos, I.; Reid, T.; Boelaert, M. |  | 10.1371/journal.pntd.0009356 | https://lstmed.idm.oclc.org/login?url=https://search.ebscohost.com/login.aspx?direct=true&AuthType=sso&db=mdc&AN=33872317&site=ehost-live&scope=site |
| 2020 | Progress towards elimination of lymphatic filariasis in the Eastern Mediterranean Region | International health | Ramzy, Reda M. R.; Al Kubati, Abdul Samid | Elephantiasis, Filarial*/drug therapy | 10.1093/inthealth/ihaa037 | https://lstmed.idm.oclc.org/login?url=https://search.ebscohost.com/login.aspx?direct=true&AuthType=sso&db=mdc&AN=33349874&site=ehost-live&scope=site |
| 2013 | Spatially explicit Schistosoma infection risk in eastern Africa using Bayesian geostatistical modelling | Acta tropica | Schur, Nadine; Hürlimann, Eveline; Stensgaard, Anna-Sofie; Chimfwembe, Kingford; Mushinge, Gabriel; Simoonga, Christopher; Kabatereine, Narcis B.; Kristensen, Thomas K.; Utzinger, Jürg; Vounatsou, Penelope | Topography, Medical* | 10.1016/j.actatropica.2011.10.006 | https://lstmed.idm.oclc.org/login?url=https://search.ebscohost.com/login.aspx?direct=true&AuthType=sso&db=mdc&AN=22019933&site=ehost-live&scope=site |
| 2023 | Gender differences in the surgical management of trachomatous trichiasis: an exploratory analysis of global trachoma survey data, 2015-2019 | International health | Sullivan, Kristin M.; Harding-Esch, Emma M.; Batcho, Wilfrid E.; Issifou, Amadou A. Bio; Lopes, Maria de Fátima Costa; Szwarcwald, Celia Landmann; Vaz Ferreira Gomez, Daniela; Bougouma, Clarisse; Christophe, Nassa; Kabore, Martin; Bucumi, Victor; Bella, Assumpta L.; Epee, Emilienne; Yaya, Georges... | Trichiasis*/epidemiology | 10.1093/inthealth/ihad067 | https://lstmed.idm.oclc.org/login?url=https://search.ebscohost.com/login.aspx?direct=true&AuthType=sso&db=mdc&AN=38048383&site=ehost-live&scope=site |
| 2017 | Visceral leishmaniasis in Somalia: A review of epidemiology and access to care | PLoS neglected tropical diseases | Sunyoto, T.; Potet, J.; Boelaert, M. | Health Services Accessibility* | 10.1371/journal.pntd.0005231 | https://lstmed.idm.oclc.org/login?url=https://search.ebscohost.com/login.aspx?direct=true&AuthType=sso&db=mdc&AN=28278151&site=ehost-live&scope=site |
| 2019 | Exploring global and country-level barriers to an effective supply of leishmaniasis medicines and diagnostics in eastern Africa: a qualitative study | BMJ open | Sunyoto, Temmy; Potet, Julien; den Boer, Margriet; Ritmeijer, Koert; Postigo, Jose A. R.; Ravinetto, Raffaella; Alves, Fabiana; Picado, Albert; Boelaert, Marleen | Agglutination Tests/*statistics & numerical data | 10.1136/bmjopen-2019-029141 | https://lstmed.idm.oclc.org/login?url=https://search.ebscohost.com/login.aspx?direct=true&AuthType=sso&db=mdc&AN=31152044&site=ehost-live&scope=site |
| 2022 | Risk factors for Brucellosis and knowledge-attitude practice among pastoralists in Afar and Somali regions of Ethiopia | Preventive veterinary medicine | Tschopp, Rea; GebreGiorgis, Ashenafi; Abdulkadir, Oumer; Molla, Wassie; Hamid, Muhammed; Tassachew, Yayehyirad; Andualem, Henok; Osman, Mahlet; Waqjira, Mulugeta Waji; Mohammed, Abdulkadir; Negron, Maria; Walke, Henry; Kadzik, Melissa; Mamo, Gezahegne | Brucellosis*/epidemiology | 10.1016/j.prevetmed.2021.105557 | https://lstmed.idm.oclc.org/login?url=https://search.ebscohost.com/login.aspx?direct=true&AuthType=sso&db=mdc&AN=34902652&site=ehost-live&scope=site |
| 2021 | Integrated human-animal sero-surveillance of Brucellosis in the pastoral Afar and Somali regions of Ethiopia | PLoS neglected tropical diseases | Tschopp, R.; Gebregiorgis, A.; Tassachew, Y.; Andualem, H.; Osman, M.; Waqjira, M. W.; Hattendorf, J.; Mohammed, A.; Hamid, M.; Molla, W.; Mitiku, S. A.; Walke, H.; Negron, M.; Kadzik, M.; Mamo, G. | Livestock* | 10.1371/journal.pntd.0009593 | https://lstmed.idm.oclc.org/login?url=https://search.ebscohost.com/login.aspx?direct=true&AuthType=sso&db=mdc&AN=34358232&site=ehost-live&scope=site |

#### Database: WEB OF SCIENCE NTDS

Total records retrieved: 104

| Year | Title | Journal | Authors | Keywords | DOI | URL |
| --- | --- | --- | --- | --- | --- | --- |
| 2020 | Visceral Leishmaniasis, Northern Somalia, 2013-2019 | Emerging Infectious Diseases | Aalto, M. K.; Sunyoto, T.; Yusuf, M. A. A.; Mohamed, A. A.; Van der Auwera, G.; Dujardin, J. C. | Immunology | 10.3201/eid2601.181851 | <Go to ISI>://WOS:000505702700024 |
| 2021 | Preliminary findings of COVID-19 infection in health workers in Somalia: A reason for concern | International Journal of Infectious Diseases | Abdi, A.; Ahmed, A. Y.; Abdulmunim, M.; Karanja, M. J.; Solomon, A.; Muhammad, F.; Kumlachew, M.; Obtel, M.; Malik, Smmr | Somalia | 10.1016/j.ijid.2021.01.066 | <Go to ISI>://WOS:000632935600018 |
| 1991 | HIGH PREVALENCE OF ANTI-HEPATITIS DELTA VIRUS-ANTIBODY IN CHRONIC LIVER-DISEASE IN SOMALIA | Transactions of the Royal Society of Tropical Medicine and Hygiene | Aceti, A.; Mohamed, O. M.; Paparo, B. S.; Mohamud, O. M.; Quaranta, G.; Maalin, K. A.; Sebastiani, A. | b virus | 10.1016/0035-9203(91)90249-x | <Go to ISI>://WOS:A1991GD47500033 |
| 1989 | SERO-EPIDEMIOLOGY OF HEPATITIS DELTA VIRUS-INFECTION IN SOMALIA | Transactions of the Royal Society of Tropical Medicine and Hygiene | Aceti, A.; Paparo, B. S.; Celestino, D.; Pennica, A.; Caferro, M.; Grilli, A.; Sebastiani, A.; Mohamud, O. M.; Abdirahman, M.; Bile, K. | Public, Environmental & Occupational Health | 10.1016/0035-9203(89)90516-6 | <Go to ISI>://WOS:A1989AE11200034 |
| 1993 | HEPATITIS-C VIRUS-INFECTION IN CHRONIC LIVER-DISEASE IN SOMALIA | American Journal of Tropical Medicine and Hygiene | Aceti, A.; Taliani, G.; Bruni, R.; Sharif, O. S.; Moallin, K. A.; Celestino, D.; Quaranta, G.; Sebastiani, A. | anti-hcv antibodies | 10.4269/ajtmh.1993.48.581 | <Go to ISI>://WOS:A1993LB69200017 |
| 1990 | ANTIBODIES REACTIVE WITH NON-POLYMORPHIC EPITOPES ON HLA MOLECULES CAUSE FALSE-POSITIVE HIV ANTIBODY TESTS ON AFRICAN SAMPLES OF SERUM | Journal of Infection | Aceti, A.; Terzaroli, P.; Paparo, B. S.; Celestino, D.; Sebastiani, A.; Bile, K.; Tanigaki, N.; Tosi, R.; Ameglio, F. | Infectious Diseases | 10.1016/s0163-4453(90)92478-4 | <Go to ISI>://WOS:A1990CL13500014 |
| 2022 | Epidemiology of Multidrug Resistant Non-Fermentative Gram Negative Bacilli in Patients with Hospital Acquired Pneumonia: An Alarming Report from Somalia | Infection and Drug Resistance | Adan, F. N.; Jeele, M. O. O.; Omar, N. M. S. | hospital acquired pneumonia | 10.2147/idr.S387370 | <Go to ISI>://WOS:000880435700001 |
| 2023 | Improved leprosy elimination efforts in Somalia, 2015-2021: achievements in a challenging environment and the way forward | Leprosy Review | Aden, A.; Ali, A. A.; Amran, J.; Osman, M.; Warusavithana, S.; Pemmaraju, V. R.; Atta, H.; Hutin, Y. | Leprosy | 10.47276/lr.94.2.98 | <Go to ISI>://WOS:001024927900002 |
| 1992 | AN EPIDEMIC OF NEISSERIA-GONORRHOEAE IN A SOMALI ORPHANAGE | International Journal of Std & Aids | Ahmed, H. J.; Ilardi, I.; Antognoli, A.; Leone, F.; Sebastiani, A.; Amiconi, G. | neisseria-gonorrhoeae | 10.1177/095646249200300113 | <Go to ISI>://WOS:A1992HF45500013 |
| 1988 | HUMAN TOXOPLASMOSIS IN SOMALIA - PREVALENCE OF TOXOPLASMA ANTIBODIES IN A VILLAGE IN THE LOWER SCEBELLI REGION AND IN MOGADISHU | Transactions of the Royal Society of Tropical Medicine and Hygiene | Ahmed, H. J.; Mohammed, H. H.; Yusuf, M. W.; Ahmed, S. F.; Huldt, G. | Public, Environmental & Occupational Health | 10.1016/0035-9203(88)90465-8 | <Go to ISI>://WOS:A1988N315400053 |
| 1992 | ISOLATION OF DRUG-RESISTANT STRAINS OF TRYPANOSOMA-CONGOLENSE FROM THE LOWER SHABELLE REGION OF SOUTHERN SOMALIA | Tropical Animal Health and Production | Ainanshe, O. A.; Jennings, F. W.; Holmes, P. H. | Agriculture | 10.1007/bf02356946 | <Go to ISI>://WOS:A1992JB21300001 |
| 2023 | Public health impact of the spread of <i>Anopheles stephensi</i> in the WHO Eastern Mediterranean Region countries in Horn of Africa and Yemen: need for integrated vector surveillance and control | Malaria Journal | Al-Eryani, S. M.; Irish, S. R.; Carter, T. E.; Lenhart, A.; Aljasari, A.; Montoya, L. F.; Awash, A. A.; Mohammed, E.; Ali, S.; Esmail, M. A.; Hussain, A.; Amran, J. G.; Kayad, S.; Nouredayem, M.; Adam, M. A.; Azkoul, L.; Assada, M.; Baheshm, Y. A.; Eltahir, W.; Hutin, Y. J. | Anopheles stephensi | 10.1186/s12936-023-04545-y | <Go to ISI>://WOS:001010787900001 |
| 2022 | Survival analysis of all critically ill patients with COVID-19 admitted to the main hospital in Mogadishu, Somalia, 30 March-12 June 2020: which interventions are proving effective in fragile states? | International Journal of Infectious Diseases | Ali, M. M.; Malik, M. R.; Ahmed, A. Y.; Bashir, A. M.; Mohamed, A.; Abdi, A.; Obtel, M. | COVID-19 | 10.1016/j.ijid.2021.11.018 | <Go to ISI>://WOS:000807520900036 |
| 2022 | Morphological identification and genetic characterization of <i>Anopheles stephensi</i> in Somaliland | Parasites & Vectors | Ali, S.; Samake, J. N.; Spear, J.; Carter, T. E. | Malaria | 10.1186/s13071-022-05339-y | <Go to ISI>://WOS:000825729900002 |
| 1990 | INVITRO ACTIVITY OF ARTEMISININ, ITS DERIVATIVES, AND PYRONARIDINE AGAINST DIFFERENT STRAINS OF PLASMODIUM-FALCIPARUM | Transactions of the Royal Society of Tropical Medicine and Hygiene | Alin, M. H.; Bjorkman, A.; Ashton, M. | Public, Environmental & Occupational Health | 10.1016/0035-9203(90)90129-3 | <Go to ISI>://WOS:A1990EG35200007 |
| 2021 | Positive Effect of Single-Dose Measles Vaccination in Reducing the Incidence of Pneumonia in Children with Measles | Journal of Tropical Pediatrics | Bagci, Z.; Daki, Y. Y. | measles | 10.1093/tropej/fmaa085 | <Go to ISI>://WOS:000637542600017 |
| 2022 | Prevalence of Acute Kidney Injury in Covid-19 Patients- Retrospective Single-Center Study | Infection and Drug Resistance | Bashir, A. M.; Mukhtar, M. S.; Mohamed, Y. G.; Cetinkaya, O.; Fiidow, O. A. | Somalia | 10.2147/idr.S357997 | <Go to ISI>://WOS:000783812900011 |
| 2019 | ONE HEALTH REGIONAL NETWORK FOR THE HORN OF AFRICA | Transactions of the Royal Society of Tropical Medicine and Hygiene | Baylis, M.; Asrat, D.; Fevre, F.; Kahsay, M.; Mor, S.; Ongore, D.; Pulford, J.; Wesonga, F. | Public, Environmental & Occupational Health |  | <Go to ISI>://WOS:000493064400344 |
| 1991 | LATE SEROCONVERSION TO HEPATITIS-B IN A SOMALI VILLAGE INDICATES THE IMPORTANT ROLE OF VENEREAL TRANSMISSION | Journal of Tropical Medicine and Hygiene | Bile, K.; Abdirahman, M.; Mohamud, O.; Aden, C.; Isse, A.; Nilsson, L.; Norder, H.; Magnius, L. | virus-infection |  | <Go to ISI>://WOS:A1991GV94500001 |
| 1993 | IMPORTANT ROLE OF HEPATITIS-C VIRUS-INFECTION AS A CAUSE OF CHRONIC LIVER-DISEASE IN SOMALIA | Scandinavian Journal of Infectious Diseases | Bile, K.; Aden, C.; Norder, H.; Magnius, L.; Lindberg, G.; Nilsson, L. | non-b-hepatitis | 10.3109/00365549309008543 | <Go to ISI>://WOS:A1993MG84500002 |
| 1994 | CONTRASTING ROLES OF RIVERS AND WELLS AS SOURCES OF DRINKING-WATER ON ATTACK AND FATALITY RATES IN A HEPATITIS-E EPIDEMIC IN SOMALIA | American Journal of Tropical Medicine and Hygiene | Bile, K.; Isse, A.; Mohamud, O.; Allebeck, P.; Nilsson, L.; Norder, H.; Mushahwar, I. K.; Magnius, L. O. | non-b-hepatitis | 10.4269/ajtmh.1994.51.466 | <Go to ISI>://WOS:A1994PN55300015 |
| 2014 | Dengue fever outbreak in Mogadishu, Somalia 2011: Co-circulation of three dengue virus serotypes | International Journal of Infectious Diseases | Bosa, H. K.; Montgomery, J. M.; Kimuli, I.; Lutwama, J. J. | Infectious Diseases | 10.1016/j.ijid.2014.03.412 | <Go to ISI>://WOS:000209704000006 |
| 1989 | SEROLOGICAL EVIDENCE OF DENGUE FEVER AMONG REFUGEES, HARGEYSA, SOMALIA | Journal of Medical Virology | Botros, B. A. M.; Watts, D. M.; Soliman, A. K.; Salib, A. W.; Moussa, M. I.; Mursal, H.; Douglas, C.; Farah, M. | Virology | 10.1002/jmv.1890290202 | <Go to ISI>://WOS:A1989AY86400001 |
| 2015 | Use of a bibliometric literature review to assess medical research capacity in post-conflict and developing countries: Somaliland 1991-2013 | Tropical Medicine & International Health | Boyce, R.; Rosch, R.; Finlayson, A.; Handuleh, D.; Walhad, S. A.; Whitwell, S.; Leather, A. | research | 10.1111/tmi.12590 | <Go to ISI>://WOS:000362583100013 |
| 1990 | COMPARATIVE PHARMACOKINETICS OF AMIKACIN SULFATE IN CALVES AND SHEEP | Research in Veterinary Science | Carli, S.; Montesissa, C.; Sonzogni, O.; Madonna, M.; Saidfaqi, A. | Veterinary Sciences | 10.1016/s0034-5288(18)30996-2 | <Go to ISI>://WOS:A1990CV76900018 |
| 1988 | A 2-YEAR STUDY OF ENTERIC INFECTIONS ASSOCIATED WITH DIARRHEAL DISEASES IN CHILDREN IN URBAN SOMALIA | Transactions of the Royal Society of Tropical Medicine and Hygiene | Casalino, M.; Yusuf, M. W.; Nicoletti, M.; Bazzicalupo, P.; Coppo, A.; Colonna, B.; Cappelli, C.; Bianchini, C.; Falbo, V.; Ahmed, H. J.; Omar, K. H.; Maxamuud, K. B.; Maimone, F. | Public, Environmental & Occupational Health | 10.1016/0035-9203(88)90542-1 | <Go to ISI>://WOS:A1988Q036200047 |
| 1995 | VIBRIO-CHOLERAE IN THE HORN-OF-AFRICA - EPIDEMIOLOGY, PLASMIDS, TETRACYCLINE RESISTANCE GENE AMPLIFICATION, AND COMPARISON BETWEEN O1 AND NON-O1 STRAINS | American Journal of Tropical Medicine and Hygiene | Coppo, A.; Colombo, M.; Pazzani, C.; Bruni, R.; Mohamud, K. A.; Omar, K. H.; Mastrandrea, S.; Salvia, A. M.; Rotigliano, G.; Maimone, F. | states gulf-coast | 10.4269/ajtmh.1995.53.351 | <Go to ISI>://WOS:A1995TD72700006 |
| 1986 | SOMALIA - ENDOCRINOLOGIC ASPECTS OF REPRODUCTION IN THE FEMALE CAMEL | World Animal Review | Cristofori, P.; Aria, G.; Seren, E.; Bono, G.; Aaden, A. S.; Nur, H. M. | Agriculture |  | <Go to ISI>://WOS:A1986A720500004 |
| 2022 | Prevalence of Multidrug-Resistant TB Among Smear-Positive Pulmonary TB Patients in Banadir, Somalia: A Multicenter Study | Infection and Drug Resistance | Dirie, A. M. H.; Çolakoglu, S.; Abdulle, O. M.; Abdi, B. M.; Osman, M. A.; Shire, A. M.; Hussein, A. M. | TB | 10.2147/idr.S386497 | <Go to ISI>://WOS:000894699700001 |
| 1989 | CAMEL TRYPANOSOMIASIS AND ITS VECTORS IN SOMALIA | Veterinary Parasitology | Dirie, M. F.; Wallbanks, K. R.; Aden, A. A.; Bornstein, S.; Ibrahim, M. D. | Parasitology | 10.1016/0304-4017(89)90039-3 | <Go to ISI>://WOS:A1989AL38700003 |
| 2024 | Mycetoma case series in Somalia | Tropical Doctor | Dogan, A.; Ali, A. M.; Ali, M. A.; Abdullahi, I. M. | Actinomycetoma | 10.1177/00494755231201664 | <Go to ISI>://WOS:001067709100001 |
| 2023 | Prevalence of Post COVID-19 Condition among Healthcare Workers: Self-Reported Online Survey in Four African Countries, December 2021-January 2022 | Covid | Elnadi, H.; Al-Mustapha, A. I.; Odetokun, I. A.; Anjorin, A. A.; Mosbah, R.; Fasina, F. O.; Razouqi, Y.; Awiagah, K. S.; Nyandwi, J. B.; Mhgoob, Z. E.; Gachara, G.; Mohamud, M. F. Y.; Damaris, B. F.; Maisara, A. M. O.; Radwan, M. | PCC | 10.3390/covid3110114 | <Go to ISI>://WOS:001182208600001 |
| 1988 | A FILTER-PAPER TECHNIQUE FOR THE DETECTION OF IGG AND IGM CLASS SCHISTOSOME-SPECIFIC ANTIBODIES IN AN ENDEMIC AREA | Annals of Tropical Medicine and Parasitology | Evengard, B.; Hagi, H.; Linder, E. | Public, Environmental & Occupational Health | 10.1080/00034983.1988.11812248 | <Go to ISI>://WOS:A1988P034200013 |
| 2022 | Hemoplasmas and ticks in cattle from Somalia | Acta Tropica | Ferrari, L. D. R.; Hassan-Kadle, A. A.; Collere, F. C. M.; Coradi, V. S.; Ibrahim, A. M.; Osman, A. M.; Shair, M. A.; André, M. R.; Vieira, Tswj; Machado, R. Z.; Vieira, R. F. C. | Hemotropic mycoplasmas | 10.1016/j.actatropica.2022.106696 | <Go to ISI>://WOS:000866460500001 |
| 2004 | Mapping the potential distribution of <i>Phlebotomus martini</i> and <i>P-orientalis</i> (Diptera: Psychodidae), vectors of kala-azar in East Africa by use of geographic information systems | Acta Tropica | Gebre-Michael, T.; Malone, J. B.; Balkew, M.; Ali, A.; Berhe, N.; Hailu, A.; Herzi, A. A. | kala-azar | 10.1016/j.actatropica.2003.09.021 | <Go to ISI>://WOS:000188776600011 |
| 2023 | Increasing prevalence of malaria and acute dengue virus coinfection in Africa: a meta-analysis and meta-regression of cross-sectional studies | Malaria Journal | Gebremariam, T. T.; Schalling, Hdfh; Kurmane, Z. M.; Danquah, J. B. | Prevalence | 10.1186/s12936-023-04723-y | <Go to ISI>://WOS:001079379500001 |
| 2009 | Seroprevalence of camel brucellosis (Camelus dromedarius) in Somaliland | Tropical Animal Health and Production | Ghanem, Y. M.; El-Khodery, S. A.; Saad, A. A.; Abdelkader, A. H.; Heybe, A.; Musse, Y. A. | Camel brucellosis | 10.1007/s11250-009-9377-9 | <Go to ISI>://WOS:000271421200022 |
| 2018 | Using non-exceedance probabilities of policy-relevant malaria prevalence thresholds to identify areas of low transmission in Somalia | Malaria Journal | Giorgi, E.; Osman, A. A.; Hassan, A. H.; Ali, A. A.; Ibrahim, F.; Amran, J. G. H.; Noor, A. M.; Snow, R. W. | plasmodium-falciparum | 10.1186/s12936-018-2238-0 | <Go to ISI>://WOS:000425536000006 |
| 2023 | Minimal Change Nephrotic Syndrome with Acute Kidney Injury after the Administration of Pfizer-BioNTech COVID-19 Vaccine | Case Reports in Infectious Diseases | Gulumsek, E.; Ozturk, D. D.; Ozturk, H. A.; Saler, T.; Erdogan, K. E.; Bashir, A. M.; Sumbul, H. E. | change disease | 10.1155/2023/5122228 | <Go to ISI>://WOS:000943639400001 |
| 1990 | ANTIBODY-RESPONSES IN SCHISTOSOMIASIS-HAEMATOBIA IN SOMALIA - RELATION TO AGE AND INFECTION INTENSITY | Annals of Tropical Medicine and Parasitology | Hagi, H.; Huldt, G.; Loftenius, A.; Schroder, H. | Public, Environmental & Occupational Health | 10.1080/00034983.1990.11812451 | <Go to ISI>://WOS:A1990DB56100009 |
| 1989 | SILICATE PNEUMOCONIOSIS IN CAMELS (CAMELUS-DROMEDARIUS L) | Journal of Veterinary Medicine Series a-Physiology Pathology Clinical Medicine | Hansen, H. J.; Jama, F. M.; Nilsson, C.; Norrgren, L.; Abdurahman, O. S. | Veterinary Sciences | 10.1111/j.1439-0442.1989.tb00793.x | <Go to ISI>://WOS:A1989CJ03200009 |
| 2023 | A Late Diagnosis of Visceral Leishmaniasis Using Tru-Cut Biopsy of the Spleen and Malaria Co-Infection - A Diagnostic Challenge: A Case Report in Somalia | Infection and Drug Resistance | Hassan, M. A.; Omar, A. A.; Mohamed, I. A.; Garba, B.; Fuje, M. M. A.; Salad, S. O. | visceral leishmaniasis | 10.2147/idr.S420832 | <Go to ISI>://WOS:001078624900001 |
| 2022 | Prevalence and Species Identification of Ixodid Ticks of Small Ruminants in Benadir Region, Somalia | Veterinary Medicine International | Hassan, Y. H. S.; Jimale, K. A.; Dirie, S. S.; Salah, O. M.; Afrah, O. H.; Mohamed, M. O. S.; Dubad, A. B. | Veterinary Sciences | 10.1155/2022/9224737 | <Go to ISI>://WOS:000993224000001 |
| 2019 | Parasitological, serological and molecular survey of camel trypanosomiasis in Somalia | Parasites & Vectors | Hassan-Kadle, A. A.; Ibrahim, A. M.; Nyingilili, H. S.; Yusuf, A. A.; Vieira, Tswj; Vieira, R. F. C. | CATT/T | 10.1186/s13071-019-3853-5 | <Go to ISI>://WOS:000503802300002 |
| 2024 | One Health in Somalia: Present status, opportunities, and challenges | One Health | Hassan-Kadle, A. A.; Osman, A. M.; Ibrahim, A. M.; Mohamed, A. A.; de Oliveira, C. J. B.; Vieira, R. F. C. | One Health | 10.1016/j.onehlt.2023.100666 | <Go to ISI>://WOS:001146491500001 |
| 2021 | Rift Valley fever and <i>Brucella</i> spp. in ruminants, Somalia | Bmc Veterinary Research | Hassan-Kadle, A. A.; Osman, A. M.; Shair, M. A.; Abdi, O. M.; Yusuf, A. A.; Ibrahim, A. M.; Vieira, R. F. C. | Neglected zoonotic diseases | 10.1186/s12917-021-02980-0 | <Go to ISI>://WOS:000687184800001 |
| 2023 | Population-based sero-epidemiological investigation of SARS-CoV-2 infection in Somalia | Journal of Infection and Public Health | Hossain, M. S.; Derrow, M. M.; Mohamed, S. I.; Abukar, H. M.; Qayad, M. G.; Malik, Smmr; Mengistu, K. F.; Obsie, A. A. A.; Anwar, I. | COVID-19 | 10.1016/j.jiph.2023.04.016 | <Go to ISI>://WOS:000988525200001 |
| 1987 | EPIDEMIOLOGIC-STUDY OF PARASITIC INFECTIONS IN SOMALI NOMADS | Transactions of the Royal Society of Tropical Medicine and Hygiene | Ilardi, I.; Sebastiani, A.; Leone, F.; Madera, A.; Bile, M. K.; Shiddo, S. C.; Mohamed, H. H.; Amiconi, G. | Public, Environmental & Occupational Health | 10.1016/0035-9203(87)90027-7 | <Go to ISI>://WOS:A1987K878000024 |
| 2022 | Community Based Surveillance in Somaliland: Analysis of the Functionality and Effectiveness using the CBS Platform Nyss | International Journal of Infectious Diseases | Jung, J.; Beledi, A. H.; Riedel, N.; Ahmed, A. O.; Larsen, T. M. | Infectious Diseases | 10.1016/j.ijid.2021.12.236 | <Go to ISI>://WOS:000775894100236 |
| 2011 | Measles Control and Elimination in Somalia: The Good, the Bad, and the Ugly | Journal of Infectious Diseases | Kamadjeu, R.; Assegid, K.; Naouri, B.; Mirza, I. R.; Hirsi, A.; Mohammed, A.; Omer, M.; Dualle, A. H.; Mulugeta, A. | eastern mediterranean region | 10.1093/infdis/jir066 | <Go to ISI>://WOS:000293547600040 |
| 2024 | Diversity of Glossinidae (Diptera) species in The Gambia in relation to vegetation | Revista Brasileira De Parasitologia Veterinaria | Kargbo, A.; Jallow, M.; Vieira, Tswj; Amoutchi, A. I.; Koua, H. K.; Osman, A. M.; Vieira, R. F. D. | Glossina morsitans submorsitans | 10.1590/s1984-29612024010 | <Go to ISI>://WOS:001169842200001 |
| 2021 | Clinical characteristics of acute liver failure associated with hepatitis A infection in children in Mogadishu, Somalia: a hospital-based retrospective study | Bmc Infectious Diseases | Keles, E.; Hassan-Kadle, M. A.; Osman, M. M.; Eker, H. H.; Abusoglu, Z.; Baydili, K. N.; Osman, A. M. | Acute liver failure | 10.1186/s12879-021-06594-7 | <Go to ISI>://WOS:000693837600002 |
| 1981 | PREVALENCE OF SCHISTOSOMA-HAEMATOBIUM IN THE KORYOLE AND MERCA DISTRICTS OF THE SOMALI-DEMOCRATIC-REPUBLIC | Annals of Tropical Medicine and Parasitology | Koura, M.; Upatham, E. S.; Awad, A. H.; Ahmed, M. D. | Public, Environmental & Occupational Health | 10.1080/00034983.1981.11687408 | <Go to ISI>://WOS:A1981LC43600008 |
| 2010 | HIV prevalence and characteristics of sex work among female sex workers in Hargeisa, Somaliland, Somalia | Aids | Kriitmaa, K.; Testa, A.; Osman, M.; Bozicevic, I.; Riedner, G.; Malungu, J.; Irving, G.; Abdalla, I. | biological and behavioural surveillance | 10.1097/01.aids.0000386735.87177.2a | <Go to ISI>://WOS:000279697100008 |
| 1992 | EFFECT OF DIFFERENT FRACTIONS OF HEPARIN ON PLASMODIUM-FALCIPARUM MEROZOITE INVASION OF RED-BLOOD-CELLS INVITRO | American Journal of Tropical Medicine and Hygiene | Kulane, A.; Ekre, H. P.; Perlmann, P.; Rombo, L.; Wahlgren, M.; Wahlin, B. | spontaneous erythrocyte rosettes | 10.4269/ajtmh.1992.46.589 | <Go to ISI>://WOS:A1992HX46300013 |
| 2018 | Detection and molecular characterization of tick-borne pathogens infecting sheep and goats in Blue Nile and West Kordofan states in Sudan | Ticks and Tick-Borne Diseases | Lee, S. H.; Mossaad, E.; Ibrahim, A. M.; Ismail, A. A.; Moumouni, P. F. A.; Liu, M. M.; Ringo, A. E.; Gao, Y.; Guo, H. P.; Li, J. X.; Efstratiou, A.; Musinguzi, P.; Angara, T. E. E.; Suganuma, K.; Inoue, N.; Xuan, X. N. | Anaplasma ovis | 10.1016/j.ttbdis.2018.01.014 | <Go to ISI>://WOS:000433128600021 |
| 1986 | CLONAL SPREAD OF MULTIPLY RESISTANT STRAINS OF VIBRIO-CHOLERAE O1 IN SOMALIA | Journal of Infectious Diseases | Maimone, F.; Coppo, A.; Pazzani, C.; Ismail, S. O.; Guerra, R.; Procacci, P.; Rotigliano, G.; Omar, K. H. | Immunology | 10.1093/infdis/153.4.802 | <Go to ISI>://WOS:A1986A498100029 |
| 1988 | OUTBREAK OF CAMEL CONTAGIOUS ECTHYMA IN CENTRAL SOMALIA | Tropical Animal Health and Production | Moallin, A. S. M.; Zessin, K. H. | Agriculture | 10.1007/bf02240091 | <Go to ISI>://WOS:A1988Q403200013 |
| 2022 | Antimicrobial Resistance and Predisposing Factors Associated with Catheter-Associated UTI Caused by Uropathogens Exhibiting Multidrug-Resistant Patterns: A 3-Year Retrospective Study at a Tertiary Hospital in Mogadishu, Somalia | Tropical Medicine and Infectious Disease | Mohamed, A. H.; Omar, N. M. S.; Osman, M. M.; Mohamud, H. A.; Eraslan, A.; Gur, M. | catheter-associated UTI | 10.3390/tropicalmed7030042 | <Go to ISI>://WOS:000774574300001 |
| 2024 | Epidemiological investigation of dengue fever outbreak and its socioeconomic determinants in Banadir region, Somalia | Bmc Infectious Diseases | Mohamed, M. A.; Hassan, N. Y.; Osman, M. M.; Gedi, S.; Maalin, B. A. A.; Sultan, K. M.; Garba, B.; Osman, A. A.; Osman, A. Y.; Ahmed, A. D. | Dengue fever | 10.1186/s12879-024-09276-2 | <Go to ISI>://WOS:001220488800002 |
| 1979 | C-DROMEDARIUS ERYTHROCYTES - INVITRO RESISTENCE TO HEMOLYSIS | Archivio Veterinario Italiano | Mohamed, M. H.; Locatelli, A.; Macchioni, G. | Veterinary Sciences |  | <Go to ISI>://WOS:A1979HR34200012 |
| 2023 | Determinants of HIV/Aids Knowledge Among Females in Somalia: Findings from 2018 to 2019 SDHS Data | Hiv Aids-Research and Palliative Care | Mohamud, L. A.; Hassan, A. M.; Nasir, J. A. | knowledge of HIV | 10.2147/hiv.S414290 | <Go to ISI>://WOS:001038306400001 |
| 2020 | Loss of Taste and Smell are Common Clinical Characteristics of Patients with COVID-19 in Somalia: A Retrospective Double Centre Study | Infection and Drug Resistance | Mohamud, M. F. Y.; Mohamed, Y. G.; Ali, A. M.; Adam, B. A. | anosmia | 10.2147/idr.S263632 | <Go to ISI>://WOS:000571494000001 |
| 2020 | A cost effectiveness analysis of active case finding and passive case finding for case detection of TB at Kisugu Health Center III, Kampala, Uganda | International Journal of Infectious Diseases | Muhoozi, M.; Tusabe, J.; Tabwenda, L.; Jimale, S. M.; Mukama, P. A. | Infectious Diseases | 10.1016/j.ijid.2020.09.1184 | <Go to ISI>://WOS:000612135101347 |
| 2023 | Zoonoses research in Somalia: A scoping review using a One Health approach | One Health | Mumin, F. I.; Fenton, A.; Osman, A. Y.; Mor, S. M. | Zoonoses | 10.1016/j.onehlt.2023.100626 | <Go to ISI>://WOS:001081387900001 |
| 1990 | CAMEL PAPILLOMATOSIS IN SOMALIA | Journal of Veterinary Medicine Series B-Infectious Diseases and Veterinary Public Health | Munz, E.; Moallin, A. S. M.; Mahnel, H.; Reimann, M. | Veterinary Sciences | 10.1111/j.1439-0450.1990.tb01046.x | <Go to ISI>://WOS:A1990DH75300006 |
| 1999 | IgM antibodies in hospitalized children with febrile illness during an inter-epidemic period of measles, in Somalia | Journal of Clinical Virology | Nur, Y. A.; Groen, J.; Yusuf, M. A.; Osterhaus, Adme | measles | 10.1016/s1386-6532(98)00002-x | <Go to ISI>://WOS:000078699700004 |
| 1979 | E-ANTIGEN (HBEAG) IN LEPROSY | International Journal of Leprosy and Other Mycobacterial Diseases | Nuti, M.; Tarabini, C. G.; Tarabini, C. G. L.; Thamer, G. | Microbiology |  | <Go to ISI>://WOS:A1979HF06000037 |
| 1979 | LEPROSY AND HEPATITIS-B VIRUS MARKERS - INCIDENCE OF HBSAG AND HBEAG IN SOMALIAN PATIENTS | International Journal of Leprosy and Other Mycobacterial Diseases | Nuti, M.; Tarabini, G.; Palermo, P.; Tarabini, G. L.; Thamer, G. | Microbiology |  | <Go to ISI>://WOS:A1979JA95900004 |
| 2023 | Clinical Characteristics of Acute Kidney Injury Associated with Tropical Acute Febrile Illness | Tropical Medicine and Infectious Disease | Omar, F. D.; Phumratanaprapin, W.; Silachamroon, U.; Hanboonkunupakarn, B.; Sriboonvorakul, N.; Thaipadungpanit, J.; Pan-ngum, W. | acute kidney injury | 10.3390/tropicalmed8030147 | <Go to ISI>://WOS:000959039200001 |
| 2024 | <i>Bartonella</i> species in dromedaries and ruminants from Lower Shabelle and Benadir regions, Somalia | Zoonoses and Public Health | Osman, A. M.; Hassan-Kadle, A. A.; Dias, C. M.; Ibrahim, A. M.; Collere, F. C. M.; Shair, M. A.; Montiani-Ferreira, F.; André, M. R.; Yusuf, A. A.; Vieira, Tswj; Machado, R. Z.; Vieira, R. F. C. | Bartonella bovis | 10.1111/zph.13158 | <Go to ISI>://WOS:001235800000001 |
| 2021 | Seroprevalence of Newcastle disease in backyard chickens in selected districts of Banadir region, Somalia | Tropical Animal Health and Production | Osman, A. M.; Oladele, O. A.; Ibrahim, A. M.; Mahamoud, M. A.; Mohamed, M. A.; Nwachukwu, N. O. | Newcastle disease | 10.1007/s11250-021-02622-5 | <Go to ISI>://WOS:000625109900001 |
| 2023 | Evaluating Fermentation Quality, Aerobic Stability, and Rumen-Degradation (In Situ) Characteristics of Various Protein-Based Total Mixed Rations | Animals | Rehemujiang, H.; Yusuf, H. A.; Ma, T.; Diao, Q. Y.; Kong, L. X.; Kang, L. Y.; Tu, Y. | aerobic stability | 10.3390/ani13172730 | <Go to ISI>://WOS:001144788600001 |
| 2022 | Do Spiders Ride on the Fear of Scorpions? A Cross-Cultural Eye Tracking Study | Animals | Rudolfová, V.; Stolhoferová, I.; Elmi, H. S. A.; Rádlová, S.; Rexová, K.; Berti, D. A.; Král, D.; Sommer, D.; Landová, E.; Frydlová, P.; Frynta, D. | Africa | 10.3390/ani12243466 | <Go to ISI>://WOS:000902131000001 |
| 2019 | Sick and solo: a qualitative study on the life experiences of people living with HIV in Somalia | Hiv Aids-Research and Palliative Care | Salad, A. M.; Mohamed, A.; Da'ar, O. B.; Abdikarim, A.; Kour, P.; Shrestha, M.; Gele, A. A. | HIV stigma | 10.2147/hiv.S185040 | <Go to ISI>://WOS:000461475100001 |
| 2016 | Modelling the potential benefits of different strategies to control infection with <i>Trypanosoma evansi</i> in camels in Somaliland | Tropical Animal Health and Production | Salah, A.; Robertson, I.; Mohamed, A. S. | Trypanosoma evansi | 10.1007/s11250-015-0942-0 | <Go to ISI>://WOS:000369009100026 |
| 2015 | Estimating the economic impact of <i>Trypanosoma evansi</i> infection on production of camel herds in Somaliland | Tropical Animal Health and Production | Salah, A. A.; Robertson, I.; Mohamed, A. | Trypanosoma evansi | 10.1007/s11250-015-0780-0 | <Go to ISI>://WOS:000351532400010 |
| 2019 | Prevalence and distribution of <i>Trypanosoma evansi</i> in camels in Somaliland | Tropical Animal Health and Production | Salah, A. A.; Robertson, I. D.; Mohamed, A. S. | Trypanosoma evansi | 10.1007/s11250-019-01947-6 | <Go to ISI>://WOS:000494799400031 |
| 1984 | OBSERVATIONS ON BLOOD-BORNE PARASITES OF DOMESTIC LIVESTOCK IN THE LOWER JUBA REGION OF SOMALIA | Tropical Animal Health and Production | Schoepf, K.; Mohamed, H. A. M.; Katende, J. M. | Agriculture | 10.1007/bf02265329 | <Go to ISI>://WOS:A1984TY37600008 |
| 1991 | LOW PREVALENCE OF HUMAN IMMUNODEFICIENCY VIRUS-1 (HIV-1), HIV-2, AND HUMAN T-CELL LYMPHOTROPIC VIRUS-1 INFECTION IN SOMALIA | American Journal of Tropical Medicine and Hygiene | Scott, D. A.; Corwin, A. L.; Constantine, N. T.; Omar, M. A.; Guled, A.; Yusef, M.; Roberts, C. R.; Watts, D. M. | htlv-i antibodies | 10.4269/ajtmh.1991.45.653 | <Go to ISI>://WOS:A1991HA01500001 |
| 2023 | Indigestible foreign bodies in the forestomach of slaughtered goats in Mogadishu, Somalia | Veterinary World | Shair, M. A.; Hassan-Kadle, A. A.; Osman, A. M.; Ahmed, K. M. Y.; Yusuf, A. A.; Barros-Filho, I. R.; Vieira, R. F. C. | Capra hircus | 10.14202/vetworld.2023.1829-1832 | <Go to ISI>://WOS:001098259000008 |
| 1990 | REINFECTION OF SOMALI CHILDREN WITH TRICHURIS-TRICHIURA AFTER CHEMOTHERAPY - RELEVANCE OF IMMUNOSTIMULATION | Transactions of the Royal Society of Tropical Medicine and Hygiene | Shiddo, S.; Ilardi, I.; Mussa, C.; Mohamud, M. A.; Aceti, A.; Leone, F.; Sebastiani, A.; Laghi; Amiconi, G. | population-dynamics | 10.1016/0035-9203(90)90100-s | <Go to ISI>://WOS:A1990ER99800029 |
| 1995 | VISCERAL LEISHMANIASIS IN SOMALIA - PREVALENCE OF LEISHMANIN-POSITIVE AND SEROPOSITIVE INHABITANTS IN AN ENDEMIC AREA | Transactions of the Royal Society of Tropical Medicine and Hygiene | Shiddo, S. A.; Akuffo, H. O.; Mohamed, A. A.; Huldt, G.; Nilsson, L. A.; Ouchterlony, O.; Thorstensson, R. | leishmaniasis | 10.1016/0035-9203(95)90640-1 | <Go to ISI>://WOS:A1995QY30000004 |
| 1994 | REFERENCE RANGES FOR IGG, IGM AND IGA IN THE SERUM OF URBAN AND RURAL SOMALIS | Tropical and Geographical Medicine | Shiddo, S. A.; Huldt, G.; Jama, H.; Nilsson, L. A.; Ouchterlony, O.; Warsame, M.; Jonsson, J. | immunoglobulin levels |  | <Go to ISI>://WOS:A1994MY89300008 |
| 1995 | VISCERAL LEISHMANIASIS IN SOMALIA - PREVALENCE OF MARKERS OF INFECTION AND DISEASE MANIFESTATIONS IN A VILLAGE IN AN ENDEMIC AREA | Transactions of the Royal Society of Tropical Medicine and Hygiene | Shiddo, S. A.; Mohamed, A. A.; Akuffo, H. O.; Mohamud, K. A.; Herzi, A. A.; Mohamed, H. H.; Huldt, G.; Nilsson, L. A.; Ouchterlony, O.; Thorstensson, R. | leishmaniasis | 10.1016/0035-9203(95)90008-x | <Go to ISI>://WOS:A1995RT59500006 |
| 1995 | VISCERAL LEISHMANIASIS IN SOMALIA - CIRCULATING ANTIBODIES AS MEASURED BY DAT, IMMUNOFLUORESCENCE AND ELISA | Tropical and Geographical Medicine | Shiddo, S. A.; Mohamed, A. A.; Huldt, C.; Loftenius, A.; Nilsson, L. A.; Jonsson, J.; Ouchterlony, O.; Thorstensson, R. | visceral leishmaniasis |  | <Go to ISI>://WOS:A1995QY26900005 |
| 2022 | Assessment of Polypharmacy, Drug Use Patterns, and Associated Factors at the Edna Adan University Hospital, Hargeisa, Somaliland | Journal of Tropical Medicine | Sidamo, T.; Deboch, A.; Abdi, M.; Debebe, F.; Dayib, K.; Balla, T. B. | elderly-patients | 10.1155/2022/2858987 | <Go to ISI>://WOS:000853682500001 |
| 1979 | RESULTS OF THE LEPROMIN TEST USING HUMAN LEPROMIN (H) AND ARMADILLO LEPROMIN (A) | International Journal of Leprosy and Other Mycobacterial Diseases | Tarabinicastellani, G.; Tarabinicastellani, G. L.; Nuti, M. | Microbiology |  | <Go to ISI>://WOS:A1979HF06000180 |
| 1979 | LEPROSY CONTROL CAMPAIGN IN SOMALIA - INTEGRATION INTO THE NATIONAL-HEALTH-SERVICES | International Journal of Leprosy and Other Mycobacterial Diseases | Tarabinicastellani, G. L.; Tarabinicastellani, G. | Microbiology |  | <Go to ISI>://WOS:A1979HF06000021 |
| 1993 | ENZYME-LINKED STAINING METHOD FOR LIGHT-MICROSCOPIC DETECTION OF ANTIBODIES TO PARASITE ANTIGENS ON THE MEMBRANE OF PLASMODIUM-FALCIPARUM-INFECTED ERYTHROCYTES | Acta Tropica | Udomsangpetch, R.; Kulane, A.; Berzins, K.; Perlmann, H.; Perlmann, P.; Flyg, B. W. | malaria | 10.1016/0001-706x(93)90050-l | <Go to ISI>://WOS:A1993MD23100008 |
| 1981 | STUDIES ON THE TRANSMISSION OF SCHISTOSOMA-HAEMATOBIUM AND THE BIONOMICS OF BULINUS (PH) ABYSSINICUS IN THE SOMALI-DEMOCRATIC-REPUBLIC | Annals of Tropical Medicine and Parasitology | Upatham, E. S.; Koura, M.; Ahmed, M. D.; Awad, A. H. | Public, Environmental & Occupational Health | 10.1080/00034983.1981.11687409 | <Go to ISI>://WOS:A1981LC43600009 |
| 2022 | Social contacts and other risk factors for respiratory infections among internally displaced people in Somaliland | Epidemics | van Zandvoort, K.; Bobe, M. O.; Hassan, A. I.; Abdi, M. I.; Ahmed, M. S.; Soleman, S. M.; Warsame, M. Y.; Wais, M. A.; Diggle, E.; McGowan, C. R.; Satzke, C.; Mulholland, K.; Egeh, M. M.; Hassan, M. M.; Hergeeye, M. A.; Eggo, R. M.; Checchi, F.; Flasche, S. | Contact data | 10.1016/j.epidem.2022.100625 | <Go to ISI>://WOS:000862280500003 |
| 1984 | FATAL INFECTION OF AN ELEPHANT CALF CAUSED BY THE TREMATODE PROTOFASCIOLA-ROBUSTA (LORENZ, 1881) IN SOMALILAND | Zentralblatt Fur Veterinarmedizin Reihe B-Journal of Veterinary Medicine Series B-Infectious Diseases Immunology Food Hygiene Veterinary Public Health | Vitovec, J.; Kotrla, B.; Haji, H.; Hayles, L. B. | Veterinary Sciences |  | <Go to ISI>://WOS:A1984TP19600003 |
| 1980 | INDIRECT IMMUNOPEROXYDASE TEST IN THE SEROLOGICAL DIAGNOSIS OF URINARY SCHISTOSOMIASIS | Tropical and Geographical Medicine | Vullo, V.; Ferone, U.; Aceti, A.; Petracca, C.; Nuti, M.; Delia, M. | Public, Environmental & Occupational Health |  | <Go to ISI>://WOS:A1980JX78700004 |
| 2021 | Excess mortality during the COVID-19 pandemic: a geospatial and statistical analysis in Mogadishu, Somalia | International Journal of Infectious Diseases | Warsame, A.; Bashiir, F.; Freemantle, T.; Williams, C.; Vazquez, Y.; Reeve, C.; Aweis, A.; Ahmed, M.; Checchi, F.; Dalmar, A. | mortality | 10.1016/j.ijid.2021.09.049 | <Go to ISI>://WOS:000718302600030 |
| 1988 | SUSCEPTIBILITY OF PLASMODIUM-FALCIPARUM TO CHLOROQUINE AND MEFLOQUINE IN SOMALIA | Transactions of the Royal Society of Tropical Medicine and Hygiene | Warsame, M.; Lebbad, M.; Ali, S.; Wernsdorfer, W. H.; Bjorkman, A. | Public, Environmental & Occupational Health | 10.1016/0035-9203(88)90409-9 | <Go to ISI>://WOS:A1988N315400007 |
| 1989 | THE SEROREACTIVITY AGAINST PF155 (RESA) ANTIGEN IN VILLAGERS FROM A MESOENDEMIC AREA IN SOMALIA | Tropical Medicine and Parasitology | Warsame, M.; Perlmann, H.; Ali, S.; Hagi, H.; Farah, S.; Lebbad, M.; Bjorkman, A. | Parasitology |  | <Go to ISI>://WOS:A1989CF14100006 |
| 1992 | LACK OF EFFECT OF DESIPRAMINE ON THE RESPONSE TO CHLOROQUINE OF PATIENTS WITH CHLOROQUINE-RESISTANT FALCIPARUM-MALARIA | Transactions of the Royal Society of Tropical Medicine and Hygiene | Warsame, M.; Wernsdorfer, W. H.; Bjorkman, A. | plasmodium-falciparum | 10.1016/0035-9203(92)90288-n | <Go to ISI>://WOS:A1992JD30400007 |
| 1990 | ISOLATED MALARIA OUTBREAK IN SOMALIA - ROLE OF CHLOROQUINE-RESISTANT PLASMODIUM-FALCIPARUM DEMONSTRATED IN BALCAD EPIDEMIC | Journal of Tropical Medicine and Hygiene | Warsame, M.; Wernsdorfer, W. H.; Ericsson, O.; Bjorkman, A. | Public, Environmental & Occupational Health |  | <Go to ISI>://WOS:A1990DV66900007 |
| 1991 | POSITIVE RELATIONSHIP BETWEEN THE RESPONSE OF PLASMODIUM-FALCIPARUM TO CHLOROQUINE AND PYRONARIDINE | Transactions of the Royal Society of Tropical Medicine and Hygiene | Warsame, M.; Wernsdorfer, W. H.; Payne, D.; Bjorkman, A. | invitro activity | 10.1016/0035-9203(91)90345-y | <Go to ISI>://WOS:A1991GM44600003 |
| 1991 | SUSCEPTIBILITY OF PLASMODIUM-FALCIPARUM INVITRO TO CHLOROQUINE, MEFLOQUINE, QUININE AND SULFADOXINE PYRIMETHAMINE IN SOMALIA - RELATIONSHIPS BETWEEN THE RESPONSES TO THE DIFFERENT DRUGS | Transactions of the Royal Society of Tropical Medicine and Hygiene | Warsame, M.; Wernsdorfer, W. H.; Payne, D.; Bjorkman, A. | sensitivity | 10.1016/0035-9203(91)90343-w | <Go to ISI>://WOS:A1991GM44600002 |
| 1991 | THE CHANGING PATTERN OF PLASMODIUM-FALCIPARUM SUSCEPTIBILITY TO CHLOROQUINE BUT NOT TO MEFLOQUINE IN A MESOENDEMIC AREA OF SOMALIA | Transactions of the Royal Society of Tropical Medicine and Hygiene | Warsame, M.; Wernsdorfer, W. H.; Willcox, M.; Kulane, A. A.; Bjorkman, A. | resistance | 10.1016/0035-9203(91)90018-t | <Go to ISI>://WOS:A1991FN69100013 |
| 1979 | BOVINE BRUCELLOSIS IN THE SOUTHERN REGIONS OF THE SOMALI-DEMOCRATIC-REPUBLIC | Tropical Animal Health and Production | Wernery, U.; Kerani, A. A.; Viertel, P. | Agriculture | 10.1007/bf02237763 | <Go to ISI>://WOS:A1979GM75900007 |
| 2013 | Applying the ICMJE authorship criteria to operational research in low-income countries: the need to engage programme managers and policy makers | Tropical Medicine & International Health | Zachariah, R.; Reid, T.; Van den Bergh, R.; Dahmane, A.; Kosgei, R. J.; Hinderaker, S. G.; Tayler-Smith, K.; Manzi, M.; Kizito, W.; Khogali, M.; Kumar, A. M. V.; Baruani, B.; Bishinga, A.; Kilale, A. M.; Nqobili, M.; Patten, G.; Sobry, A.; Cheti, E.; Nakanwagi, A.; Enarson, D. A.; Edginton, M. E.... | ICMJE | 10.1111/tmi.12133 | <Go to ISI>://WOS:000321504000015 |

#### Database: PUBMED NTDS

Total records retrieved: 193

| Year | Title | Journal | Authors | Keywords | DOI | URL |
| --- | --- | --- | --- | --- | --- | --- |
| 1980 | Schistosomiasis investigation in Somalia | Chin Med J (Engl) |  | Adolescent |  |  |
| 2016 | Global, regional, and national incidence, prevalence, and years lived with disability for 310 diseases and injuries, 1990-2015: a systematic analysis for the Global Burden of Disease Study 2015 | Lancet |  | *Bayes Theorem | 10.1016/s0140-6736(16)31678-6 |  |
| 2017 | Control of visceral leishmaniasis in Somalia: achievements in a challenging scenario, 2013–2015 | Wkly Epidemiol Rec |  | Age Distribution |  |  |
| 2017 | Control of visceral leishmaniasis in Somalia: achievements in a challenging scenario, 2013–2015 | Wkly Epidemiol Rec |  |  |  |  |
| 2020 | Visceral Leishmaniasis, Northern Somalia, 2013-2019 | Emerg Infect Dis | Aalto, M. K.; Sunyoto, T.; Yusuf, M. A. A.; Mohamed, A. A.; Van der Auwera, G.; Dujardin, J. C. | Adolescent | 10.3201/eid2601.181851 |  |
| 2020 | Prevalence of refractive error and visual impairment among school-age children of Hargesia, Somaliland, Somalia | East Mediterr Health J | Abdi Ahmed, Z.; Alrasheed, S. H.; Alghamdi, W. | Child | 10.26719/emhj.20.077 |  |
| 1999 | Health for All by the Year 2000: what about the nomads? | Dev Pract | Abu Omar, M.; Omar, M. M. | *Delivery of Health Care | 10.1080/09614529953043 |  |
| 1989 | Field trial of the efficacy of a simplified and standard metrifonate treatments of Schistosoma haematobium | Eur J Clin Pharmacol | Aden Abdi, Y.; Gustafsson, L. L. | Adolescent | 10.1007/bf00558502 |  |
| 1989 | Poor patient compliance reduces the efficacy of metrifonate treatment of Schistosoma haematobium in Somalia | Eur J Clin Pharmacol | Aden Abdi, Y.; Gustafsson, L. L. | Adolescent | 10.1007/bf00609189 |  |
| 1987 | A simplified dosage schedule of metrifonate in the treatment of Schistosoma haematobium infection in Somalia | Eur J Clin Pharmacol | Aden-Abdi, Y.; Gustafsson, L. L.; Elmi, S. A. | Colic/chemically induced | 10.1007/bf00637666 |  |
| 2024 | Brucellosis in camel, small ruminants, and Somali pastoralists in Eastern Ethiopia: a One Health approach | Front Vet Sci | Ahad, A. A.; Megersa, B.; Edao, B. M. | Somali region | 10.3389/fvets.2024.1276275 |  |
| 2014 | Guinea worm (Dracunculiasis) eradication: update on progress and endgame challenges | Trans R Soc Trop Med Hyg | Al-Awadi, A. R.; Al-Kuhlani, A.; Breman, J. G.; Doumbo, O.; Eberhard, M. L.; Guiguemde, R. T.; Magnussen, P.; Molyneux, D. H.; Nadim, A. | Africa/epidemiology | 10.1093/trstmh/tru039 |  |
| 2023 | Public health impact of the spread of Anopheles stephensi in the WHO Eastern Mediterranean Region countries in Horn of Africa and Yemen: need for integrated vector surveillance and control | Malar J | Al-Eryani, S. M.; Irish, S. R.; Carter, T. E.; Lenhart, A.; Aljasari, A.; Montoya, L. F.; Awash, A. A.; Mohammed, E.; Ali, S.; Esmail, M. A.; Hussain, A.; Amran, J. G.; Kayad, S.; Nouredayem, M.; Adam, M. A.; Azkoul, L.; Assada, M.; Baheshm, Y. A.; Eltahir, W.; Hutin, Y. J. | Animals | 10.1186/s12936-023-04545-y |  |
| 2014 | Cystic hydatidosis in slaughtered goats from various municipal abattoirs in Oman | Trop Anim Health Prod | Al-Kitani, F.; Baqir, S.; Hussain, M. H.; Roberts, D. | *Abattoirs | 10.1007/s11250-014-0646-x |  |
| 2016 | A review of visceral leishmaniasis during the conflict in South Sudan and the consequences for East African countries | Parasit Vectors | Al-Salem, W.; Herricks, J. R.; Hotez, P. J. | Humans | 10.1186/s13071-016-1743-7 |  |
| 2019 | Epidemiology of visceral leishmaniasis in Shebelle Zone of Somali Region, eastern Ethiopia | Parasit Vectors | Alebie, G.; Worku, A.; Yohannes, S.; Urga, B.; Hailu, A.; Tadesse, D. | Acacia | 10.1186/s13071-019-3452-5 |  |
| 2024 | A Case Study on Unreported First Probable Human Rabies Following Honey Badger in Somalia | Open Access Emerg Med | Ali Osman, U. M.; Turfan, S.; Farah Yusuf Mohamud, M. | Somalia | 10.2147/oaem.S439996 |  |
| 1986 | Role of community health workers in trachoma control. Case study from a Somali refugee camp | Trop Doct | Anderson, J. D.; Bentley, C. C. | Adolescent | 10.1177/004947558601600209 |  |
| 2022 | Public Health Surveillance for Adverse Events Following COVID-19 Vaccination in Africa | Vaccines (Basel) | Anjorin, A. A.; Odetokun, I. A.; Nyandwi, J. B.; Elnadi, H.; Awiagah, K. S.; Eyedo, J.; Abioye, A. I.; Gachara, G.; Maisara, A. M.; Razouqi, Y.; Yusuf Mohamud, M. F.; Mhgoob, Z. E.; Ajayi, T.; Ntirenganya, L.; Saibu, M.; Salako, B. L.; Elelu, N.; Wright, K. O.; Fasina, F. O.; Mosbah, R. | COVID-19 vaccine | 10.3390/vaccines10040546 |  |
| 2024 | Epidemiology of Echinococcus granulosus sensu lato in the Greater Horn of Africa: A systematic review | PLoS Negl Trop Dis | Aregawi, W. G.; Levecke, B.; Ashenafi, H.; Byaruhanga, C.; Kebede, N.; Mulinge, E.; Wassermann, M.; Romig, T.; Dorny, P.; Dermauw, V. | Cattle | 10.1371/journal.pntd.0011894 |  |
| 1975 | Studies on schistosomiasis in Somalia | Am J Trop Med Hyg | Arfaa, F. | Animals | 10.4269/ajtmh.1975.24.280 |  |
| 2019 | Multiplex serology demonstrate cumulative prevalence and spatial distribution of malaria in Ethiopia | Malar J | Assefa, A.; Ali Ahmed, A.; Deressa, W.; Sime, H.; Mohammed, H.; Kebede, A.; Solomon, H.; Teka, H.; Gurrala, K.; Matei, B.; Wakeman, B.; Wilson, G. G.; Sinha, I.; Maude, R. J.; Ashton, R.; Cook, J.; Shi, Y. P.; Drakeley, C.; von Seidlein, L.; Rogier, E.; Hwang, J. | Adolescent | 10.1186/s12936-019-2874-z |  |
| 1956 | Bilharziasis survey in British Somaliland, Eritrea, Ethiopia, Somalia, the Sudan, and Yemen | Bull World Health Organ | Ayad, N. | Africa, Eastern |  |  |
| 2015 | Complete genome sequencing and phylogenetic analysis of dengue type 1 virus isolated from Jeddah, Saudi Arabia | Virol J | Azhar, E. I.; Hashem, A. M.; El-Kafrawy, S. A.; Abol-Ela, S.; Abd-Alla, A. M.; Sohrab, S. S.; Farraj, S. A.; Othman, N. A.; Ben-Helaby, H. G.; Ashshi, A.; Madani, T. A.; Jamjoom, G. | Adult | 10.1186/s12985-014-0235-7 |  |
| 2009 | A 48-year-old Somali woman with hip pain | Clin Infect Dis | Babady, N. E.; Pritt, B. S.; Walker, R. C.; Rosenblatt, J. E.; Binnicker, M. J. | Albendazole/therapeutic use | 10.1086/605083 |  |
| 2023 | Diagnostic capacity for cutaneous fungal diseases in the African continent | Int J Dermatol | Badiane, A. S.; Ramarozatovo, L. S.; Doumbo, S. N.; Dorkenoo, A. M.; Mandengue, C.; Dunaisk, C. M.; Ball, M.; Dia, M. K.; Ngaya, G. S. L.; Mahamat, H. H.; Kalombo, H.; Bah, A.; Cá, Z.; Langa, J. C.; Mohamed, A. M.; Mokomane, M.; Ahmed, S. A.; Rapalanoro Rabenja, F.; Hay, R. J.; Penney, R. O. S.; ... | Humans | 10.1111/ijd.16751 |  |
| 2022 | Somalia tackles leprosy and visceral leishmaniasis | Lancet Infect Dis | Bagcchi, S. | Humans | 10.1016/s1473-3099(22)00168-2 |  |
| 2022 | Third wave in India and an update on vaccination: A short communication | Ann Med Surg (Lond) | Baig, R.; Mateen, M. A.; Aborode, A. T.; Novman, S.; Abdul Matheen, I.; Siddiqui, O. S.; Ahmed, F. A. | Covid-19 | 10.1016/j.amsu.2022.103414 |  |
| 1960 | Antimony dimercaptosuccinate (TWSb) in the treatment of urinary bilharziasis in Somalia | Cent Afr J Med | Baruffa, G.; Friedheim, E. A. | Antimony/*therapy |  |  |
| 2023 | ICU-Managed Patients' Epidemiology, Characteristics, and Outcomes: A Retrospective Single-Center Study | Anesthesiol Res Pract | Bashir, A. M.; Osman, M. M.; Mohamed, H. N.; Hilowle, I. A.; Ahmed, H. A.; Osman, A. A.; Fiidow, O. A. |  | 10.1155/2023/9388449 |  |
| 2023 | "It is difficult for us to treat their pain". Health professionals' perceptions of Somali pastoralists in the context of pain management: a conceptual model | Med Humanit | Baum, E.; Abdi, S.; van Eeuwijk, P.; Probst-Hensch, N.; Zinsstag, J.; Tschopp, R.; Vosseler, B. | Humans | 10.1136/medhum-2022-012570 |  |
| 2018 | Visceral leishmaniasis in selected communities of Hamar and Banna-Tsamai districts in Lower Omo Valley, South West Ethiopia: Sero-epidemological and Leishmanin Skin Test Surveys | PLoS One | Bekele, F.; Belay, T.; Zeynudin, A.; Hailu, A. | Adolescent | 10.1371/journal.pone.0197430 |  |
| 1999 | Stories of growing up amid violence by refugee children of war and children of battered women living in Canada | Image J Nurs Sch | Berman, H. | Adolescent | 10.1111/j.1547-5069.1999.tb00422.x |  |
| 2020 | Emergence of Undetectable Malaria Parasites: A Threat under the Radar amid the COVID-19 Pandemic? | Am J Trop Med Hyg | Beshir, K. B.; Grignard, L.; Hajissa, K.; Mohammed, A.; Nurhussein, A. M.; Ishengoma, D. S.; Lubis, I. N. D.; Drakeley, C. J.; Sutherland, C. J. | Africa | 10.4269/ajtmh.20-0467 |  |
| 2024 | Assessing Vaccination Delivery Strategies for Zero-Dose and Under-Immunized Children in the Fragile Context of Somalia | Vaccines (Basel) | Bile, A. S.; Ali-Salad, M. A.; Mahmoud, A. J.; Singh, N. S.; Abdelmagid, N.; Sabahelzain, M. M.; Checchi, F.; Mounier-Jack, S.; Nor, B. | conflict | 10.3390/vaccines12020154 |  |
| 1996 | Susceptibility of Ethiopian bulinid snails to Schistosoma haematobium from Somalia | East Afr Med J | Birrie, H.; Balcha, F.; Bizuneh, A.; Bero, G. | Animals |  |  |
| 1993 | Schistosoma haematobium infection among Ethiopian prisoners of war (1977-1988) returning from Somalia | Ethiop Med J | Birrie, H.; Berhe, N.; Tedla, S.; Gemeda, N. | Adult |  |  |
| 2001 | Schistosoma haematobium | N Engl J Med | Boehm, D.; Pryce, D. J. | Diagnosis, Differential | 10.1056/nejm200104123441515 |  |
| 2009 | Schistosomiasis: a case study | Urol Nurs | Borch, M.; Kiernan, M.; Rust, K.; Baron, B.; Simmons, B.; Hattala, P.; Davey, A.; Yovanovich, J.; Shayder, D.; Wasilewski, A.; LaFaro, V. E. | Adult |  |  |
| 1989 | Serological evidence of dengue fever among refugees, Hargeysa, Somalia | J Med Virol | Botros, B. A.; Watts, D. M.; Soliman, A. K.; Salib, A. W.; Moussa, M. I.; Mursal, H.; Douglas, C.; Farah, M. | Antibodies, Viral/immunology | 10.1002/jmv.1890290202 |  |
| 2020 | Plasmodium falciparum isolate with histidine-rich protein 2 gene deletion from Nyala City, Western Sudan | Sci Rep | Boush, M. A.; Djibrine, M. A.; Mussa, A.; Talib, M.; Maki, A.; Mohammed, A.; Beshir, K. B.; Mohamed, Z.; Hajissa, K. | Antigens, Protozoan/*genetics | 10.1038/s41598-020-69756-8 |  |
| 2001 | Visceral leishmaniasis (kala-azar) outbreak in Somali refugees and Kenyan shepherds, Kenya | Emerg Infect Dis | Boussery, G.; Boelaert, M.; van Peteghem, J.; Ejikon, P.; Henckaerts, K. | Adolescent | 10.3201/eid0707.010746 |  |
| 2009 | Community based parasitic screening and treatment of Sudanese refugees: application and assessment of Centers for Disease Control guidelines | Am J Trop Med Hyg | Brodine, S. K.; Thomas, A.; Huang, R.; Harbertson, J.; Mehta, S.; Leake, J.; Nutman, T.; Moser, K.; Wolf, J.; Ramanathan, R.; Burbelo, P.; Nou, J.; Wilkins, P.; Reed, S. L. | Adolescent |  |  |
| 1982 | Human leptospirosis in Somalia: a serological survey | Trans R Soc Trop Med Hyg | Cacciapuoti, B.; Nuti, M.; Pinto, A.; Sabrie, A. M. | Adolescent | 10.1016/0035-9203(82)90270-x |  |
| 1968 | Clinical and epidemiological patterns of leishmaniasis in Africa | Trop Geogr Med | Cahill, K. M. | Adolescent |  |  |
| 1971 | Studies in Somalia | Trans R Soc Trop Med Hyg | Cahill, K. M. | Complement Fixation Tests | 10.1016/0035-9203(71)90182-9 |  |
| 1968 | Schistosomiasis in Somalia. A parasitological and serological survey in Giohar | Trans R Soc Trop Med Hyg | Cahill, K. M.; Kagan, I. | Africa, Eastern | 10.1016/0035-9203(68)90161-2 |  |
| 2011 | A cluster of cutaneous leishmaniasis associated with human smuggling | Am J Trop Med Hyg | Cannella, A. P.; Nguyen, B. M.; Piggott, C. D.; Lee, R. A.; Vinetz, J. M.; Mehta, S. R. | Adult | 10.4269/ajtmh.2011.10-0693 |  |
| 2020 | Clinical characteristics and treatment of actinomycetoma in northeast Mexico: A case series | PLoS Negl Trop Dis | Cárdenas-de la Garza, J. A.; Welsh, O.; Cuéllar-Barboza, A.; Suarez-Sánchez, K. P.; De la Cruz-Valadez, E.; Cruz-Gómez, L. G.; Gallardo-Rocha, A.; Ocampo-Candiani, J.; Vera-Cabrera, L. | Adolescent | 10.1371/journal.pntd.0008123 |  |
| 2021 | Skin Mycetoma in an 11-Year-Old African Boy: Case Presentation with Emphasis on Histopathological Features and Differential Diagnosis | Dermatopathology (Basel) | Cazzato, G.; Colagrande, A.; Cimmino, A.; Lospalluti, L.; Demarco, A.; Foti, C.; Romita, P.; Arezzo, F.; Loizzi, V.; Parente, P.; Resta, L.; Ingravallo, G. | actinomycetes | 10.3390/dermatopathology8040053 |  |
| 2013 | First report of the visceral leishmaniasis vector Phlebotomus martini (Diptera: Psychodidae) in Tanzania | J Med Entomol | Clark, J. W.; Kioko, E.; Odemba, N.; Ngere, F.; Kamanza, J.; Oyugi, E.; Kerich, G.; Kimbita, E.; Bast, J. D. | Animals | 10.1603/me12147 |  |
| 1964 | THE SPECIES OF ONCHOCERCA IN CATTLE IN KENYA AND SOMALIA | Ann Trop Med Parasitol | Clarkson, M. J. | Animals | 10.1080/00034983.1964.11686225 |  |
| 2007 | Tuberculosis in complex emergencies | Bull World Health Organ | Coninx, R. | Antitubercular Agents/supply & distribution/*therapeutic use | 10.2471/blt.06.037630 |  |
| 2021 | From reverse innovation to global innovation in animal health: A review | Heliyon | Crump, L.; Maidane, Y.; Mauti, S.; Tschopp, R.; Ali, S. M.; Abtidon, R.; Bourhy, H.; Keita, Z.; Doumbia, S.; Traore, A.; Bonfoh, B.; Tetchi, M.; Tiembré, I.; Kallo, V.; Paithankar, V.; Zinsstag, J. | Contact sensor | 10.1016/j.heliyon.2021.e08044 |  |
| 1991 | Estimating maternal mortality in Djibouti: an application of the sisterhood method | Int J Epidemiol | David, P.; Kawar, S.; Graham, W. | Adolescent | 10.1093/ije/20.2.551 |  |
| 2017 | Estimating the number of cases of podoconiosis in Ethiopia using geostatistical methods | Wellcome Open Res | Deribe, K.; Cano, J.; Giorgi, E.; Pigott, D. M.; Golding, N.; Pullan, R. L.; Noor, A. M.; Cromwell, E. A.; Osgood-Zimmerman, A.; Enquselassie, F.; Hailu, A.; Murray, C. J. L.; Newport, M. J.; Brooker, S. J.; Hay, S. I.; Davey, G. | Ethiopia | 10.12688/wellcomeopenres.12483.2 |  |
| 2015 | Mapping and Modelling the Geographical Distribution and Environmental Limits of Podoconiosis in Ethiopia | PLoS Negl Trop Dis | Deribe, K.; Cano, J.; Newport, M. J.; Golding, N.; Pullan, R. L.; Sime, H.; Gebretsadik, A.; Assefa, A.; Kebede, A.; Hailu, A.; Rebollo, M. P.; Shafi, O.; Bockarie, M. J.; Aseffa, A.; Hay, S. I.; Reithinger, R.; Enquselassie, F.; Davey, G.; Brooker, S. J. | Elephantiasis/*epidemiology | 10.1371/journal.pntd.0003946 |  |
| 2021 | Human fascioliasis in Africa: A systematic review | PLoS One | Dermauw, V.; Muchai, J.; Al Kappany, Y.; Fajardo Castaneda, A. L.; Dorny, P. | Africa/epidemiology | 10.1371/journal.pone.0261166 |  |
| 2020 | Coxiella burnetii in Dromedary Camels (Camelus dromedarius): A Possible Threat for Humans and Livestock in North Africa and the Near and Middle East? | Front Vet Sci | Devaux, C. A.; Osman, I. O.; Million, M.; Raoult, D. | Coxiella burnetii | 10.3389/fvets.2020.558481 |  |
| 2018 | Infectious and dermatological diseases among arriving migrants on the Italian coasts | Eur J Public Health | Di Meco, E.; Di Napoli, A.; Amato, L. M.; Fortino, A.; Costanzo, G.; Rossi, A.; Mirisola, C.; Petrelli, A. | Adult | 10.1093/eurpub/cky126 |  |
| 2010 | First record of Ae. albopictus (Skuse 1894), in Central African Republic | Trop Med Int Health | Diallo, M.; Laganier, R.; Nangouma, A. | Aedes/*classification | 10.1111/j.1365-3156.2010.02594.x |  |
| 2024 | Mycetoma case series in Somalia | Trop Doct | Doğan, A.; Ali, A. M.; Ali, M. A.; Abdullahi İ, M. | Humans | 10.1177/00494755231201664 |  |
| 2018 | Epidemiology of trachoma and its implications for implementing the "SAFE" strategy in Somali Region, Ethiopia: results of 14 population-based prevalence surveys | Ophthalmic Epidemiol | Duale, A. B.; Negussu Ayele, N.; Macleod, C. K.; Kello, A. B.; Eshetu Gezachew, Z.; Binegdie, A.; Dejene, M.; Alemayehu, W.; Flueckiger, R. M.; Massae, P. A.; Willis, R.; Kebede Negash, B.; Solomon, A. W. | Adolescent | 10.1080/09286586.2017.1409358 |  |
| 2018 | Current and future distribution of Aedes aegypti and Aedes albopictus (Diptera: Culicidae) in WHO Eastern Mediterranean Region | Int J Health Geogr | Ducheyne, E.; Tran Minh, N. N.; Haddad, N.; Bryssinckx, W.; Buliva, E.; Simard, F.; Malik, M. R.; Charlier, J.; De Waele, V.; Mahmoud, O.; Mukhtar, M.; Bouattour, A.; Hussain, A.; Hendrickx, G.; Roiz, D. | *Aedes | 10.1186/s12942-018-0125-0 |  |
| 2010 | Madura foot - mind the soil | J Plast Reconstr Aesthet Surg | El Muttardi, N.; Kulendren, D.; Jemec, B. | Adolescent | 10.1016/j.bjps.2009.12.007 |  |
| 2003 | Lymphatic filariasis in the Eastern Mediterranean Region: current status and prospects for elimination | East Mediterr Health J | El Setouhy, M.; Ramzy, R. M. | Animals |  |  |
| 2000 | Evidence of Onchocerca fasciata (Filarioidea: Onchocercidae) in camels (Camelus dromedarius): I-prevalence, nodular lesions appearance and parasite morphology | Vet Parasitol | El-Massry, A. A.; Derbala, A. A. | Abdomen/parasitology | 10.1016/s0304-4017(99)00203-4 |  |
| 2011 | Ecology and control of the sand fly vectors of Leishmania donovani in East Africa, with special emphasis on Phlebotomus orientalis | J Vector Ecol | Elnaiem, D. E. | Africa | 10.1111/j.1948-7134.2011.00109.x |  |
| 2024 | Prevalence and associated factors of sexual dysfunction in female hemodialysis patients: first report from Somalia | BMC Womens Health | Eraslan, A.; Mohamed, A. H.; Bashir, A. M.; Adani, A. A.; Cimen, S. | Female | 10.1186/s12905-024-02902-w |  |
| 2020 | Hydatid cyst of the foot: a case report | J Med Case Rep | Ewnte, B. | Echinococcosis/*diagnosis/surgery | 10.1186/s13256-019-2337-8 |  |
| 2020 | Emergence of a novel chikungunya virus strain bearing the E1:V80A substitution, out of the Mombasa, Kenya 2017-2018 outbreak | PLoS One | Eyase, F.; Langat, S.; Berry, I. M.; Mulwa, F.; Nyunja, A.; Mutisya, J.; Owaka, S.; Limbaso, S.; Ofula, V.; Koka, H.; Koskei, E.; Lutomiah, J.; Jarman, R. G.; Sang, R. | Aedes/virology | 10.1371/journal.pone.0241754 |  |
| 2022 | Understanding and addressing COVID-19 vaccine hesitancy in low and middle income countries and in people with severe mental illness: Overview and recommendations for Latin America and the Caribbean | Front Psychiatry | Faria, C. G. F.; de Matos, U. M. A.; Llado-Medina, L.; Pereira-Sanchez, V.; Freire, R.; Nardi, A. E. | Covid-19 | 10.3389/fpsyt.2022.910410 |  |
| 2018 | Madura foot: an imported case of a non-common diagnosis | Infez Med | Fasciana, T.; Colomba, C.; Cervo, A.; Di Carlo, P.; Scarlata, F.; Mascarella, C.; Giammanco, A.; Cascio, A. | *Communicable Diseases, Imported/diagnosis/drug therapy |  |  |
| 2022 | Co-creation and priority setting for applied and implementation research in One Health: Improving capacities in public and animal health systems in Kenya | One Health | Fasina, F. O.; Nanyingi, M.; Wangila, R. S.; Gikonyo, S.; Omani, R.; Nyariki, T.; Wahome, L. W.; Kiplamai, J.; Tenge, E.; Kivaria, F.; Okuthe, S.; Nzietchueng, S.; Kimani, T.; Kimutai, J.; Mucheru, G.; Njagi, O.; Njogu, G.; Rono, R.; Maina, G. N.; Mogaka, D.; Mathooko, J.; Sirdar, M. M.; Mogoa, E... | Anthropology | 10.1016/j.onehlt.2022.100460 |  |
| 2014 | Malaria situation in an endemic area, southeastern iran | J Arthropod Borne Dis | Fekri, S.; Vatandoost, H.; Daryanavard, A.; Shahi, M.; Safari, R.; Raeisi, A.; Omar, A. S.; Sharif, M.; Azizi, A.; Ali, A. A.; Nasser, A.; Hasaballah, I.; Hanafi-Bojd, A. A. | Iran |  |  |
| 2018 | Larvicidal and pupicidal evaluation of silver nanoparticles synthesized using Aquilaria sinensis and Pogostemon cablin essential oils against dengue and zika viruses vector Aedes albopictus mosquito and its histopathological analysis | Artif Cells Nanomed Biotechnol | Ga'al, H.; Fouad, H.; Mao, G.; Tian, J.; Jianchu, M. | Aedes/*drug effects | 10.1080/21691401.2017.1365723 |  |
| 2023 | Increasing prevalence of malaria and acute dengue virus coinfection in Africa: a meta-analysis and meta-regression of cross-sectional studies | Malar J | Gebremariam, T. T.; Schallig, Hdfh; Kurmane, Z. M.; Danquah, J. B. | Humans | 10.1186/s12936-023-04723-y |  |
| 2023 | Correction: Increasing prevalence of malaria and acute dengue virus coinfection in Africa: a meta-analysis and meta-regression of cross-sectional studies | Malar J | Gebremariam, T. T.; Schallig, Hdfh; Kurmane, Z. M.; Danquah, J. B. |  | 10.1186/s12936-023-04771-4 |  |
| 2023 | Prevalence of trachoma in Somali region, Ethiopia: results from trachoma impact surveys in 50 woredas | Int Health | Gebreselassie, G.; Negash, K.; Tsegaye, S.; Makonnen, M.; Deneke, B.; Desalegn, M.; Harding-Esch, E. M.; Harte, A.; Solomon, A. W.; Boyd, S.; Bakhtiari, A.; Hassen, M. A.; Hambali, A.; Dejene, M.; Beckwith, C.; Tadesse, F.; Seifu, F.; Kiflu, G.; Kebede, F. | Humans | 10.1093/inthealth/ihad063 |  |
| 2012 | Dengue and US military operations from the Spanish-American War through today | Emerg Infect Dis | Gibbons, R. V.; Streitz, M.; Babina, T.; Fried, J. R. | Dengue/epidemiology/*history | 10.3201/eid1804.110134 |  |
| 1978 | Anaemia and Schistosoma haematobium infection in the North-Eastern Province of Kenya | Trans R Soc Trop Med Hyg | Greenham, R. | Adolescent | 10.1016/0035-9203(78)90304-8 |  |
| 2015 | Exploring Somali women's reproductive health knowledge and experiences: results from focus group discussions in Mogadishu | Reprod Health Matters | Gure, F.; Yusuf, M.; Foster, A. M. | Female | 10.1016/j.rhm.2015.11.018 |  |
| 2021 | Another dengue fever outbreak in Eastern Ethiopia-An emerging public health threat | PLoS Negl Trop Dis | Gutu, M. A.; Bekele, A.; Seid, Y.; Mohammed, Y.; Gemechu, F.; Woyessa, A. B.; Tayachew, A.; Dugasa, Y.; Gizachew, L.; Idosa, M.; Tokarz, R. E.; Sugerman, D. | Adolescent | 10.1371/journal.pntd.0008992 |  |
| 1990 | Antibody responses in schistosomiasis haematobium in Somalia. Relation to age and infection intensity | Ann Trop Med Parasitol | Hagi, H.; Huldt, G.; Loftenius, A.; Schröder, H. | Adolescent | 10.1080/00034983.1990.11812451 |  |
| 1989 | Silicate pneumoconiosis in camels (Camelus dromedarius L.) | Zentralbl Veterinarmed A | Hansen, H. J.; Jama, F. M.; Nilsson, C.; Norrgren, L.; Abdurahman, O. S. | Animals | 10.1111/j.1439-0442.1989.tb00793.x |  |
| 2023 | Tropical Data: Approach and Methodology as Applied to Trachoma Prevalence Surveys | Ophthalmic Epidemiol | Harding-Esch, E. M.; Burgert-Brucker, C. R.; Jimenez, C.; Bakhtiari, A.; Willis, R.; Bejiga, M. D.; Mpyet, C.; Ngondi, J.; Boyd, S.; Abdala, M.; Abdou, A.; Adamu, Y.; Alemayehu, A.; Alemayehu, W.; Al-Khatib, T.; Apadinuwe, S. C.; Awaca, N.; Awoussi, M. S.; Baayendag, G.; Badiane, M. D.; Bailey, R... | Humans | 10.1080/09286586.2023.2249546 |  |
| 2023 | A Late Diagnosis of Visceral Leishmaniasis Using Tru-Cut Biopsy of the Spleen and Malaria Co-Infection - A Diagnostic Challenge: A Case Report in Somalia | Infect Drug Resist | Hassan, M. A.; Omar, A. A.; Mohamed, I. A.; Garba, B.; Fuje, M. M. A.; Salad, S. O. | co-infection | 10.2147/idr.S420832 |  |
| 2021 | Rift Valley fever and Brucella spp. in ruminants, Somalia | BMC Vet Res | Hassan-Kadle, A. A.; Osman, A. M.; Shair, M. A.; Abdi, O. M.; Yusuf, A. A.; Ibrahim, A. M.; Vieira, R. F. C. | Animals | 10.1186/s12917-021-02980-0 |  |
| 2021 | Spectrum and Prevalence of Thyroid Diseases at a Tertiary Referral Hospital in Mogadishu, Somalia: A Retrospective Study of 976 Cases | Int J Endocrinol | Hassan-Kadle, M. A.; Adani, A. A.; Eker, H. H.; Keles, E.; Muse Osman, M.; Mahdi Ahmed, H.; Görçin Karaketir, Ş |  | 10.1155/2021/7154250 |  |
| 2018 | Epidemiology of viral hepatitis in Somalia: Systematic review and meta-analysis study | World J Gastroenterol | Hassan-Kadle, M. A.; Osman, M. S.; Ogurtsov, P. P. | Chronic Disease/epidemiology | 10.3748/wjg.v24.i34.3927 |  |
| 2014 | Recent outbreaks of rift valley Fever in East Africa and the middle East | Front Public Health | Himeidan, Y. E.; Kweka, E. J.; Mahgoub, M. M.; El Rayah el, A.; Ouma, J. O. | Aedes mosquitoes | 10.3389/fpubh.2014.00169 |  |
| 2017 | Urban Chikungunya in the Middle East and North Africa: A systematic review | PLoS Negl Trop Dis | Humphrey, J. M.; Cleton, N. B.; Reusken, Cbem; Glesby, M. J.; Koopmans, M. P. G.; Abu-Raddad, L. J. | Aedes/virology | 10.1371/journal.pntd.0005707 |  |
| 2017 | Microscopic and Molecular Detection of Camel Piroplasmosis in Gadarif State, Sudan | Vet Med Int | Ibrahim, A. M.; Kadle, A. A.; Nyingilili, H. S. |  | 10.1155/2017/9345231 |  |
| 2022 | Delayed treatment of neglected open knee dislocation; case report | Int J Surg Case Rep | Ibrahim, H. S.; Çiçek, E. I.; Taşkoparan, H.; Duran Hashi, A. Y. | Arthrodesis | 10.1016/j.ijscr.2022.106937 |  |
| 2023 | Neglected bilateral geno valgum deformity managed with a dynamic compression plate and the ilizarov construct: a case report | Ann Med Surg (Lond) | Ibrahim, Y. B.; Mohamed, A. Y.; Ibrahim, H. S.; Cicek, E. I.; Mohamed, A. H.; May, H. | case report | 10.1097/ms9.0000000000000405 |  |
| 1987 | Epidemiological study of parasitic infections in Somali nomads | Trans R Soc Trop Med Hyg | Ilardi, I.; Sebastiani, A.; Leone, F.; Madera, A.; Bile, M. K.; Shiddo, S. C.; Mohamed, H. H.; Amiconi, G. | Adolescent | 10.1016/0035-9203(87)90027-7 |  |
| 2023 | Sero-prevalence of visceral leishmaniasis and its associated factors among asymptomatic individuals visiting Denan health center, southeastern Ethiopia | Trop Dis Travel Med Vaccines | Ismail, A.; Yared, S.; Dugassa, S.; Abera, A.; Animut, A.; Erko, B.; Gebresilassie, A. | Denan | 10.1186/s40794-023-00196-8 |  |
| 2016 | Somalia: A Nation at the Crossroads of Extreme Poverty, Conflict, and Neglected Tropical Diseases | PLoS Negl Trop Dis | Jaffer, A.; Hotez, P. J. |  | 10.1371/journal.pntd.0004670 |  |
| 2024 | Twenty years of herpes simplex virus type 2 (HSV-2) research in low-income and middle-income countries: systematic evaluation of progress made in addressing WHO prioritiesfor research in HSV-2 epidemiology and diagnostics | BMJ Glob Health | Jama, M.; Owen, E. M.; Nahal, B.; Obasi, A.; Clarke, E. | Female | 10.1136/bmjgh-2023-012717 |  |
| 2009 | Appendectomy to remember | J Travel Med | Jama, S.; Manivel, J. C.; Abd Alla, M. D.; Stauffer, W. M. | Adult | 10.1111/j.1708-8305.2009.00306.x |  |
| 2024 | Retrospective study on the dengue fever outbreak in Puntland State, Somalia | BMC Infect Dis | Jama, S. S.; Abshir, S. N.; Jama, J. S.; Abdi, M. M. | Humans | 10.1186/s12879-024-09552-1 |  |
| 2024 | Correction: Retrospective study on the dengue fever outbreak in Puntland State, Somalia | BMC Infect Dis | Jama, S. S.; Abshir, S. N.; Jama, J. S.; Abdi, M. M. |  | 10.1186/s12879-024-09692-4 |  |
| 2023 | Rampart of Health-Specific Leadership and Social Support of Colleagues to Overcome Burnout in an Emotionally Demanding Situations: The Mediating Role of Stress | J Healthc Leadersh | Javaid, M. U.; Rehman, N.; Mirza, M. Z.; Ibrahim, A. M. | Malaysia | 10.2147/jhl.S420584 |  |
| 1995 | Helping the helper: 528th Combat Stress Center in Somalia | Mil Med | Jiggetts, S. M.; Hall, D. P., Jr. | Combat Disorders/physiopathology/psychology/*therapy |  |  |
| 2017 | Transitions into puberty and access to sexual and reproductive health information in two humanitarian settings: a cross-sectional survey of very young adolescents from Somalia and Myanmar | Confl Health | Kågesten, A. E.; Zimmerman, L.; Robinson, C.; Lee, C.; Bawoke, T.; Osman, S.; Schlecht, J. | Humanitarian settings | 10.1186/s13031-017-0127-8 |  |
| 1998 | Molecular and epidemiologic analysis of dengue virus isolates from Somalia | Emerg Infect Dis | Kanesa-thasan, N.; Chang, G. J.; Smoak, B. L.; Magill, A.; Burrous, M. J.; Hoke, C. H., Jr. | Adult | 10.3201/eid0402.980220 |  |
| 1994 | Dengue serotypes 2 and 3 in US forces in Somalia | Lancet | Kanesa-thasan, N.; Iacono-Connors, L.; Magill, A.; Smoak, B.; Vaughn, D.; Dubois, D.; Burrous, J.; Hoke, C. | Dengue/*microbiology | 10.1016/s0140-6736(94)92678-6 |  |
| 2019 | Frequency and patterns of second-line resistance conferring mutations among MDR-TB isolates resistant to a second-line drug from eSwatini, Somalia and Uganda (2014-2016) | BMC Pulm Med | Kateete, D. P.; Kamulegeya, R.; Kigozi, E.; Katabazi, F. A.; Lukoye, D.; Sebit, S. I.; Abdi, H.; Arube, P.; Kasule, G. W.; Musisi, K.; Dlamini, M. G.; Khumalo, D.; Joloba, M. L. | Amikacin/therapeutic use | 10.1186/s12890-019-0891-x |  |
| 2021 | Clinical characteristics of acute liver failure associated with hepatitis A infection in children in Mogadishu, Somalia: a hospital-based retrospective study | BMC Infect Dis | Keles, E.; Hassan-Kadle, M. A.; Osman, M. M.; Eker, H. H.; Abusoglu, Z.; Baydili, K. N.; Osman, A. M. | Child | 10.1186/s12879-021-06594-7 |  |
| 2022 | Review on camel production and marketing status in Ethiopia | Pastoralism | Kena, D. | Arid and semi-arid areas | 10.1186/s13570-022-00248-2 |  |
| 2022 | A multi-country, prospective cohort study to measure rate and risk of relapse among children recovered from severe acute malnutrition in Mali, Somalia, and South Sudan: a study protocol | BMC Nutr | King, S.; D'Mello-Guyett, L.; Yakowenko, E.; Riems, B.; Gallandat, K.; Mama Chabi, S.; Mohamud, F. A.; Ayoub, K.; Olad, A. H.; Aliou, B.; Marshak, A.; Trehan, I.; Cumming, O.; Stobaugh, H. | Community-based management of acute malnutrition | 10.1186/s40795-022-00576-x |  |
| 2017 | The migrant crisis comes to Minnesota: a dermatologist's perspective | Int J Dermatol | Knapp, A. P. | Adult | 10.1111/ijd.13787 |  |
| 2008 | Risk factors of visceral leishmaniasis in East Africa: a case-control study in Pokot territory of Kenya and Uganda | Int J Epidemiol | Kolaczinski, J. H.; Reithinger, R.; Worku, D. T.; Ocheng, A.; Kasimiro, J.; Kabatereine, N.; Brooker, S. | Adolescent | 10.1093/ije/dym275 |  |
| 2018 | Human and entomologic investigations of chikungunya outbreak in Mandera, Northeastern Kenya, 2016 | PLoS One | Konongoi, S. L.; Nyunja, A.; Ofula, V.; Owaka, S.; Koka, H.; Koskei, E.; Eyase, F.; Langat, D.; Mancuso, J.; Lutomiah, J.; Sang, R. | Antibodies, Viral/blood | 10.1371/journal.pone.0205058 |  |
| 1981 | Prevalence of Schistosoma haematobium in the Koryole and Merca Districts of the Somali Democratic Republic | Ann Trop Med Parasitol | Koura, M.; Upatham, E. S.; Awad, A. H.; Ahmed, M. D. | Adolescent | 10.1080/00034983.1981.11687408 |  |
| 2023 | Diagnostic capacity for invasive fungal infections in advanced HIV disease in Africa: a continent-wide survey | Lancet Infect Dis | Lakoh, S.; Kamudumuli, P. S.; Penney, R. O. S.; Haumba, S. M.; Jarvis, J. N.; Hassan, A. J.; Moudoute, N. L. E.; Ocansey, B. K.; Izco, S.; Kipkerich, S.; Sacarlal, J.; Awopeju, A. T.; Govender, N. P.; Munyanji, C. I. M.; Guyguy, K.; Orefuwa, E.; Denning, D. W. | Humans | 10.1016/s1473-3099(22)00656-9 |  |
| 2002 | 16-year-Old boy with gross hematuria | Mayo Clin Proc | Lischer, G. H.; Sweat, S. D. | Adolescent | 10.4065/77.5.475 |  |
| 2019 | Optimising age adjustment of trichiasis prevalence estimates using data from 162 standardised surveys from seven regions of Ethiopia | Ophthalmic Epidemiol | Macleod, C. K.; Porco, T. C.; Dejene, M.; Shafi, O.; Kebede, B.; Negussu, N.; Bero, B.; Taju, S.; Adamu, Y.; Negash, K.; Haileselassie, T.; Riang, J.; Badei, A.; Bakhtiari, A.; Willis, R.; Bailey, R. L.; Solomon, A. W. | Adolescent | 10.1080/09286586.2018.1555262 |  |
| 1989 | Pastoralists and hydatid disease: an ultrasound scanning prevalence survey in east Africa | Trans R Soc Trop Med Hyg | Macpherson, C. N.; Spoerry, A.; Zeyhle, E.; Romig, T.; Gorfe, M. | Africa, Eastern/epidemiology | 10.1016/0035-9203(89)90664-0 |  |
| 1962 | Key to the identification of east and central African freshwater snails of medical and veterinary importance | Bull World Health Organ | Mandahl-Barth, G. | Africa |  |  |
| 2003 | Emergence or re-emergence of visceral leishmaniasis in areas of Somalia, north-eastern Kenya, and south-eastern Ethiopia in 2000-01 | Trans R Soc Trop Med Hyg | Marlet, M. V.; Sang, D. K.; Ritmeijer, K.; Muga, R. O.; Onsongo, J.; Davidson, R. N. | Adolescent | 10.1016/s0035-9203(03)80012-3 |  |
| 2003 | A neglected disease of humans: a new focus of visceral leishmaniasis in Bakool, Somalia | Trans R Soc Trop Med Hyg | Marlet, M. V.; Wuillaume, F.; Jacquet, D.; Quispe, K. W.; Dujardin, J. C.; Boelaert, M. | Adolescent | 10.1016/s0035-9203(03)80099-8 |  |
| 2015 | Genetic diversity of Mycobacterium tuberculosis isolated from tuberculosis patients in the Serengeti ecosystem in Tanzania | Tuberculosis (Edinb) | Mbugi, E. V.; Katale, B. Z.; Siame, K. K.; Keyyu, J. D.; Kendall, S. L.; Dockrell, H. M.; Streicher, E. M.; Michel, A. L.; Rweyemamu, M. M.; Warren, R. M.; Matee, M. I.; van Helden, P. D. | Adult | 10.1016/j.tube.2014.11.006 |  |
| 2022 | Primary Spinal Glioblastoma Mimicking Neuroschistosomiasis: A Case Report | Cureus | McCallum, A. P.; Khattar, N. K.; Kolikonda, M. K.; Singla, S.; Alkhateeb, K. J.; Schaber, A. S.; Arnold, F. W.; Lippman, S. B.; Castillo, C. M.; Williams, B. J. | primary spinal tumor | 10.7759/cureus.30248 |  |
| 2023 | Global Burden of Cardiovascular Diseases and Risks, 1990-2022 | J Am Coll Cardiol | Mensah, G. A.; Fuster, V.; Murray, C. J. L.; Roth, G. A. | Humans | 10.1016/j.jacc.2023.11.007 |  |
| 2022 | Dengue Fever Outbreak Investigation in Werder Town, Dollo Zone, Somali Region, Ethiopia | Infect Drug Resist | Mesfin, Z.; Ali, A.; Abagero, A.; Asefa, Z. | Werder | 10.2147/idr.S368562 |  |
| 2000 | Malaria, intestinal parasites, and schistosomiasis among Barawan Somali refugees resettling to the United States: a strategy to reduce morbidity and decrease the risk of imported infections | Am J Trop Med Hyg | Miller, J. M.; Boyd, H. A.; Ostrowski, S. R.; Cookson, S. T.; Parise, M. E.; Gonzaga, P. S.; Addiss, D. G.; Wilson, M.; Nguyen-Dinh, P.; Wahlquist, S. P.; Weld, L. H.; Wainwright, R. B.; Gushulak, B. D.; Cetron, M. S. | Adolescent | 10.4269/ajtmh.2000.62.115 |  |
| 2022 | Spinal cord Schistosomiasis: A child's case with an unsatisfactory outcome that mimicked an intramedullary neoplasm. A rare case report | Ann Med Surg (Lond) | Mohamed, A. A.; Mengistu, M. G.; Haftu, H.; Mustefa, M.; Yusuf, A. A. | Necrosis | 10.1016/j.amsu.2022.104708 |  |
| 2022 | Acute kidney injury as initial presentation of visceral leishmaniasis in a young patient- A case report | Ann Med Surg (Lond) | Mohamed, A. H.; Bashir, A. M. | Acute kidney injury | 10.1016/j.amsu.2022.103821 |  |
| 1999 | Epidemics and public health in early colonial Somaliland | Soc Sci Med | Mohamed, J. | Colonialism/history | 10.1016/s0277-9536(98)00364-5 |  |
| 2024 | Epidemiological investigation of dengue fever outbreak and its socioeconomic determinants in Banadir region, Somalia | BMC Infect Dis | Mohamed, M. A.; Hassan, N. Y.; Osman, M. M.; Gedi, S.; Maalin, B. A. A.; Sultan, K. M.; Garba, B.; Osman, A. A.; Osman, A. Y.; Ahmed, A. D. | Child | 10.1186/s12879-024-09276-2 |  |
| 2018 | Haemoptysis and fever in a young refugee from Somalia | Int J Infect Dis | Mondoni, M.; Viganò, O.; Ferrarese, M.; Bimbatti, M.; Cavallini, M.; Codecasa, L.; D'Arminio Monforte, A.; Bulfamante, G.; Centanni, S.; Sotgiu, G. | Bronchoscopy | 10.1016/j.ijid.2018.10.002 |  |
| 2021 | Brucella Seroprevalence and Associated Risk Factors in Occupationally Exposed Humans in Selected Districts of Southern Province, Zambia | Front Public Health | Mubanga, M.; Mfune, R. L.; Kothowa, J.; Mohamud, A. S.; Chanda, C.; McGiven, J.; Bumbangi, F. N.; Hang'ombe, B. M.; Godfroid, J.; Simuunza, M.; Muma, J. B. | Adolescent | 10.3389/fpubh.2021.745244 |  |
| 2004 | Schistosomiasis--an unusual cause of ureteral obstruction: a case history and perspective | Clin Med Res | Neal, P. M. | Adult | 10.3121/cmr.2.4.216 |  |
| 2023 | Developing a hierarchical framework for assessing the strategic effectiveness of sustainable waste management in the Somaliland construction industry | Environ Sci Pollut Res Int | Negash, Y. T.; Hassan, A. M.; Tseng, M. L.; Ali, M. H.; Lim, M. K. | *Construction Industry | 10.1007/s11356-023-27060-8 |  |
| 2013 | Prevalence and distribution of schistosomiasis in afder and gode zone of somali region, ethiopia | J Glob Infect Dis | Negussu, N.; Wali, M.; Ejigu, M.; Debebe, F.; Aden, S.; Abdi, R.; Mohamed, Y.; Deribew, A.; Deribe, K. | S. haematobium | 10.4103/0974-777x.122007 |  |
| 2023 | National and Regional Fraction of Cancer Incidence and Death Attributable to Current Tobacco and Water-Pipe Smoking in the Eastern Mediterranean Countries in 2020 | Nicotine Tob Res | Nemati, S.; Naji, P.; Abdi, S.; Lotfi, F.; Saeedi, E.; Mehravar, S. A.; Fattahi, P.; Sheikh, M.; Vand Rajabpour, M.; Eftekharzadeh, A.; Zendehdel, K. | Adult | 10.1093/ntr/ntac179 |  |
| 2020 | Prevalence and pattern of waterborne parasitic infections in eastern Africa: A systematic scoping review | Food Waterborne Parasitol | Ngowi, H. A. | Burden | 10.1016/j.fawpar.2020.e00089 |  |
| 2009 | Visor flap for total upper and lower lip reconstruction: a case report | J Med Case Rep | Nthumba, P.; Carter, L. |  | 10.4076/1752-1947-3-7312 |  |
| 1999 | IgM antibodies in hospitalized children with febrile illness during an inter-epidemic period of measles, in Somalia | J Clin Virol | Nur, Y. A.; Groen, J.; Yusuf, M. A.; Osterhaus, A. D. | Antibodies, Bacterial/*blood | 10.1016/s1386-6532(98)00002-x |  |
| 1979 | IgE serum levels in urinary schistosomiasis | Tropenmed Parasitol | Nuti, M.; Rasi, G.; Simoni, L.; Bonini, S. | Humans |  |  |
| 1979 | Leprosy and hepatitis B virus markers: incidence of HBsAg and HBeAg in Somalian patients | Int J Lepr Other Mycobact Dis | Nuti, M.; Tarabini, G.; Palermo, P.; Tarabini, G. L.; Thamer, G. | Adolescent |  |  |
| 2018 | Investigation of laboratory confirmed Dengue outbreak in North-eastern Kenya, 2011 | PLoS One | Obonyo, M.; Fidhow, A.; Ofula, V. | Adolescent | 10.1371/journal.pone.0198556 |  |
| 2023 | Clinical Characteristics of Acute Kidney Injury Associated with Tropical Acute Febrile Illness | Trop Med Infect Dis | Omar, F. D.; Phumratanaprapin, W.; Silachamroon, U.; Hanboonkunupakarn, B.; Sriboonvorakul, N.; Thaipadungpanit, J.; Pan-Ngum, W. | acute febrile illness | 10.3390/tropicalmed8030147 |  |
| 1986 | Yaws assessment in Somalia | Southeast Asian J Trop Med Public Health | Omar, M. A. | Health Status |  |  |
| 2017 | Understanding the unique experiences, perspectives and sexual and reproductive health needs of very young adolescents: Somali refugees in Ethiopia | Confl Health | Ortiz-Echevarria, L.; Greeley, M.; Bawoke, T.; Zimmerman, L.; Robinson, C.; Schlecht, J. | Adolescence | 10.1186/s13031-017-0129-6 |  |
| 2021 | Integrated community based human and animal syndromic surveillance in Adadle district of the Somali region of Ethiopia | One Health | Osman, Y.; Ali, S. M.; Schelling, E.; Tschopp, R.; Hattendorf, J.; Muhumed, A.; Zinsstag, J. | Community based | 10.1016/j.onehlt.2021.100334 |  |
| 2024 | Atypical perforated appendicitis secondary to schistosomiasis: a rare case report | Ann Med Surg (Lond) | Osoble Osman, F. A.; Mohamed, Y. G.; Salad, N. M.; Yahye, N. M. | appendicitis | 10.1097/ms9.0000000000001480 |  |
| 2016 | Louse-borne relapsing fever - report of four cases in Switzerland, June-December 2015 | BMC Infect Dis | Osthoff, M.; Schibli, A.; Fadini, D.; Lardelli, P.; Goldenberger, D. | Adolescent | 10.1186/s12879-016-1541-z |  |
| 2001 | Consider schistosomiasis | Minn Med | Park, M. A.; Mueller, P. S. | Adolescent |  |  |
| 2007 | High prevalence and presumptive treatment of schistosomiasis and strongyloidiasis among African refugees | Clin Infect Dis | Posey, D. L.; Blackburn, B. G.; Weinberg, M.; Flagg, E. W.; Ortega, L.; Wilson, M.; Secor, W. E.; Sanders-Lewis, K.; Won, K.; Maguire, J. H. | Abdominal Pain/etiology | 10.1086/522529 |  |
| 2010 | Leishmaniasis in the World Health Organization Eastern Mediterranean Region | Int J Antimicrob Agents | Postigo, J. A. | Animals | 10.1016/j.ijantimicag.2010.06.023 |  |
| 2007 | Epidemiology and clinical features of patients with visceral leishmaniasis treated by an MSF clinic in Bakool region, Somalia, 2004-2006 | PLoS Negl Trop Dis | Raguenaud, M. E.; Jansson, A.; Vanlerberghe, V.; Deborggraeve, S.; Dujardin, J. C.; Orfanos, G.; Reid, T.; Boelaert, M. | Adolescent | 10.1371/journal.pntd.0000085 |  |
| 2021 | Correction: Epidemiology and Clinical Features of Patients with Visceral Leishmaniasis Treated by an MSF Clinic in Bakool Region, Somalia, 2004-2006 | PLoS Negl Trop Dis | Raguenaud, M. E.; Jansson, A.; Vanlerberghe, V.; Van der Auwera, G.; Deborggraeve, S.; Dujardin, J. C.; Orfanos, I.; Reid, T.; Boelaert, M. |  | 10.1371/journal.pntd.0009356 |  |
| 2020 | Progress towards elimination of lymphatic filariasis in the Eastern Mediterranean Region | Int Health | Ramzy, R. M. R.; Al Kubati, A. S. | Egypt | 10.1093/inthealth/ihaa037 |  |
| 2014 | Near death of a pregnant Somali woman due to neglected eclampsia | Clin Exp Obstet Gynecol | Rouzi, A. A.; Almrstani, A. M. | Adolescent |  |  |
| 2022 | Do Spiders Ride on the Fear of Scorpions? A Cross-Cultural Eye Tracking Study | Animals (Basel) | Rudolfová, V.; Štolhoferová, I.; Elmi, H. S. A.; Rádlová, S.; Rexová, K.; Berti, D. A.; Král, D.; Sommer, D.; Landová, E.; Frýdlová, P.; Frynta, D. | Africa | 10.3390/ani12243466 |  |
| 2021 | The Search for Putative Hits in Combating Leishmaniasis: The Contributions of Natural Products Over the Last Decade | Nat Prod Bioprospect | Sakyi, P. O.; Amewu, R. K.; Devine, Rnoa; Ismaila, E.; Miller, W. A.; Kwofie, S. K. | Chemoinformatics | 10.1007/s13659-021-00311-2 |  |
| 2022 | Substance use, affective symptoms, and suicidal ideation among Russian, Somali, and Kurdish migrants in Finland | Transcult Psychiatry | Salama, E.; Castaneda, A. E.; Suvisaari, J.; Rask, S.; Laatikainen, T.; Niemelä, S. | Affective Symptoms | 10.1177/1363461520906028 |  |
| 2013 | Spatially explicit Schistosoma infection risk in eastern Africa using Bayesian geostatistical modelling | Acta Trop | Schur, N.; Hürlimann, E.; Stensgaard, A. S.; Chimfwembe, K.; Mushinge, G.; Simoonga, C.; Kabatereine, N. B.; Kristensen, T. K.; Utzinger, J.; Vounatsou, P. | Adolescent | 10.1016/j.actatropica.2011.10.006 |  |
| 1991 | Low prevalence of human immunodeficiency virus-1 (HIV-1), HIV-2, and human T cell lymphotropic virus-1 infection in Somalia | Am J Trop Med Hyg | Scott, D. A.; Corwin, A. L.; Constantine, N. T.; Omar, M. A.; Guled, A.; Yusef, M.; Roberts, C. R.; Watts, D. M. | Adolescent | 10.4269/ajtmh.1991.45.653 |  |
| 2016 | HIV/AIDS among pastoralists and refugees in north-east Africa: a neglected problem | Afr J AIDS Res | Serbessa, M. K.; Mariam, D. H.; Kassa, A.; Alwan, F.; Kloos, H. | Acquired Immunodeficiency Syndrome/*epidemiology/prevention & control | 10.2989/16085906.2016.1148060 |  |
| 2014 | Case report: Non-invasive management of Madura foot with oral posaconazole and ciprofloxacin | Am J Trop Med Hyg | Sharma, A. M.; Sharma, N.; Nat, A.; Rane, M.; Endy, T. P. | Administration, Oral | 10.4269/ajtmh.14-0335 |  |
| 1995 | Illness in Journalists and Relief Workers Involved in International Humanitarian Assistance Efforts in Somalia, 1992-93 | J Travel Med | Sharp, T. W.; DeFraites, R. F.; Thornton, S. A.; Burans, J. P.; Wallace, M. R. |  | 10.1111/j.1708-8305.1995.tb00630.x |  |
| 1995 | Dengue fever in U.S. troops during Operation Restore Hope, Somalia, 1992-1993 | Am J Trop Med Hyg | Sharp, T. W.; Wallace, M. R.; Hayes, C. G.; Sanchez, J. L.; DeFraites, R. F.; Arthur, R. R.; Thornton, S. A.; Batchelor, R. A.; Rozmajzl, P. J.; Hanson, R. K.; et al. | Adult |  |  |
| 1999 | Where health care has no access: the nomadic populations of sub-Saharan Africa | Trop Med Int Health | Sheik-Mohamed, A.; Velema, J. P. | Adult | 10.1046/j.1365-3156.1999.00473.x |  |
| 1995 | Visceral leishmaniasis in Somalia: prevalence of markers of infection and disease manifestations in a village in an endemic area | Trans R Soc Trop Med Hyg | Shiddo, S. A.; Aden Mohamed, A.; Akuffo, H. O.; Mohamud, K. A.; Herzi, A. A.; Herzi Mohamed, H.; Huldt, G.; Nilsson, L. A.; Ouchterlony, O.; Thorstensson, R. | Adolescent | 10.1016/0035-9203(95)90008-x |  |
| 1995 | Visceral leishmaniasis in Somalia: prevalence of leishmanin-positive and seropositive inhabitants in an endemic area | Trans R Soc Trop Med Hyg | Shiddo, S. A.; Akuffo, H. O.; Mohamed, A. A.; Huldt, G.; Nilsson, L. A.; Ouchterlony, O.; Thorstensson, R. | Adolescent | 10.1016/0035-9203(95)90640-1 |  |
| 1994 | Reference ranges for IgG, IgM and IgA in the serum of urban and rural Somalis | Trop Geogr Med | Shiddo, S. A.; Huldt, G.; Jama, H.; Nilsson, L. A.; Ouchterlony, O.; Warsame, M.; Jonsson, J. | Adolescent |  |  |
| 1996 | Visceral leishmaniasis in Somalia. Significance of IgG subclasses and of IgE response | Immunol Lett | Shiddo, S. A.; Huldt, G.; Nilsson, L. A.; Ouchterlony, O.; Thorstensson, R. | Animals | 10.1016/0165-2478(96)02529-1 |  |
| 1995 | Visceral leishmaniasis in Somalia. Circulating antibodies as measured by DAT, immunofluorescence and ELISA | Trop Geogr Med | Shiddo, S. A.; Mohamed, A. A.; Huldt, G.; Loftenius, A.; Nilsson, L.; Jonsson, J.; Ouchterlony, O.; Thorstensson, R. | Adolescent |  |  |
| 1999 | Partial nucleotide sequencing and molecular evolution of epidemic causing Dengue 2 strains | J Infect Dis | Singh, U. B.; Maitra, A.; Broor, S.; Rai, A.; Pasha, S. T.; Seth, P. | Base Sequence | 10.1086/315043 |  |
| 1978 | Schistosoma haematobium in the Wabi Shebelle Valley of Ethiopia | Am J Trop Med Hyg | Sole, G. D.; Lemma, A.; Mazengia, B. | Adolescent | 10.4269/ajtmh.1978.27.928 |  |
| 2020 | Banat donkey, a neglected donkey breed from the central Balkans (Serbia) | PeerJ | Stanisic, L.; Aleksić, J. M.; Dimitrijevic, V.; Kovačević, B.; Stevanovic, J.; Stanimirovic, Z. | Conservation of genetic resources | 10.7717/peerj.8598 |  |
| 2023 | Gender differences in the surgical management of trachomatous trichiasis: an exploratory analysis of global trachoma survey data, 2015-2019 | Int Health | Sullivan, K. M.; Harding-Esch, E. M.; Batcho, W. E.; Issifou, A. A. B.; Lopes, M. F. C.; Szwarcwald, C. L.; Vaz Ferreira Gomez, D.; Bougouma, C.; Christophe, N.; Kabore, M.; Bucumi, V.; Bella, A. L.; Epee, E.; Yaya, G.; Trujillo-Trujillo, J.; Dejene, M.; Gebretsadik, F. S.; Gebru, G.; Kebede, F.;... | Humans | 10.1093/inthealth/ihad067 |  |
| 2006 | Hematuria in children due to schistosomiasis in a nonendemic setting | Clin Pediatr (Phila) | Summer, A. P.; Stauffer, W.; Maroushek, S. R.; Nevins, T. E. | Adolescent | 10.1177/000992280604500210 |  |
| 2017 | Visceral leishmaniasis in Somalia: A review of epidemiology and access to care | PLoS Negl Trop Dis | Sunyoto, T.; Potet, J.; Boelaert, M. | *Health Services Accessibility | 10.1371/journal.pntd.0005231 |  |
| 2019 | Exploring global and country-level barriers to an effective supply of leishmaniasis medicines and diagnostics in eastern Africa: a qualitative study | BMJ Open | Sunyoto, T.; Potet, J.; den Boer, M.; Ritmeijer, K.; Postigo, J. A. R.; Ravinetto, R.; Alves, F.; Picado, A.; Boelaert, M. | Agglutination Tests/*statistics & numerical data | 10.1136/bmjopen-2019-029141 |  |
| 2021 | Determinants of Postnatal Care Check-ups in Ethiopia: A Multi-Level Analysis | Ethiop J Health Sci | Tadele, A.; Getinet, M. | Cesarean Section | 10.4314/ejhs.v31i4.9 |  |
| 2022 | Spatial variation and associated factors of deworming among children aged 24 to 59 months in Ethiopia: spatial and multilevel logistic analysis | BMC Public Health | Tareke, A. A. | Child | 10.1186/s12889-022-13156-2 |  |
| 2008 | An update review on Commiphora molmol and related species | J Egypt Soc Parasitol | Tonkal, A. M.; Morsy, T. A. | Anti-Infective Agents/therapeutic use |  |  |
| 2010 | Infection due to a novel mycobacterium, mimicking multidrug-resistant Mycobacterium tuberculosis | Clin Microbiol Infect | Tortoli, E.; Rogasi, P. G.; Fantoni, E.; Beltrami, C.; De Francisci, A.; Mariottini, A. | Anti-Bacterial Agents/pharmacology/therapeutic use | 10.1111/j.1469-0691.2009.03063.x |  |
| 2022 | Risk factors for Brucellosis and knowledge-attitude practice among pastoralists in Afar and Somali regions of Ethiopia | Prev Vet Med | Tschopp, R.; GebreGiorgis, A.; Abdulkadir, O.; Molla, W.; Hamid, M.; Tassachew, Y.; Andualem, H.; Osman, M.; Waqjira, M. W.; Mohammed, A.; Negron, M.; Walke, H.; Kadzik, M.; Mamo, G. | Abortion, Veterinary | 10.1016/j.prevetmed.2021.105557 |  |
| 2021 | Integrated human-animal sero-surveillance of Brucellosis in the pastoral Afar and Somali regions of Ethiopia | PLoS Negl Trop Dis | Tschopp, R.; Gebregiorgis, A.; Tassachew, Y.; Andualem, H.; Osman, M.; Waqjira, M. W.; Hattendorf, J.; Mohammed, A.; Hamid, M.; Molla, W.; Mitiku, S. A.; Walke, H.; Negron, M.; Kadzik, M.; Mamo, G. | Adult | 10.1371/journal.pntd.0009593 |  |
| 1981 | Studies on the transmission of Schistosoma haematobium and the bionomics of Bulinus (Ph.) abyssinicus in the Somali Democratic Republic | Ann Trop Med Parasitol | Upatham, E. S.; Koura, M.; Ahmed, M. D.; Awad, A. H. | Animals | 10.1080/00034983.1981.11687409 |  |
| 2020 | Molecular surveillance of Pfcrt and k13 propeller polymorphisms of imported Plasmodium falciparum cases to Zhejiang Province, China between 2016 and 2018 | Malar J | Wang, X.; Ruan, W.; Zhou, S.; Huang, F.; Lu, Q.; Feng, X.; Yan, H. | Adolescent | 10.1186/s12936-020-3140-0 |  |
| 2016 | Prioritization in Somali health system strengthening: a qualitative study | Int Health | Warsame, A.; Handuleh, J.; Patel, P. | Delivery of Health Care/*organization & administration | 10.1093/inthealth/ihv060 |  |
| 1995 | A recent case of visceral leishmaniasis in Somalia | Ann Trop Med Parasitol | Woolhead, A. | Adult | 10.1080/00034983.1995.11813003 |  |
| 2024 | Breeding habitats, bionomics and phylogenetic analysis of Aedes aegypti and first detection of Culiseta longiareolata, and Ae. hirsutus in Somali Region, eastern Ethiopia | PLoS One | Yared, S.; Gebressilasie, A.; Worku, A.; Mohammed, A.; Gunarathna, I.; Rajamanickam, D.; Waymire, E.; Balkew, M.; Carter, T. E. | Male | 10.1371/journal.pone.0296406 |  |
| 2016 | Recent Chikungunya Virus Infection in 2 Travelers Returning from Mogadishu, Somalia, to Italy, 2016 | Emerg Infect Dis | Zammarchi, L.; Fortuna, C.; Venturi, G.; Rinaldi, F.; Capobianco, T.; Remoli, M. E.; Rossolini, G. M.; Rezza, G.; Bartoloni, A. | Chikungunya Fever/epidemiology/*transmission/*virology | 10.3201/eid2211.161225 |  |
| 2023 | Eumycetoma caused by Biatriospora mackinnonii in a young pregnant woman from Somalia | Int J Infect Dis | Zimmer, F.; Kellner, N.; Nenoff, P.; Lübbert, C. | Female | 10.1016/j.ijid.2022.12.021 |  |
| 2022 | Mito-phylogenetic relationship of the new subspecies of gentle monkey Cercopithecus mitis manyaraensis, Butynski & De Jong, 2020 | Primate Biol | Zinner, D.; Knauf, S.; Chuma, I. S.; Butynski, T. M.; De Jong, Y. A.; Keyyu, J. D.; Kaitila, R.; Roos, C. |  | 10.5194/pb-9-11-2022 |  |
| 1957 | Sandflies & onchocerciasis in Somaliland | Bull World Health Organ | Zuretti, S. | Animals |  |  |

#### Database: CINAHL

Total records retrieved: 33

| Year | Title | Journal | Authors | Keywords | DOI | URL |
| --- | --- | --- | --- | --- | --- | --- |
| 2013 | Case Study | Urologic Nursing |  | Hematuria -- Etiology |  | https://lstmed.idm.oclc.org/login?url=https://search.ebscohost.com/login.aspx?direct=true&AuthType=sso&db=ccm&AN=89676029&site=ehost-live&scope=site |
| 2016 | Global, regional, and national incidence, prevalence, and years lived with disability for 310 diseases and injuries, 1990-2015: a systematic analysis for the Global Burden of Disease Study 2015 | Lancet |  | Persons with Disabilities | 10.1016/S0140-6736(16)31678-6 | https://lstmed.idm.oclc.org/login?url=https://search.ebscohost.com/login.aspx?direct=true&AuthType=sso&db=ccm&AN=119386597&site=ehost-live&scope=site |
| 2017 | Control of visceral leishmaniasis in Somalia: achievements in a challenging scenario, 2013-2015 | Weekly Epidemiological Record |  | Leishmaniasis, Visceral -- Epidemiology -- Somalia |  | https://lstmed.idm.oclc.org/login?url=https://search.ebscohost.com/login.aspx?direct=true&AuthType=sso&db=ccm&AN=125308128&site=ehost-live&scope=site |
| 2020 | Hydatid cyst of the foot: a case report | Journal of Medical Case Reports |  | Echinococcosis -- Diagnosis | 10.1186/s13256-019-2337-8 | https://lstmed.idm.oclc.org/login?url=https://search.ebscohost.com/login.aspx?direct=true&AuthType=sso&db=ccm&AN=141132431&site=ehost-live&scope=site |
| 2022 | WHE1 acute emergencies monthly summary -- July and August 2022 | Weekly Epidemiological Record |  | World Health |  | https://lstmed.idm.oclc.org/login?url=https://search.ebscohost.com/login.aspx?direct=true&AuthType=sso&db=ccm&AN=158882534&site=ehost-live&scope=site |
| 2020 | Visceral Leishmaniasis, Northern Somalia, 2013-2019 | Emerging Infectious Diseases | Aalto, Mikko K.; Temmy, Sunyoto; Yusuf, Mohamed Ahmed Ali; Mohamed, Abdiaziz Ahmed; Van der Auwera, Gert; Dujardin, Jean-Claude; Sunyoto, Temmy | Leishmaniasis -- Epidemiology | 10.3201/eid2601.181851 | https://lstmed.idm.oclc.org/login?url=https://search.ebscohost.com/login.aspx?direct=true&AuthType=sso&db=ccm&AN=140950294&site=ehost-live&scope=site |
| 2020 | Prevalence of refractive error and visual impairment among school-age children of Hargesia, Somaliland, Somalia | Eastern Mediterranean Health Journal | Ahmed, Zahra Abdi; Alrasheed, Saif Hassan; Alghamdi, Waleed | Refractive Errors -- Epidemiology -- Djibouti | 10.26719/emhj.20.077 | https://lstmed.idm.oclc.org/login?url=https://search.ebscohost.com/login.aspx?direct=true&AuthType=sso&db=ccm&AN=147141634&site=ehost-live&scope=site |
| 2022 | Somalia tackles leprosy and visceral leishmaniasis | Lancet Infectious Diseases | Bagcchi, Sanjeet | Hansen's Disease -- Complications | 10.1016/S1473-3099(22)00168-2 | https://lstmed.idm.oclc.org/login?url=https://search.ebscohost.com/login.aspx?direct=true&AuthType=sso&db=ccm&AN=155886802&site=ehost-live&scope=site |
| 2007 | Tuberculosis in complex emergencies | Bulletin of the World Health Organization | Coninx, R. | Antitubercular Agents -- Therapeutic Use | 10.2471/blt.06.037630 | https://lstmed.idm.oclc.org/login?url=https://search.ebscohost.com/login.aspx?direct=true&AuthType=sso&db=ccm&AN=106008151&site=ehost-live&scope=site |
| 2024 | Mycetoma case series in Somalia | Tropical Doctor | Doğan, Ahmet; Ali, Ahmed Mohamed; Ali, Mukhtar Abdullahi; Abdullahi, İsmail Mohamoud |  | 10.1177/00494755231201664 | https://lstmed.idm.oclc.org/login?url=https://search.ebscohost.com/login.aspx?direct=true&AuthType=sso&db=ccm&AN=174794453&site=ehost-live&scope=site |
| 2012 | Dengue and US military operations from the Spanish-American War through today | Emerging Infectious Diseases | Gibbons, R. V.; Streitz, M.; Babina, T.; Fried, J. R.; Gibbons, Robert V.; Streitz, Matthew; Babina, Tatyana; Fried, Jessica R. | Dengue -- History | 10.3201/eid1804.110134 | https://lstmed.idm.oclc.org/login?url=https://search.ebscohost.com/login.aspx?direct=true&AuthType=sso&db=ccm&AN=104546548&site=ehost-live&scope=site |
| 2015 | Exploring Somali women’s reproductive health knowledge and experiences: results from focus group discussions in Mogadishu | Reproductive Health Matters | Gure, Faduma; Yusuf, Marian; Foster, Angel M. | Reproductive Health | 10.1016/j.rhm.2015.11.018 | https://lstmed.idm.oclc.org/login?url=https://search.ebscohost.com/login.aspx?direct=true&AuthType=sso&db=ccm&AN=111893128&site=ehost-live&scope=site |
| 2021 | Spectrum and Prevalence of Thyroid Diseases at a Tertiary Referral Hospital in Mogadishu, Somalia: A Retrospective Study of 976 Cases | International Journal of Endocrinology | Hassan-Kadle, Mohamed A.; Adani, Abdulkamil Abdullahi; Eker, Hasan Huseyin; Keles, Esra; Muse Osman, Marian; Mahdi Ahmed, Hussein; Görçin Karaketir, Şeyma |  | 10.1155/2021/7154250 | https://lstmed.idm.oclc.org/login?url=https://search.ebscohost.com/login.aspx?direct=true&AuthType=sso&db=ccm&AN=154309963&site=ehost-live&scope=site |
| 2009 | Appendectomy to remember | Journal of Travel Medicine | Jama, S.; Manivel, J. C.; Abd Alla, M. D.; Stauffer, W. M. | Appendicitis | 10.1111/j.1708-8305.2009.00306.x | https://lstmed.idm.oclc.org/login?url=https://search.ebscohost.com/login.aspx?direct=true&AuthType=sso&db=ccm&AN=105222532&site=ehost-live&scope=site |
| 2024 | Retrospective study on the dengue fever outbreak in Puntland State, Somalia | BMC Infectious Diseases | Jama, Saaid Said; Abshir, Said Nuriye; Jama, Jibril Said; Abdi, Mohamed Mohamud |  | 10.1186/s12879-024-09552-1 | https://lstmed.idm.oclc.org/login?url=https://search.ebscohost.com/login.aspx?direct=true&AuthType=sso&db=ccm&AN=178621402&site=ehost-live&scope=site |
| 2021 | Clinical characteristics of acute liver failure associated with hepatitis A infection in children in Mogadishu, Somalia: a hospital-based retrospective study | BMC Infectious Diseases | Keles, Esra; Hassan-Kadle, Mohamed A.; Osman, Marian Muse; Eker, Hasan Huseyin; Abusoglu, Zeynep; Baydili, Kursad Nuri; Osman, Aamir Muse | Liver Failure, Acute -- Epidemiology | 10.1186/s12879-021-06594-7 | https://lstmed.idm.oclc.org/login?url=https://search.ebscohost.com/login.aspx?direct=true&AuthType=sso&db=ccm&AN=152168241&site=ehost-live&scope=site |
| 2005 | Trematodes in two travellers | Journal of the Royal Society of Medicine | Kumari, S. B.; Allan, P. S.; Kumari, S. B.; Allan, P. S. | Schistosomiasis | 10.1258/jrsm.98.1.25 | https://lstmed.idm.oclc.org/login?url=https://search.ebscohost.com/login.aspx?direct=true&AuthType=sso&db=ccm&AN=106586659&site=ehost-live&scope=site |
| 2018 | Mycetoma in Middle East, A 112 Years Review...5th Iranian Congress in Medical Mycology, Tehran, Iran, Dec 4-6, 2018 | Current Medical Mycology | Mahmoudabadi, Ali Zarei; Taghipour, Simin; Gharaghani, Maral; Kiasat, Neda | Mycoses -- Epidemiology -- Middle East | 10.18502/cmm.4.S1.2018.178 | https://lstmed.idm.oclc.org/login?url=https://search.ebscohost.com/login.aspx?direct=true&AuthType=sso&db=ccm&AN=133659351&site=ehost-live&scope=site |
| 2018 | Mycetoma in Middle East, A 112 Years Review...5th Iranian Congress in Medical Mycology, Tehran, Iran, Dec 4-6, 2018 | Current Medical Mycology | Mahmoudabadi, Ali Zarei; Taghipour, Simin; Gharaghani, Maral; Kiasat, Neda | Mycoses -- Epidemiology -- Middle East | 10.18502/cmm.4.S1.2018.178 | https://lstmed.idm.oclc.org/login?url=https://search.ebscohost.com/login.aspx?direct=true&AuthType=sso&db=ccm&AN=133659351&site=ehost-live&scope=site |
| 2018 | Infectious and dermatological diseases among arriving migrants on the Italian coasts | European Journal of Public Health | Meco, Eugenia Di; Napoli, Anteo Di; Amato, Loredana Maria; Fortino, Antonio; Costanzo, Gianfranco; Rossi, Alessandra; Mirisola, Concetta; Petrelli, Alessio; Team, The Inmp | Communicable Diseases -- Diagnosis | 10.1093/eurpub/cky126 | https://lstmed.idm.oclc.org/login?url=https://search.ebscohost.com/login.aspx?direct=true&AuthType=sso&db=ccm&AN=131920514&site=ehost-live&scope=site |
| 2018 | Infectious and dermatological diseases among arriving migrants on the Italian coasts | European Journal of Public Health | Meco, Eugenia Di; Napoli, Anteo Di; Amato, Loredana Maria; Fortino, Antonio; Costanzo, Gianfranco; Rossi, Alessandra; Mirisola, Concetta; Petrelli, Alessio; Team, The Inmp | Communicable Diseases -- Diagnosis | 10.1093/eurpub/cky126 | https://lstmed.idm.oclc.org/login?url=https://search.ebscohost.com/login.aspx?direct=true&AuthType=sso&db=ccm&AN=131920475&site=ehost-live&scope=site |
| 1999 | Epidemics and public health in early colonial Somaliland | Social Science & Medicine | Mohamed, J. | Disease Outbreaks -- History -- Somalia | 10.1016/s0277-9536(98)00364-5 | https://lstmed.idm.oclc.org/login?url=https://search.ebscohost.com/login.aspx?direct=true&AuthType=sso&db=ccm&AN=107115102&site=ehost-live&scope=site |
| 2024 | Epidemiological investigation of dengue fever outbreak and its socioeconomic determinants in Banadir region, Somalia | BMC Infectious Diseases | Mohamed, Mohamed Abdelrahman; Hassan, Nuralein Yusuf; Osman, Marian Muse; Gedi, Saido; Maalin, Bisma Abdullahi Ali; Sultan, Kasim Mahdi; Garba, Bashiru; Osman, Ali Abdirahman; Osman, Abdinasir Yusuf; Ahmed, Abdifatah Diriye |  | 10.1186/s12879-024-09276-2 | https://lstmed.idm.oclc.org/login?url=https://search.ebscohost.com/login.aspx?direct=true&AuthType=sso&db=ccm&AN=176562327&site=ehost-live&scope=site |
| 2004 | Schistosomiasis -- an unusual cause of ureteral obstruction: a case history and perspective | Clinical Medicine & Research | Neal, P. M. | Schistosomiasis -- Complications | 10.3121/cmr.2.4.216 | https://lstmed.idm.oclc.org/login?url=https://search.ebscohost.com/login.aspx?direct=true&AuthType=sso&db=ccm&AN=106489952&site=ehost-live&scope=site |
| 2023 | National and Regional Fraction of Cancer Incidence and Death Attributable to Current Tobacco and Water-Pipe Smoking in the Eastern Mediterranean Countries in 2020 | Nicotine & Tobacco Research | Nemati, Saeed; Naji, Parnian; Abdi, Sepideh; Lotfi, Fereshte; Saeedi, Elnaz; Mehravar, Sepideh A.; Fattahi, Pedram; Sheikh, Mahdi; Rajabpour, Mojtaba Vand; Eftekharzadeh, Anita; Zendehdel, Kazem; Vand Rajabpour, Mojtaba | Neoplasms -- Epidemiology | 10.1093/ntr/ntac179 | https://lstmed.idm.oclc.org/login?url=https://search.ebscohost.com/login.aspx?direct=true&AuthType=sso&db=ccm&AN=160560553&site=ehost-live&scope=site |
| 2016 | Louse-borne relapsing fever - report of four cases in Switzerland, June-December 2015 | BMC Infectious Diseases | Osthoff, Michael; Schibli, Adrian; Fadini, Davide; Lardelli, Pietro; Goldenberger, Daniel | Lice Infestations -- Microbiology | 10.1186/s12879-016-1541-z | https://lstmed.idm.oclc.org/login?url=https://search.ebscohost.com/login.aspx?direct=true&AuthType=sso&db=ccm&AN=115435710&site=ehost-live&scope=site |
| 2011 | Clinical issues in refugee healthcare : The Somali Bantu population | Nurse Practitioner | Parve, Julie; Kaul, Teri | Cultural Competence | 10.1097/01.NPR.0000398777.52008.3f | https://lstmed.idm.oclc.org/login?url=https://search.ebscohost.com/login.aspx?direct=true&AuthType=sso&db=ccm&AN=108244483&site=ehost-live&scope=site |
| 2022 | Substance use, affective symptoms, and suicidal ideation among Russian, Somali, and Kurdish migrants in Finland | Transcultural Psychiatry | Salama, Essi; Castaneda, Anu E.; Suvisaari, Jaana; Rask, Shadia; Laatikainen, Tiina; Niemelä, Solja | Migrants -- Psychosocial Factors -- Finland | 10.1177/1363461520906028 | https://lstmed.idm.oclc.org/login?url=https://search.ebscohost.com/login.aspx?direct=true&AuthType=sso&db=ccm&AN=155344933&site=ehost-live&scope=site |
| 2016 | HIV/AIDS among pastoralists and refugees in north-east Africa: a neglected problem | African Journal of AIDS Research (AJAR) | Serbessa, Mirgissa Kaba; Mariam, Damen Haile; Kassa, Afework; Alwan, Fathia; Kloos, Helmut | HIV Infections -- Epidemiology -- Africa, Eastern | 10.2989/16085906.2016.1148060 | https://lstmed.idm.oclc.org/login?url=https://search.ebscohost.com/login.aspx?direct=true&AuthType=sso&db=ccm&AN=114016402&site=ehost-live&scope=site |
| 2006 | Hematuria in children due to schistosomiasis in a nonendemic setting | Clinical Pediatrics | Summer, A. P.; Stauffer, W.; Maroushek, S. R.; Nevins, T. E. | Hematuria -- Etiology | 10.1177/000992280604500210 | https://lstmed.idm.oclc.org/login?url=https://search.ebscohost.com/login.aspx?direct=true&AuthType=sso&db=ccm&AN=106103818&site=ehost-live&scope=site |
| 2022 | Spatial variation and associated factors of deworming among children aged 24 to 59 months in Ethiopia: spatial and multilevel logistic analysis | BMC Public Health | Tareke, Abiyu Abadi | Diarrhea | 10.1186/s12889-022-13156-2 | https://lstmed.idm.oclc.org/login?url=https://search.ebscohost.com/login.aspx?direct=true&AuthType=sso&db=ccm&AN=156219838&site=ehost-live&scope=site |
| 2016 | Recent Chikungunya Virus Infection in 2 Travelers Returning from Mogadishu, Somalia, to Italy, 2016 | Emerging Infectious Diseases | Zammarchi, Lorenzo; Fortuna, Claudia; Venturi, Giulietta; Rinaldi, Francesca; Capobianco, Teresa; Remoli, Maria Elena; Rossolini, Gian Maria; Rezza, Giovanni; Bartoloni, Alessandro |  | 10.3201/eid2211.161225 | https://lstmed.idm.oclc.org/login?url=https://search.ebscohost.com/login.aspx?direct=true&AuthType=sso&db=ccm&AN=119048608&site=ehost-live&scope=site |
