## Supplementary material for "Exploring Neglected Tropical Diseases in Somalia: A Scoping Review of Research Efforts and Gaps": S2_Included Papers Characteristics

**Study Characteristics**

| Title | Author(s) | Publication year | NTD focus (type of disease) | Key findings area | Key findings points |
| --- | --- | --- | --- | --- | --- |
| Forgotten diseases: Achieving health equity to end the neglect of poverty-related diseases in Somalia | WHO | 2022 | Overall NTDs | Policy | • **Burden:** Most prevalent NTDs in Somalia are leprosy, schistosomiasis, soil-transmitted helminths, and visceral leishmaniasis; **~5–6 million** people live in highly endemic areas.  • **Leprosy:** Somalia named a **WHO global priority** in **2019**; after the MoH created an NTD section (2015), detections rose from **107 (2015)** to **2,638 (2021)** despite a 2020 COVID-related dip.  • **VL services:** **Nine** highly endemic regions are affected, but only **three** treatment centres (in **Bakool** and **Bay**) operate—indicating limited access. |
| Ending the neglect: eliminating worm infections as a public health problem in Somalia | WHO | 2022 | Schistosomiasis; Soil-transmitted helminthiases | Policy | • **Burden:** Common NTDs in Somalia include **schistosomiasis**, **soil-transmitted helminths**, **leprosy**, and **visceral leishmaniasis**; about **5-6 million** people live in highly endemic areas.  • **Mapping (2016–2017):** WHO and Somalia’s MoH identified **unexpected intestinal schistosomiasis (S. mansoni)** foci in **South West State** and **Banadir**.  • **Co-endemicity:** **Hookworm** is also common in these locations. |
| Dengue fever outbreak in Mogadishu, Somalia 2011: Co-circulation of three dengue virus serotypes | Bosa et al. | 2014 | Dengue fever | Epidemiology | • **Outbreak & surveillance:** June 2011 AFI outbreak among AMISOM peacekeepers in Mogadishu; initial tests found **82% dengue-positive**, prompting enhanced surveillance in two military hospitals.  • **Findings (Jun–Aug 2011, n=134):** **62% RT-PCR positive**, **18% MAC-ELISA positive**; multiple serotypes with **DENV-1 predominant**; co-infections present. Mostly male (**96%**), median age **32**; **60% hospitalized**; **leucopenia 87%**, **thrombocytopenia 83%**, **hemorrhagic signs 13%**.  • **Implications:** Evidence of **co-circulating DENV serotypes**, likely **under-reported** dengue in civilians, and risk of **regional spread** via rotating peacekeepers. |
| A recent case of visceral leishmaniasis in Somalia | A. Woolhead | 1995 | Visceral leishmaniasis | Epidemiology | • **Early evidence:** Clinical VL (“kala-azar”) in Somalia was first reported in **1965** among nomads north of Mogadishu (Middle Webi Shabelle).  • **Endemic focus:** **1967** work suggested an **endemic focus** of VL in that same area.  • **Continued presence:** A **confirmed case in Jan 1994** (19-year-old woman in **Baidoa**, southern Somalia) indicates ongoing transmission. |
| Visceral leishmaniasis in Somalia. Significance of IgG subclasses and of IgE response | Shiddo | 1996 | Visceral leishmaniasis | Epidemiology | • **VL immunoglobulins:** **IgG1↑ (main driver of total IgG), IgG2↓;** IgG3/IgG4 ~unchanged; anti-Leishmania antibodies present across all IgG subclasses.  • **IgE:** Total IgE similar to controls; **specific IgE** detected only in a few patients.  • **Diagnostics:** Western blot showed **consistent 74-kDa and 88-kDa bands** in all VL sera → potential markers. *(Somali controls vs Swedish: IgG1/IgG3 higher, IgG2 lower.)* |
| Visceral leishmaniasis in Somalia: prevalence of leishmanin-positive and seropositive inhabitants in an endemic area | Shiddo et al | 1995 | Visceral leishmaniasis | Epidemiology | **Village prevalence:** **26%** leishmanin+, **11%** seropositive, **14%** splenomegaly (23% among seropositives); malaria hypoendemic. Most infections were **subclinical** (only 3 ill). Bb |
| Visceral leishmaniasis in Somalia: prevalence of markers of infection and disease manifestations in a village in an endemic area | Shiddo et al | 1995 | Visceral leishmaniasis | Epidemiology | • **Historical presence:** Visceral leishmaniasis has been documented in **southern Somalia since 1934**, per reports summarized by **Moise (1955)** and **Baruffa (1965)**—evidence of long-standing endemicity. |
| Atypical perforated appendicitis secondary to schistosomiasis: a rare case report | Osoble Osman et al | 2024 | Schistosomiasis | Epidemiology | • **Etiology:** Schistosomiasis is a **rare cause of appendicitis**, often linked to urogenital bilharzia; consider it in patients from **endemic areas**.  • **Diagnosis:** Imaging (e.g., CT) may suggest appendicitis but **cannot determine the cause**; **histopathology is required** to confirm schistosomal appendicitis.  • **Management:** Standard care is **surgical appendectomy**, with **praziquantel** recommended as an **adjunct** therapy. |
| Acute kidney injury as initial presentation of visceral leishmaniasis in a young patient- A case report | Mohamed et al | 2022 | Visceral leishmaniasis | Epidemiology; Clinical Management | • **Case:** 19-year-old male from El-Barde (Bakool, Somalia) with 3-month intermittent fever, fatigue, weakness, and anorexia.  • **Context:** Leishmaniasis presents as cutaneous, mucocutaneous, visceral (VL), or post–kala-azar dermal forms.  • **Key finding:** VL can involve the kidneys; this report documents a successfully managed **acute kidney injury due to VL** (renal impairment may be acute or chronic). |
| Spinal cord Schistosomiasis: A child's case with an unsatisfactory outcome that mimicked an intramedullary neoplasm. A rare case report | Mohamed et al | 2022 | Schistosomiasis | Epidemiology; Clinical Management | • **Rare but severe:** Neuroschistosomiasis is an uncommon, life-threatening complication of schistosomiasis that can involve the brain and spinal cord; only a few cases are reported.  • **Mechanism:** Likely due to embolization of worms/eggs to CNS microcirculation; trapped eggs trigger a granulomatous inflammatory reaction causing local tissue damage and scarring.  • **Case note:** A 4-year-old child presented with one month of lower-extremity weakness—compatible with neuroschistosomiasis presentation. |
| Malaria, intestinal parasites, and schistosomiasis among Barawan Somali refugees resettling to the United States: a strategy to reduce morbidity and decrease the risk of imported infections | Miller et al | 2000 | Schistosomiasis; Soil-transmitted helminthiases | Epidemiology; Clinical Management | • **1997 screening (390/≈4,000 refugees):** 7% malaria parasitemia, 38% intestinal parasites; **2 cases** with *S. haematobium* eggs.  • **Pre-departure treatment:** Single-dose sulfadoxine–pyrimethamine (malaria) and 600 mg albendazole (≥2 yrs, non-pregnant women).  • **Impact:** Likely reduced morbidity/importation; without treatment ≈**280** malaria and **1,500** intestinal parasite cases were expected on arrival. |
| Emergence or re-emergence of visceral leishmaniasis in areas of Somalia, north-eastern Kenya, and south-eastern Ethiopia in 2000-01 | Marlet et al | 2003 | Visceral leishmaniasis | Epidemiology | • **Regional pattern:** VL has long circulated across the Somali pastoral belt (Kenya–Somalia–Ethiopia), with reports from NE Kenya since **1935**, a WWII servicemen outbreak, and community memories from the **1930s/1950s**.  • **Somalia focus:** First report **1934**; series in **1955** and **1963** (Middle Shabelle), which appears **endemic** (younger ages). Later, **39 cases** in **Kismayo (1995–96)** and a **1995** case in **Baidoa**; SE Ethiopia had few/no reports until **1984**, with an isolated **1987** case. |
| Prevalence of Schistosoma haematobium in the Koryole and Merca Districts of the Somali Democratic Republic | Koura et al | 1981 | Schistosomiasis | Epidemiology | • **Programme:** WHO-assisted Schistosomiasis Control Project launched in **1976**; **Koryole** (pilot area) and **Merca** (additional area).  • **Prevalence:** Overall **28.8%**; excluding Gedo & Bay, **Middle Juba** lowest (**25.4%**) and **Middle Shebelle** highest (**42.1%**), **P < 0.014**.  • **Distribution:** *S. haematobium* infections confined to the **Juba** and **Shebelle** river valleys; **northern Somalia** lacks rivers. |
| Molecular and epidemiologic analysis of dengue virus isolates from Somalia | Kanesa-thasan et al | 1998 | Dengue fever | Epidemiology | • **1993 outbreak (U.S. troops in Somalia):** 14 virus isolates—**12 DENV-2**, **1 DENV-3**, **1 mixed DENV-2/3**.  • **Cases:** All male, **19–39 years** (median 22), with signs/symptoms consistent with dengue.  • **Epidemiology:** Evidence of **spatial and temporal clustering**—notably **DENV-2** clusters in **Mogadishu, Kismayo, Afgoi**—with distinct genotypes circulating at different times in the same locale. |
| Retrospective study on the dengue fever outbreak in Puntland State, Somalia | Jama et al | 2024 | Dengue fever | Epidemiology | • **Positivity:** 118 of 956 suspected cases were dengue-positive (**≈12.3%**).  • **Geography:** Nugal **43.1%** (Garowe 38.1%, Burtinle 5%); Mudug **16.1%** (Galkayo), Sool **15.3%** (Las Anod), Karkar **13.6%** (Gardo), Bari **9.3%** (Bossaso), Sanaag **2.5%** (Dhahar).  • **Hospitalization (≤18 yrs):** **7** patients hospitalized (**4 female, 3 male**). |
| Antibody responses in schistosomiasis haematobium in Somalia. Relation to age and infection intensity | H.Hagi et al | 1990 | Schistosomiasis | Epidemiology | • **Prevalence:** *Schistosoma haematobium* infection was **89%** overall.  • **Age pattern:** Peak prevalence—and highest infection intensity—occurred in **10–14-year-olds**. |
| Schistosomiasis in Somalia. A parasitological and serological survey in Giohar | Cahill and Kagan | 1968 | Schistosomiasis | Epidemiology | • **Prevalence:** In Giohar District (n=125 males, 10-35 yrs), **42.4%** had urinary *S. haematobium* (Giohar **46.6%**, Mahaddei Uen **42.5%**, El Ad–El Gambole **32%**); **no** *S. mansoni* detected.  • **Exposure:** Serology positive in **36.6% (CL)**, **56.5% (BF)**, **56.1% (IFA)** → substantial prior/ongoing exposure.  • **Transmission context:** **Bulinus/Physopsis** snails observed in irrigation canals, supporting **hyperendemic** transmission and the need for detailed vector studies. |
| Visceral leishmaniasis (kala-azar) outbreak in Somali refugees and Kenyan shepherds, Kenya | Boussery et al | 2001 | Visceral leishmaniasis | Epidemiology; Clinical Management | • **Outbreak:** April 2000 VL cluster in Dadaab refugee camps (Ifo, Dagahaley, Hagadera). By Aug 2000, **34 cases** (26 DAT-positive, 8 confirmed); **L. donovani** identified; **29.4% CFR** (**10 deaths**, 6 before treatment).  • **Who/why:** Predominantly **young (median 15 yrs)**; many **recent arrivals** (Ogadeni Somali and Kenyan shepherds). **Drought-related malnutrition** likely amplified risk.  • **Response & implications:** Surveillance and diagnostic/treatment capacity strengthened; **insecticide spraying** initiated. Raised concern for a wider **VL epidemic in Somalia** given weak health infrastructure. |
| Serological evidence of dengue fever among refugees, Hargeysa, Somalia | Botros et al | 1989 | Dengue fever | Epidemiology | • **Refugee-camp outbreaks (near Hargeysa):** Malaria-like febrile illness in **1985**, with similar outbreaks over **3 consecutive years**; the **1987** episode was milder—likely due to acquired immunity.  • **Etiology:** Serology (EIA, IFA, HI) indicates **dengue virus**, with **IgM to DENV-2** and ≥4-fold titer rises in two cases; virus not isolated (sample refrigeration issues). |
| Poor patient compliance reduces the efficacy of metrifonate treatment of Schistosoma haematobium in Somalia | Adan Abdi and Gustafsson | 1989 | Schistosomiasis | Epidemiology; Clinical Management | • **Treatment & practicality:** Metrifonate is cheap, safe, and effective for *S. haematobium* (Somalia), but its **complex dosing** requires multiple visits—driving **high delivery costs** and **drop-outs**.  • **Compliance:** In two rural Somali villages, **≤48%** of patients completed the full course (participants were only **7%** of the population), indicating **poor adherence** that can undermine impact.  • **Relapse vs re-infection:** In endemic areas **>90% of relapses occur within 4–6 months** post-treatment; **re-infection typically appears after ~6 months**, making differentiation difficult. |
| Field trial of the efficacy of a simplified and standard metrifonate treatments of Schistosoma haematobium | Adan Abdi and Gustafsson | 1989 | Schistosomiasis | Epidemiology; Clinical Management | • **Regimens compared:** Standard **3×7.5 mg/kg** given two weeks apart vs. **abbreviated 3×5 mg/kg in one day**.  • **Result/implication:** The **one-day abbreviated regimen achieved similar egg-reduction and cure rates** as the standard schedule → potentially better for adherence and program logistics. |
| Schistosomiasis investigation in Somalia | The Chinese Somali Schistosomiasis Control Investigation Team | 1980 | Schistosomiasis | Epidemiology | • **Burden & symptoms:** Extremely high *S. haematobium* prevalence—**Bananie 84.7%**, **Barrie 80.4%**—with hematuria, dysuria, low-back and lower-abdominal pain common.  • **Ecology:** Long-standing endemicity in the **Shebeli/Juba river valleys**; **Bulinus abyssinicus** confirmed as the local intermediate host (no shedding from **B. forskalii**).  • **Control note:** Snail control with molluscicides/plant agents (e.g., Na-PCP) reported **~62–100%** snail reduction in trials; authors stress alignment with agriculture and community mobilization. |
| Recent Chikungunya Virus Infection in 2 Travelers Returning from Mogadishu, Somalia, to Italy, 2016 | Zammarchi et al | 2016 | Chikungunya | Epidemiology; Clinical Management | • **ravel-linked cases (Italy, 2016):** Two Somali women returning from Mogadishu developed fever, rash, and severe bilateral arthralgia/edema **17–20 days after arriving in Somalia**; both were **CHIKV-antibody positive** and managed with NSAIDs/corticosteroids.  • **Evidence of local circulation:** Relatives in Mogadishu had similar illness and media reported cases, indicating an **ongoing chikungunya outbreak** despite no CDC report for Somalia as of April 2016.  • **Likely drivers:** Possible seeding from **Kenya** plus **heavy rains/flooding** in early 2016; presence of **Aedes aegypti** vectors supports transmission. |
| Schistosomiasis -- an unusual cause of ureteral obstruction: a case history and perspective | Peter M Neal | 2004 | Schistosomiasis | Epidemiology | • **Case:** 32-year-old Somali immigrant in Wisconsin—urinary schistosomiasis confirmed by ureter/bladder **biopsy** despite negative urine; treated with **praziquantel**.  • **Clinical takeaway:** In immigrants/travelers, suspect S. haematobium even with negative urine; **tissue biopsy** may be required.  • **Public health:** U.S. lacks native snail hosts, but importation/host shifts pose risks; ensure **praziquantel access**. |
| Epidemiological investigation of dengue fever outbreak and its socioeconomic determinants in Banadir region, Somalia | Mohamed et al | 2024 | Dengue fever | Epidemiology | • **Epidemiology:** Among **735** febrile patients (56% male; majority ≤14 years), **10.8%** were DENV IgM–positive and **11.8%** NS1–positive; fever and myalgia were the commonest symptoms.  • **Outbreak:** Confirms a **dengue outbreak** in **Banadir (Oct 2022)**; most affected districts: **Deynile, Hodan, Wadajir** (211 suspected cases reported).  • **Action needed:** Strengthen **surveillance**, **laboratory diagnostics**, and **intersectoral coordination**, with clear **regulatory/financing frameworks** for outbreak response. |
| Mycetoma case series in Somalia | Dogan et al | 2024 | Mycetoma | Epidemiology; Clinical Management | • **Epidemiology/Risk:** Somalia lies in the “mycetoma belt.” Cases mostly involve rural workers (farmers, livestock handlers); lower limbs are most affected.  • **Presentation/Diagnosis:** Chronic swelling with draining sinuses and grains (diagnostic triad); diagnosis relies on biopsy/histopathology due to limited access to fungal culture.  • **Treatment/Challenges:** Long courses of antifungals/antibacterials (e.g., itraconazole, TMP-SMX; occasional surgery); high recurrence and resource constraints. Key gaps: better epidemiologic data, affordable diagnostics, comparative treatment outcomes, relapse-prevention strategies. |
| Somalia tackles leprosy and visceral leishmaniasis | Sanjeet Bagcchi | 2022 | Visceral leishmaniasis; Leprosy | Policy | • **Burden & focus:** ~**5–6 million** Somalis live in areas heavily affected by NTDs—especially **leprosy, visceral leishmaniasis, schistosomiasis,** and **STH**—with highest burden in poorer regions.  • **Progress & trends:** After the MoH created an NTD section (2015), **leprosy detections rose from 107 (2015) to 2,638 (2021)**; **~5,000 VL cases** were treated (2013–2021), mainly in **Baidoa/Bay**.  • **Gaps & response:** Control efforts face **stigma, weak infrastructure, insecurity, staff shortages, funding gaps,** and **COVID-19** disruptions. WHO supports training, detection, and NGO partnerships; experts call for **more global/government funding** and **community health worker training** to accelerate elimination. |
| Visceral Leishmaniasis, Northern Somalia, 2013-2019 | Aalto et al. | 2020 | Visceral leishmaniasis | Epidemiology; Clinical Management | • **New VL focus (Bosaso, 2013–2019):** **118** confirmed cases due to *L. donovani*; first reports in 2014 led WHO to supply rK39 tests—evidence the area may be **endemic**.  • **Who & presentation:** **91% children**, **66% male**; typical features **persistent fever, wasting, splenomegaly,** and **moderate–severe pancytopenia**.  • **Diagnosis & treatment:** Malaria first ruled out (only **2** positives); VL treated mainly with **sodium stibogluconate + paromomycin** IM (SSG alone when PARO unavailable). Authors urge **stronger surveillance**, better **diagnostics/treatment access**, and **surveys** to define endemicity. |
| Control of visceral leishmaniasis in Somalia: achievements in a challenging scenario, 2013-2015 | WHO | 2017 | Visceral leishmaniasis; Other Leishmaniasis | Epidemiology; Policy | • **High burden & transmission:** Somalia is a WHO high-burden VL country; *L. donovani* (likely anthroponotic) with vectors *Phlebotomus martini/vansomerenae*; health system remains fragile.  • **Program response:** Scale-up since 2011; national guidelines (2012); three treatment centres (Huddur, Tieglow, Baidoa); DHIS2 surveillance (2017); emerging focus in **Bosaso**.  • **Burden & who’s affected (2013–2015):** **3,112 cases (~1k/yr)**, representing **35–70%** of estimated national cases; highest incidence in **Tieglow**; mostly **children <5** with high malnutrition; surveillance is **passive** (not notifiable). |
| Epidemiology and Clinical Features of Patients with Visceral Leishmaniasis Treated by an MSF Clinic in Bakool Region, Somalia, 2004-2006 | Raguenaud et al | 2007 | Visceral leishmaniasis | Epidemiology | • **Caseload & surge:** **1,671** VL admissions at Huddur (2002–2006); ~**140/yr** until a **7× spike in 2006**. **82%** of patients were from Huddur/Tijelow; *L. donovani* genotype in 2006 matched 2002.  • **Outcomes:** Predominantly **pediatric** cases; **93.2%** clinical recovery, **3.9%** case-fatality rate.  • **Implications:** Recommend **decentralized care**, **targeted active screening**, and **community sensitization**. True burden remains uncertain given access/security limits; stronger surveillance needed. |
| Prevalence and distribution of schistosomiasis in afder and gode zone of somali region, ethiopia | Negussu et al | 2013 | Schistosomiasis | Epidemiology | • **Setting & surge:** Retrospective review of VL cases at Huddur (Bakool, Somalia), 2002–2006. After ~**140 admissions/year** (2002–2005), cases **spiked 7× in 2006** (to ~**1000**). *L. donovani* genotype in 2006 matched 2002.  • **Patients & outcomes:** **1,671** admissions; **82%** from Huddur/Tijelow; predominantly **pediatric**; **93.2%** clinical recovery, **3.9%** case-fatality.  • **Implications:** True burden unknown (access limits). Authors call for **decentralized treatment**, **targeted active screening**, and **community sensitization** to reduce morbidity and mortality. |
| A neglected disease of humans: a new focus of visceral leishmaniasis in Bakool, Somalia | Marlet et al | 2003 | Visceral leishmaniasis | Epidemiology | • **First report:** Visceral leishmaniasis documented for the first time in Somalia’s **Bakool** region.  • **Species note:** Further work needed to precisely type the parasite; may differ from cases in **Gedo** and **Wajir**.  • **Population:** Cases in Bakool are **mainly pediatric**. |
| A Late Diagnosis of Visceral Leishmaniasis Using Tru-Cut Biopsy of the Spleen and Malaria Co-Infection - A Diagnostic Challenge: A Case Report in Somalia | Hassan et al | 2023 | Visceral leishmaniasis | Epidemiology; Clinical Management | • **Case:** 24-year-old man initially misdiagnosed with malaria; later **confirmed visceral leishmaniasis (VL)** (work-up included a tru-cut spleen biopsy).  • **Complexity:** **Co-infection with** *Plasmodium* **spp.** complicated diagnosis; detailed labs and imaging were needed.  • **Implication:** Highlights frequent **misdiagnosis/delays** in resource-limited, endemic settings and the need to consider **VL in persistent febrile illness**, even when malaria is present. |
| A Case Study on Unreported First Probable Human Rabies Following Honey Badger in Somalia | Ali Osman et al. | 2024 | Rabies | Epidemiology; Clinical Management | • **Case:** Honey badger bites in rural Somalia led to fatal pediatric rabies—4-year-old girl died; 14-year-old boy developed hydrophobia after delayed care and died 48 hours post-admission.  • **Implication:** Rabies is almost always fatal once symptomatic—**immediate PEP** after any wild-animal bite is essential; highlights access gaps in rural Somalia. |
| Visceral leishmaniasis in Somalia: a review of epidemiology and access to care | Sunyoto et al | 2017 | Visceral leishmaniasis | Policy | • **Endemic & uncertain burden:** VL has persisted in southern Somalia since **1934**; true burden remains **unknown** due to conflict and poor access.  • **Control today:** **Early diagnosis and treatment**—mostly by **non-state actors**—is the only practical option, but coverage and quality are **insufficient**.  • **What is needed:** **Locally adapted, innovative care**, better **diagnostics/treatments**, and secure access making VL care **a moral imperative**. |
| Improved leprosy elimination efforts in Somalia, 2015-2021: achievements in a challenging environment and the way forward | A. Aden et al. | 2023 | Leprosy | Epidemiology; Policy | • **Data:** WHO GHO (2012-15) + National Leprosy Control Programme database (2015–21).   - • **Geography:** **77%** of 2015-21 cases in four regions—**Lower Shabelle (27%)**, **Middle Juba (20%)**, **Lower Juba (17%)**, **Middle Shabelle (14%)**. |
